# Global landscape of molecular and immunological diagnostic tests for human leishmaniasis: a systematic review and meta-analysis

**DOI:** 10.1101/2025.09.12.25335647

**Authors:** Mayron Antonio Candia-Puma, Brychs Milagros Roque-Pumahuanca, Laura Yesenia Machaca-Luque, Leydi Pola-Romero, Alexsandro Sobreira Galdino, Ricardo Andrez Machado-de-Ávila, Rodolfo Cordeiro Giunchetti, Eduardo Antonio Ferraz Coelho, Vanessa Adaui, Miguel Angel Chávez-Fumagalli

**Author notes:** These authors contributed equally to this work (Mayron Antonio Candia-Puma, Brychs Milagros Roque-Pumahuanca).

## Abstract

**Background:** Leishmaniasis constitutes a serious global public health concern. It is a complex parasitic disease characterized by a spectrum of clinical forms with varying severity, driven by host immune responses and immunopathology. Its accurate diagnosis is essential for guiding timely treatment. Yet, diagnosing leishmaniasis is challenging and requires a combination of tests.

**Methodology/Principal findings:** We conducted a systematic review and meta-analysis to evaluate the applicability and diagnostic accuracy of molecular and immunological tests for the laboratory diagnosis of human tegumentary leishmaniasis (TL) and visceral leishmaniasis (VL). We searched the PubMed database for studies published between 1990 and 2021 on leishmaniasis diagnosis. Following PRISMA statement recommendations, we included 165 publications that met the selection criteria. Among the evaluated tests, polymerase chain reaction (PCR)-based tests showed good diagnostic performance, with area under the curve values (restricted to observed FPRs, AUC_FPR_) of 0.919 and 0.965 for the diagnosis of TL and VL, respectively. For TL, serological tests showed median sensitivities ranging from 83.3% to 88.1% and median specificities ranging from 82.0% to 92.4%, whereas the leishmanin (Montenegro) skin test had a median sensitivity of 91.7% and a median specificity of 83.4%. For VL, the direct agglutination test (DAT) and enzyme-linked immunosorbent assay (ELISA) exhibited higher sensitivity (median 93.0-93.8%) than rapid diagnostic tests (RDT, 89.1%) and the immunofluorescence antibody test (IFAT, 82.0%). These four tests displayed high specificity (median 95.5-96.9%). DAT showed comparable performance to molecular tests, with an AUC_FPR_ of 0.966.

**Conclusions/Significance:** Molecular tests exhibited high accuracy in diagnosing tegumentary and visceral leishmaniasis. Nevertheless, these tests have yet to be incorporated into routine clinical practice in endemic regions, as they still require specialized technical expertise and robust laboratory infrastructure. We advocate for the development and implementation of diagnostic procedures tailored to the needs of each endemic setting, considering local contexts and available resources, wherein molecular tests could facilitate early, accurate diagnosis of leishmaniasis.

**Author Summary:** Leishmaniasis remains a significant global public health challenge, with millions of people at risk of infection, particularly in rural areas of tropical and subtropical regions. This parasitic disease presents with diverse clinical forms, ranging from skin ulcers to life-threatening mucosal or visceral organ damage. Early and accurate diagnosis of leishmaniasis is critical for timely treatment, in turn impacting disease control. Diagnosing leishmaniasis requires reliable laboratory tests. This work is a comprehensive systematic review and meta-analysis to evaluate the diagnostic accuracy of widely used molecular and immunological tests for human leishmaniasis, focusing on both tegumentary and visceral forms. Our findings reveal that PCR-based molecular tests enable highly accurate diagnosis of all forms of leishmaniasis, outperforming serological tests in many instances. Despite this, their use in endemic regions remains limited due to the need for specialized equipment and trained personnel. We emphasize the performance of immunological tests, such as DAT, LST, and ELISA, which are used in clinical and research settings and can serve as a diagnostic complement in resource-limited settings. Ultimately, this work underscores the importance of developing tailored diagnostic algorithms that incorporate molecular tests alongside simpler, field-friendly diagnostic tools to improve early diagnosis of leishmaniasis and patient outcomes in diverse epidemiological contexts.

## Introduction

Leishmaniasis is a neglected vector-borne infectious disease caused by protozoan parasites belonging to the genus *Leishmania* (Trypanosomatida: Trypanosomatidae), which encompasses over 20 species pathogenic to humans(1). The transmission occurs through the bite of infected female phlebotomine sandflies of the genus *Phlebotomus* in the Old World and *Lutzomyia* in the New World (2). Being endemic in nearly 100 countries globally, leishmaniasis poses a health risk to approximately 350 million people (3). According to the World Health Organization (WHO), an estimated 700,000 to 1 million new cases and 20,000 to 30,000 deaths due to leishmaniasis occur annually (4,5). Populations with limited resources are primarily affected by leishmaniasis, resulting in substantial financial losses. Factors such as global warming, globalization, and war/conflict are significant contributors to the spread of insect vector species to new areas, furthering the emergence of leishmaniasis (6,7). These elements collectively establish leishmaniasis as a major global public health concern.

*Leishmania* infections can present as either asymptomatic or symptomatic. When the infection becomes symptomatic, it can result in diverse clinical manifestations of variable severity, depending on the infecting *Leishmania* species, host factors (largely immune responses), and vector biology (8). Tegumentary leishmaniasis (TL) refers to a collection of skin conditions, including localized cutaneous, disseminated cutaneous, diffuse cutaneous, and mucocutaneous leishmaniasis, and post-kala-azar dermal leishmaniasis (a skin sequela of visceral leishmaniasis). The most prevalent form, localized cutaneous leishmaniasis (CL), comprises painless skin sores and ulcers on exposed body parts, such as the face, arms, and legs, with a low risk of life-threatening complications but leaving lifelong scars that result in stigma (7,9). Mucocutaneous leishmaniasis (MCL) extends to mucous membranes of the nose, mouth, or throat, causing destructive ulcers that can lead to severe disfigurement (9). Visceral leishmaniasis (VL), also known as kala-azar, represents the most severe form of the disease, affecting internal organs, with symptoms including prolonged fever, weight loss, hepatomegaly, splenomegaly, anemia, and compromised immunity; if left untreated, VL can be fatal (7). Common histopathological findings in all clinical forms of leishmaniasis include chronic granulomatous inflammation defined by the presence of macrophages harboring *Leishmania* amastigotes, plasma cells, and T lymphocytes (10).

While efforts to develop effective vaccines against human leishmaniasis are underway (11), early and accurate diagnosis and timely chemotherapy remain the major tools in the fight against the different forms of the disease. In turn, the effectiveness of these tools is influenced by the varied host-parasite interactions, immune responses, and immunopathology that underlie the different clinical manifestations of human leishmaniasis (8).

Due to the wide clinical spectrum of leishmaniasis, which often overlaps and resembles other diseases of infectious or non-infectious origin (12), the clinical management of leishmaniasis is challenging and requires specific expertise plus diagnostic tools. Its diagnosis encompasses clinical evaluation, epidemiological context, and laboratory tests. The choice of diagnostic tools is based on the clinical presentation as well as available diagnostic tests, resources, infrastructure, and technical expertise (13,14). Traditional diagnostic tools include microscopic examination of stained smears and culture (13); although being highly specific, both methods suffer from insufficient sensitivity (15–17). Immunoassays for serological diagnosis show variable sensitivity and specificity, depending on the antigen used and clinical presentation (18–23). Serological tests are of limited utility in TL diagnosis because of the typical low to moderate antibody titers, with the exception being diffuse CL that is associated with a high humoral response (24). By contrast, in VL, characterized by a high parasite load in internal organs, serological testing is of clinical utility since VL patients typically present high titers of anti-*Leishmania* antibodies (25). The leishmanin (Montenegro) skin test, which measures the delayed-type hypersensitivity response to *Leishmania* antigens, has been used to detect exposure to TL- and VL-causing *Leishmania* species and the development of cell-mediated immunity (26). A range of T cell-mediated immune responses characterizes the clinico-immunological spectrum of TL (24). At one end of the spectrum, an exaggerated cell-mediated immune response mediates immunopathology in patients with mucosal leishmaniasis, despite the ability to control the parasite load (i.e., low parasite numbers within the lesions) (24). At the other end of the spectrum, patients with diffuse CL have a lack of T cell-mediated immunity, and their lesions present a high parasite load (24). Immunohistochemistry has also proven useful as a complementary tool for the diagnosis of cutaneous and mucosal leishmaniasis (27). Parasite loads in cutaneous lesions correlate inversely with disease stage (i.e., chronic lesions harbor fewer amastigotes), which has an impact on the sensitivity of diagnostic tests (17,28).

Polymerase chain reaction (PCR)-based techniques have become important molecular tools in leishmaniasis diagnosis, as they provide high sensitivity and specificity for both detecting and identifying the infecting parasite species and are directly applied to clinical specimens (29). Molecular tools are especially valuable to help diagnose atypical and difficult-to-diagnose cases (30). Currently, the main shortcoming is that these modern diagnostic tools are mainly restricted to reference centers and research laboratories due to the requirement of infrastructure, resources, and trained personnel (31). Hence, low-resource health centers in leishmaniasis endemic areas mostly can rely only on microscopy, as they have limited access (if any) to molecular tools. This may result in delayed diagnosis, in turn impacting the timely initiation of appropriate treatment for patients. By recognizing the limitations in the confirmatory diagnosis of leishmaniasis, advances in research efforts continue to be made to develop easy-to-use diagnostic tools, which are much needed to enhance the accessibility of diagnosis and support leishmaniasis control efforts (32,33). Promising avenues include the use of portable diagnostic devices, such as handheld PCR machines (34), and the development of isothermal nucleic acid amplification tests, such as LAMP and RPA, either alone or in combination with CRISPR-Cas technology (32).

Here, we conducted a comprehensive systematic review and meta-analysis spanning 31 years of published biomedical literature to summarize the evidence on the diagnostic accuracy of molecular and immunological tests available for diagnosing human leishmaniasis. We hope that this work provides valuable insights to researchers, healthcare workers, and public health practitioners involved in leishmaniasis case management and control.

## Methods

### Study protocol

This systematic review was conducted following the Preferred Reporting Items for Systematic Reviews and Meta-Analyses (PRISMA) guidelines (S1 Table) (35). The protocol for this systematic review was registered on the International Platform of Registered Systematic Review and Meta-analysis Protocols (INPLASY) website (registration number INPLASY2023110066) and is available in full at inplasy.com (https://inplasy.com/inplasy-2023-11-0066/).

### Information sources and search strategy

We used the Medical Subject Headings (MeSH) term *“leishmaniasis”* to find terms related to the diagnosis of leishmaniasis in the biomedical literature. The results were plotted in a network diagram of the co-occurrence of MeSH terms in the VOSviewer software (version 1.6.18) (36). To select terms related to leishmaniasis diagnostic tests, we examined clusters in the network map. Additionally, a second round of searches was performed by associating each MeSH term found in the cluster analysis with the MeSH terms *“sensitivity and specificity,”* which are commonly considered indicators for evaluating the diagnostic performance of a test (37), and the MeSH term *“leishmaniasis”.* Records were retrieved from the bibliographic database PubMed (https://pubmed.ncbi.nlm.nih.gov/, last accessed on 21 December 2021) for the period 1990-2021.

### Selection criteria and data extraction

Three separate stages were engaged in the selection of studies for this review. In the initial identification phase, only studies on human patients published from 1990 to 2021 were considered. Duplicate articles, non-English publications, review papers, and meta-analyses were excluded. The subsequent screening phase involved checking titles and abstracts of the identified articles. In the eligibility/qualification phase, full-text studies highly relevant to the research question, specifically focusing on diagnostic tests for leishmaniasis, were retrieved. Data extraction from each selected study included information on the diagnostic test, total sample size, number of leishmaniasis patients, clinical characteristics of the patients, clinical form of leishmaniasis, sample type, and controls. Only studies assessing diagnostic accuracy with sensitivity and specificity measures were included, while those with limited, incomplete, or conflicting information were excluded. Traditional diagnostic tests such as microscopy, culture, and histopathology were not included because we decided to focus on molecular and immunological testing due to their increasing application in the detection of early *Leishmania* infections in clinical and research laboratories (14). Data extraction was conducted by B.M.R.-P. and independently validated by M.A.C.-P., with any discrepancies resolved through discussion and consultation with M.A.C.-F.

### Statistical analysis

Results were entered into a Microsoft Excel (version 19.0, Microsoft Corporation, Redmond, WA, USA) spreadsheet and analyzed in the R programming environment (version 4.2.3) using the *“mada”* package (version 0.5.11), available at: https://cran.r-project.org/web/packages/mada/index.html (accessed on 21 December 2021). Mada, which stands for meta-analysis of diagnostic accuracy, applies a statistical method used to combine and analyze multiple studies evaluating the accuracy of a diagnostic test or procedure. This statistical package takes into account variability between studies, examines possible sources of heterogeneity, and may incorporate methods such as subgroup analysis or meta-regression to explore factors affecting test performance (38,39). Initially, the number of true negatives (TN), false negatives (FN), true positives (TP), and false positives (FP) was analyzed separately for each diagnostic test, while the assessment of sensitivity and specificity was used to determine the diagnostic performance. Sensitivity was defined as the true positive rate, i.e., the probability that a positive test result will be obtained for a subject who has the disease, and calculated as TP/(TP + FN). Specificity was defined as the true negative rate, i.e., the probability that a negative test result will be obtained for a subject who does not have a disease or condition, and calculated as TN/(TN + FP). Also, the following ratios were calculated: the positive likelihood ratio (LR+), the negative likelihood ratio (LR-), and the diagnostic odds ratio (DOR). The LR+ was defined as the true positivity rate divided by the false positivity rate. The LR-was defined as the ratio between the false negative rate and the true negative rate. The lower the LR-value, the larger the decrease in the odds of having a condition when the test result is negative, indicating better diagnostic accuracy. The DOR of a test is the ratio of the odds of a positive test in those with disease relative to the odds of a positive test in those without disease. The DOR combines both the positive and negative likelihood ratios and provides a comprehensive measure of diagnostic accuracy. A DOR greater than 1 indicates that the test result is associated with an increased likelihood of having the condition, while a DOR less than 1 indicates a decreased likelihood (40). The summary receiver operating characteristic (sROC) curve was fitted according to the *“Reitsma”* model parameters of the *“mada”* package and used to compare the diagnostic accuracy of the tests (41). The sROC curve combines sensitivity and specificity data from individual studies to create a summary plot that represents the relationship between the true positive rate (sensitivity) and false positive rate (1-specificity) across a range of diagnostic thresholds. The sROC curve provides a visual summary of the trade-off between sensitivity and specificity of a diagnostic test, and the area under the curve (AUC) is a quantitative measure of overall test performance that also serves to identify sources of heterogeneity by visually analyzing the dispersion of individual study points around the curve, where extensive scattering indicates high heterogeneity (42). Unlike the conventional AUC, which integrates the entire ROC curve and therefore includes stretches where no study actually provided data, the AUC restricted to the observed false-positive rate (AUC_FPR_) measures test performance only across the FPR values that were truly documented. By cutting away the extrapolated portions, the AUC_FPR_ keeps the estimate grounded in evidence and prevents inflation by speculation. This makes it especially valuable when false positives are costly, as in low-prevalence diseases where even a small FPR can generate many misdiagnoses and unnecessary expenses (43). The confidence level for all calculations was set at 95%, using a continuity correction of 0.5 if relevant.

## Results

### Data sources and study selection

A flowchart of the study strategy is given in Fig. 1. A search for the MeSH term *“leishmaniasis”* was performed in PubMed, and a MeSH term co-occurrence network map was developed. The search yielded 1084 scientific articles from 1990 to 2021. The minimum number of keyword occurrences was set to five, resulting in a network graph containing 3,632 keywords (Fig. 2). Analysis of the network map revealed the formation of five main clusters. The cluster related to serological diagnostic tests (yellow color) included terms such as *“enzyme-linked immunosorbent assay,” “fluorescent antibody technique,” “direct agglutination test,”* and *“Western blot.”* In the cluster related to molecular diagnostic tests (purple color), terms such as *“polymerase chain reaction,” “real-time polymerase chain reaction,”* and *“nucleic acid amplification techniques”* were found. Furthermore, terms such as *“leishmaniasis, visceral,” “leishmaniasis,” “leishmaniasis, cutaneous,” “antiprotozoal agents,” “Leishmania infantum,” “Brazil,”* and *“Leishmania donovani”* were common (Fig. 2). The terms discovered in the initial analysis were used in a second search in the PubMed database. The new search strings were created by combining the new terms with *“leishmaniasis”* and *“sensitivity and specificity,”* as delineated in S1 File.

**Fig 1.**
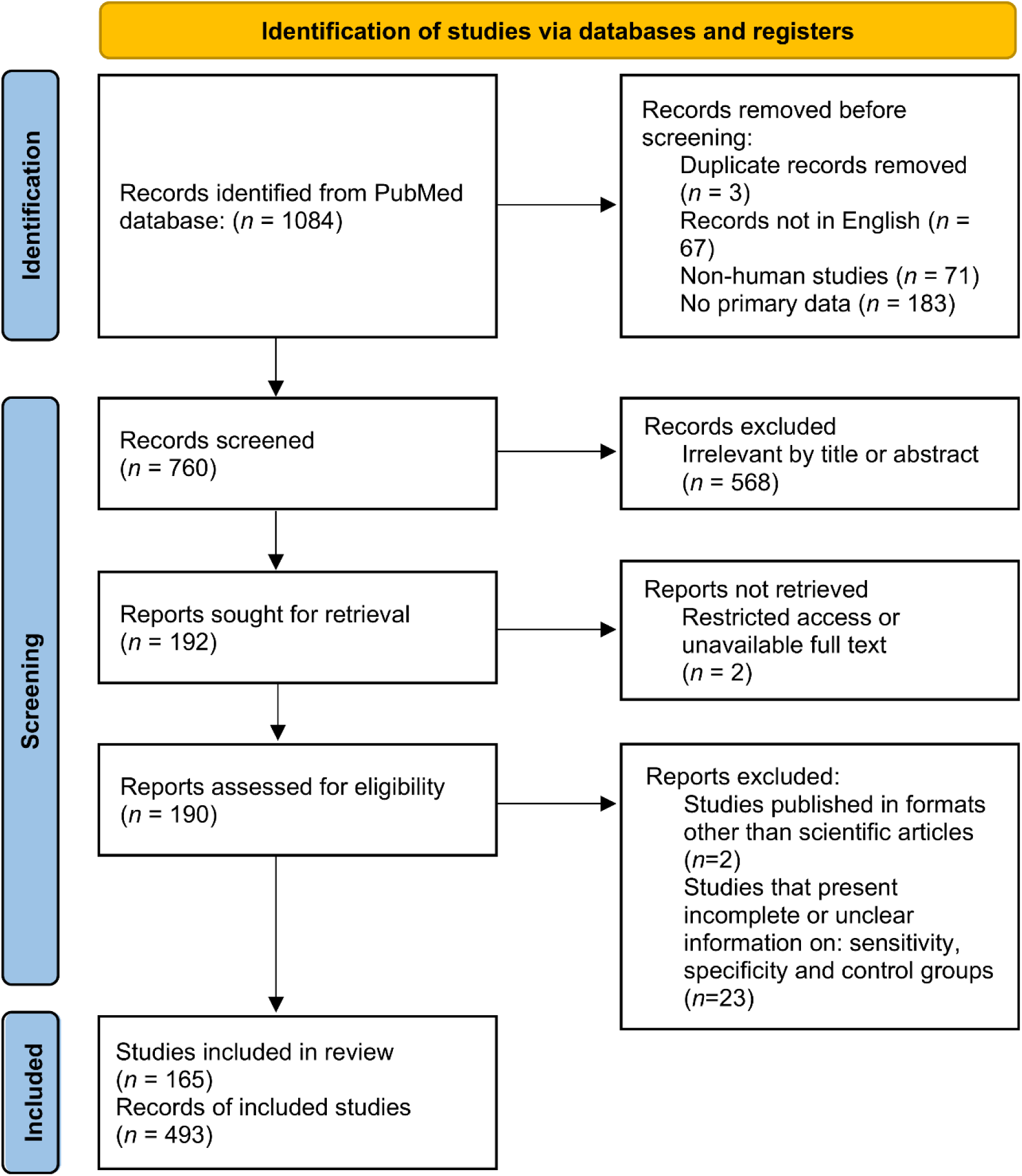
A flowchart of the systematic review and meta-analysis of the study selection process.

**Fig 2.**
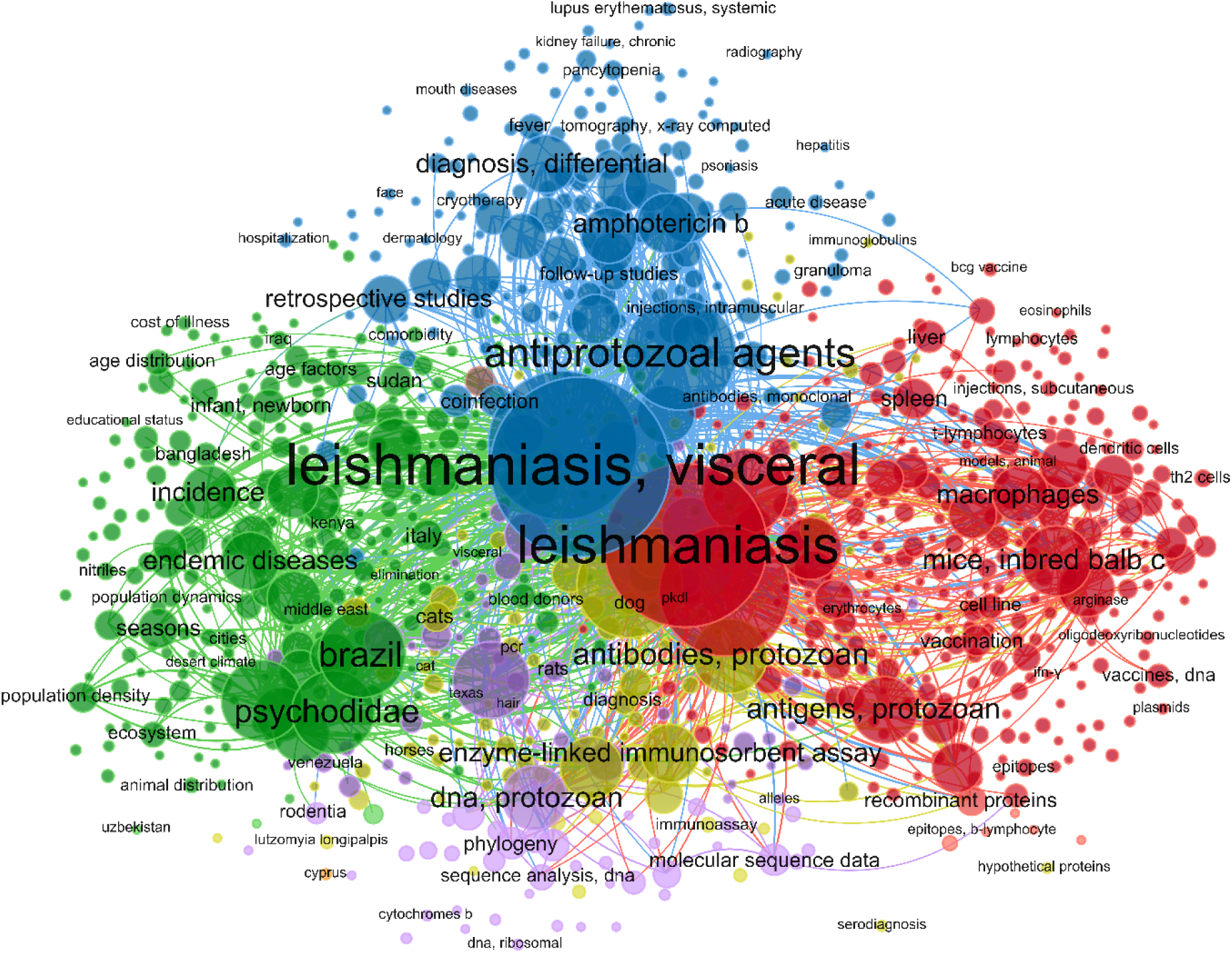
Based on the occurrence of MeSH terms in selected articles, a network diagram was created with VOSviewer using the PubMed database for different leishmaniasis diagnostic tests.

Among the retrieved studies on the performance of immunological diagnostic tests for leishmaniasis, there were 5 studies for LST, 277 for ELISA, 89 for IFAT, 10 for IHA, 78 for RDT, 138 for DAT, and 33 for WB. The retrieved studies on molecular diagnostic tests for leishmaniasis comprised 389 for PCR, 64 for qPCR, and 3 for LAMP. The three-step selection criteria we followed resulted in the exclusion of 324 articles during the identification phase, 568 articles during the screening phase, and 27 articles during the eligibility phase. As a result, 165 articles were selected for the present systematic review and meta-analysis. Some of these studies reported the evaluation of more than one diagnostic index test, resulting in a total of 493 records included in the present review. The PRISMA flow diagram summarizes record search and selection and reasons for exclusion (Fig. 1).

### Descriptive analysis of included studies

The systematic review synthesizes data from 48 studies evaluating diagnostic tests for TL, with a strong representation from Brazil (31 studies, 64.6%), followed by India, Morocco, Colombia, and others. Serum was the predominant sample type (27 studies, 56.3%), followed by biopsies and aspirates, while diagnostic methods included ELISA (20 studies), PCR (18 studies), and combinations thereof. Case-control designs were most frequent (25 studies, 52.1%), followed by cross-sectional studies (10 studies, 20.8%). The most commonly studied species were Leishmania braziliensis (26 studies) and L. donovani (7 studies), with controls rigorously selected from healthy endemic and non-endemic individuals (28 studies) and patients with Chagas disease (11 studies), malaria (6 studies), tuberculosis (4 studies), and other dermatological or systemic conditions. This comprehensive dataset underscores the heterogeneity and geographic specificity of TL diagnostics while highlighting the methodological consistency in control group selection to enhance comparative validity and clinical relevance.

The summary of findings from global studies on VL is presented in S3 Table, which emphasizes diagnostic methodologies, sample characteristics, and study designs. Of the 120 studies analyzed across 20 endemic countries, the systematic review reveals that serological methods, particularly the Direct Agglutination Test (DAT), enzyme-linked immunosorbent assay (ELISA), and rapid diagnostic tests (RDTs), were the most frequently employed diagnostic tools, collectively representing over 85% of all evaluations, while molecular techniques such as PCR were applied in approximately 15% of studies, primarily for confirmatory purposes. The majority of investigations focused on Leishmania infantum (predominantly in the Mediterranean and South America) and L. donovani (in East Africa and Southeast Asia), with sample sizes ranging from 30 to over 500 participants per study. Serum was the principal sample matrix (used in ∼80% of studies), followed by urine, peripheral blood, and bone marrow aspirates. Controls were rigorously selected to include healthy endemic and non-endemic individuals, as well as patients with diseases known to cause serological cross-reactivity (e.g., malaria, tuberculosis, and Chagas disease), thereby enabling robust specificity assessments. Study designs were predominantly case-control (70%), with prospective and comparative cohorts also well-represented, underscoring a global effort to validate diagnostic accuracy under varied epidemiological conditions.

The index tests most frequently evaluated among all included studies were ELISA, DAT, PCR, and RDT (Fig. 3A). Concerning the geographical region of the studies, Brazil, India, and Sudan were the countries for which we found the highest number of records related to diagnostic tests for leishmaniasis (Fig. 3B).

**Fig 3.**
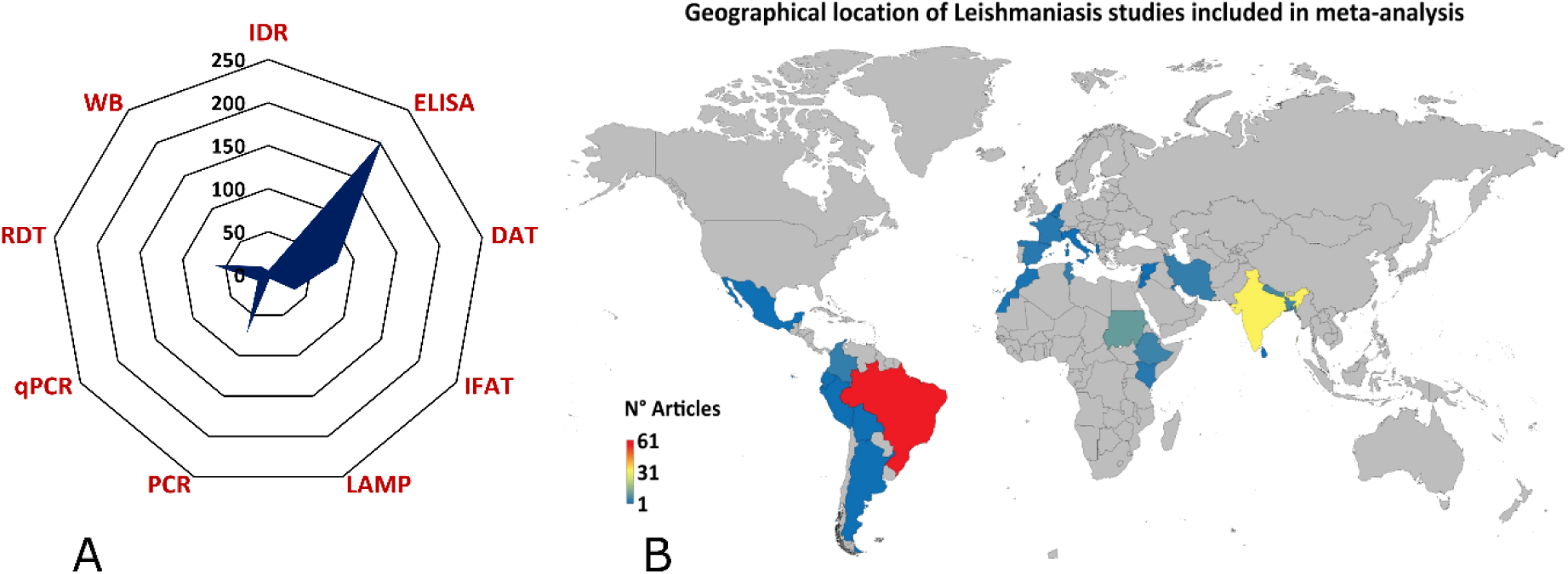
The geographical location of leishmaniasis studies. (A) The radial graph shows the type of diagnostic tests used in the leishmaniasis studies included in the meta-analysis. (B) Demographic representation of global leishmaniasis studies included in the meta-analysis (lower-blue to upper-red numbers).

### Meta-analysis of diagnostic performance of laboratory tests for leishmaniasis

#### Tegumentary leishmaniasis

##### Analyzed tests

In the context of diagnostic tests for TL, sixty-one primary reports met the eligibility criteria. There were 5 studies for LST (44–48), 20 for ELISA (45,49–67), 6 for IFAT (44,47,51,58,68,69), 3 for WB (51,59,70), 23 for PCR (44–47,71–89), and 4 for qPCR (84,90–92). Median diagnostic performance is summarized in Table 1. LST and conventional PCR yielded the greatest sensitivities (91.7% and 90.6%, respectively). The highest specificities were observed for WB (92.4%), PCR (95.1%), and qPCR (98.3%). A negative correlation between sensitivity and false-positive rate was noted for ELISA and qPCR, indicating that a greater sensitivity in these assays coincided with fewer false positives. ELISA and IFAT demonstrated DOR > 1 but remained inferior to molecular tests in discriminative ability.

**Table 1.**
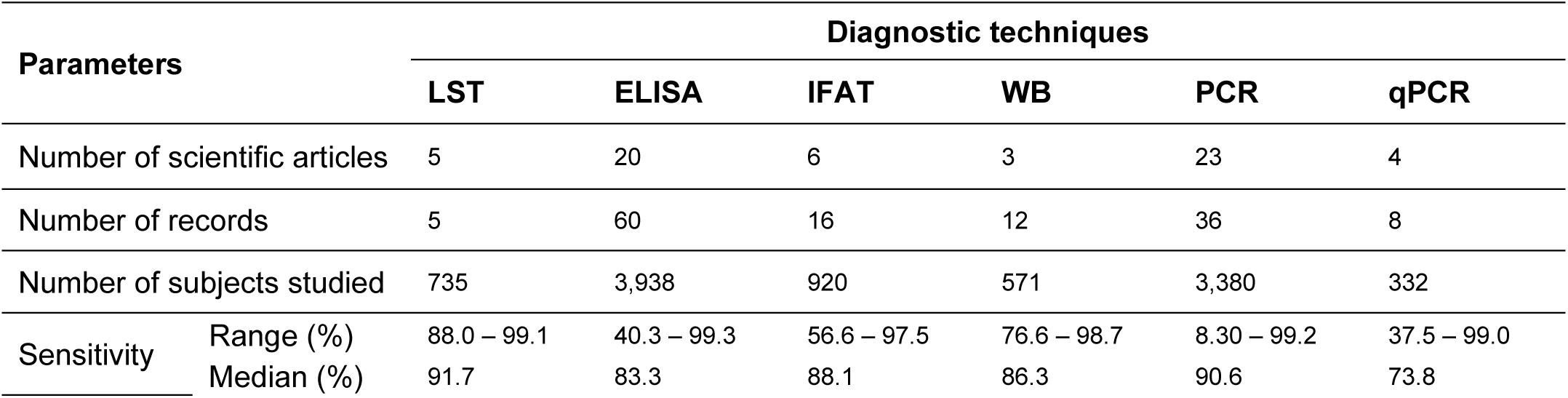
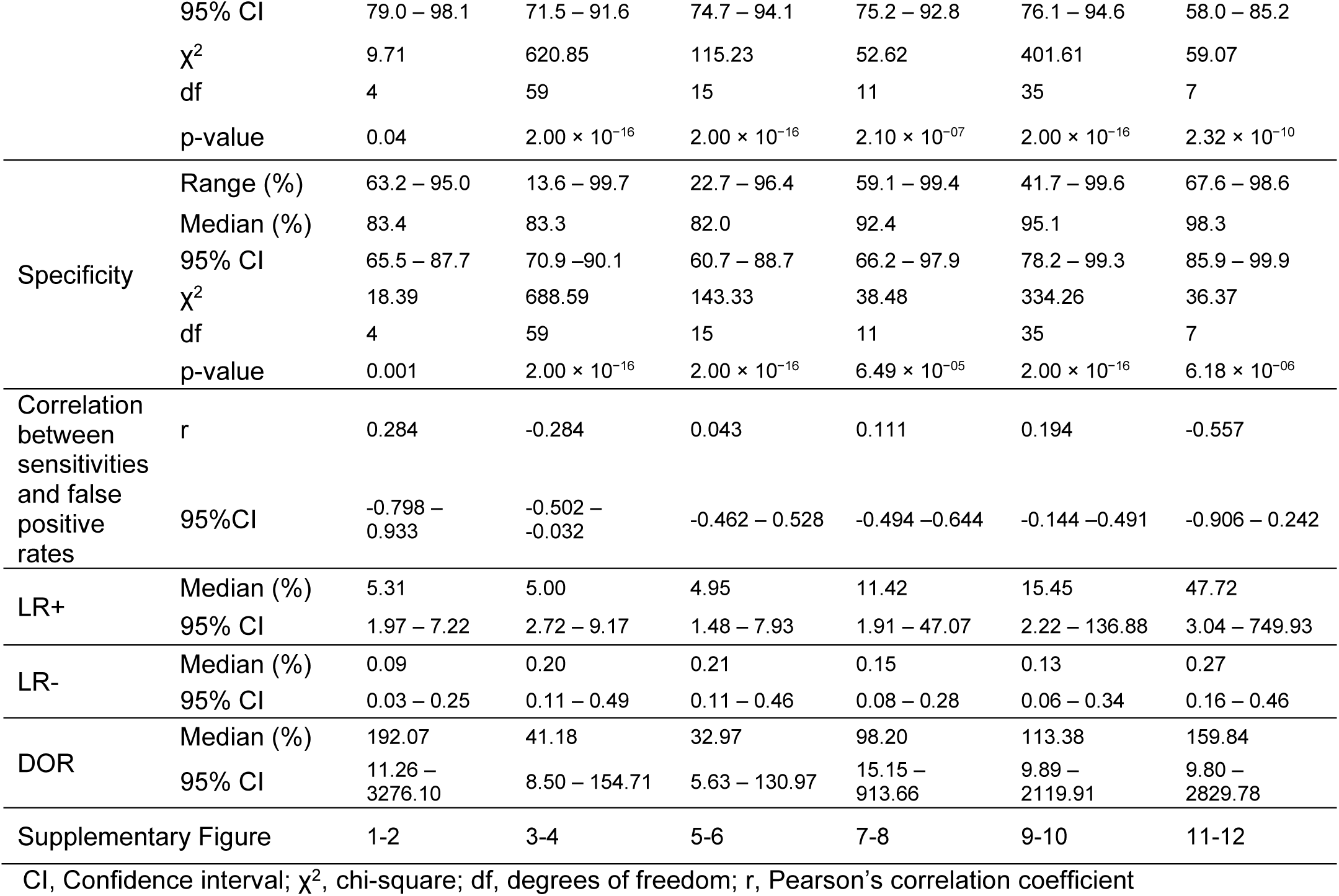
Meta-analysis of diagnostic tests for tegumentary leishmaniasis.

| Parameters |  | Diagnostic techniques |  |  |  |  |  |
| --- | --- | --- | --- | --- | --- | --- | --- |
|  |  | LST | ELISA | IFAT | WB | PCR | qPCR |
| Number of scientific articles |  | 5 | 20 | 6 | 3 | 23 | 4 |
| Number of records |  | 5 | 60 | 16 | 12 | 36 | 8 |
| Number of subjects studied |  | 735 | 3,938 | 920 | 571 | 3,380 | 332 |
| Sensitivity | Range (%) | 88.0 – 99.1 | 40.3 – 99.3 | 56.6 – 97.5 | 76.6 – 98.7 | 8.30 – 99.2 | 37.5 – 99.0 |
|  | Median (%) | 91.7 | 83.3 | 88.1 | 86.3 | 90.6 | 73.8 |
|  | 95% CI | 79.0 – 98.1 | 71.5 – 91.6 | 74.7 – 94.1 | 75.2 – 92.8 | 76.1 – 94.6 | 58.0 – 85.2 |
| | $\chi^2$ | 9.71 | 620.85 | 115.23 | 52.62 | 401.61 | 59.07 |
|  | df | 4 | 59 | 15 | 11 | 35 | 7 |
| | p-value | 0.04 | $2.00 \times 10^{-16}$ | $2.00 \times 10^{-16}$ | $2.10 \times 10^{-07}$ | $2.00 \times 10^{-16}$ | $2.32 \times 10^{-10}$ |
| Specificity | Range (%) | 63.2 – 95.0 | 13.6 – 99.7 | 22.7 – 96.4 | 59.1 – 99.4 | 41.7 – 99.6 | 67.6 – 98.6 |
|  | Median (%) | 83.4 | 83.3 | 82.0 | 92.4 | 95.1 | 98.3 |
|  | 95% CI | 65.5 – 87.7 | 70.9 – 90.1 | 60.7 – 88.7 | 66.2 – 97.9 | 78.2 – 99.3 | 85.9 – 99.9 |
| | $\chi^2$ | 18.39 | 688.59 | 143.33 | 38.48 | 334.26 | 36.37 |
|  | df | 4 | 59 | 15 | 11 | 35 | 7 |
| | p-value | 0.001 | $2.00 \times 10^{-16}$ | $2.00 \times 10^{-16}$ | $6.49 \times 10^{-05}$ | $2.00 \times 10^{-16}$ | $6.18 \times 10^{-06}$ |
| Correlation between sensitivities and false positive rates | r | 0.284 | -0.284 | 0.043 | 0.111 | 0.194 | -0.557 |
|  | 95%CI | -0.798 – 0.933 | -0.502 – -0.032 | -0.462 – 0.528 | -0.494 – 0.644 | -0.144 – 0.491 | -0.906 – 0.242 |
| LR+ | Median (%) | 5.31 | 5.00 | 4.95 | 11.42 | 15.45 | 47.72 |
|  | 95% CI | 1.97 – 7.22 | 2.72 – 9.17 | 1.48 – 7.93 | 1.91 – 47.07 | 2.22 – 136.88 | 3.04 – 749.93 |
| LR- | Median (%) | 0.09 | 0.20 | 0.21 | 0.15 | 0.13 | 0.27 |
|  | 95% CI | 0.03 – 0.25 | 0.11 – 0.49 | 0.11 – 0.46 | 0.08 – 0.28 | 0.06 – 0.34 | 0.16 – 0.46 |
| DOR | Median (%) | 192.07 | 41.18 | 32.97 | 98.20 | 113.38 | 159.84 |
|  | 95% CI | 11.26 – 3276.10 | 8.50 – 154.71 | 5.63 – 130.97 | 15.15 – 913.66 | 9.89 – 2119.91 | 9.80 – 2829.78 |
| Supplementary Figure |  | 1-2 | 3-4 | 5-6 | 7-8 | 9-10 | 11-12 |

CI, Confidence interval; $\chi^2$ , chi-square; df, degrees of freedom; r, Pearson's correlation coefficient

### Other diagnostic tests

Two studies on the performance of RDT (91,93) and two studies on DAT (94,95) for TL diagnosis were selected. According to the established criteria, at least 5 studies were needed for the analysis with a p-value < 0.05. Therefore, no analysis could be performed regarding these diagnostic tests.

### Summary ROC curves (sROC)

Data comparison of diagnostic tests for TL (LST, ELISA, IFAT, WB, PCR, and qPCR) was carried out using a sROC curve analysis (Fig. 4). The AUC calculated for each diagnostic test showed that WB, PCR, and qPCR offered the best diagnostic performance (Fig. 4). Furthermore, PCR performed robustly for TL when the AUC was restricted to the observed false positive rate (AUC_FPR_ = 0.919, Fig. 4).

**Fig 4.**
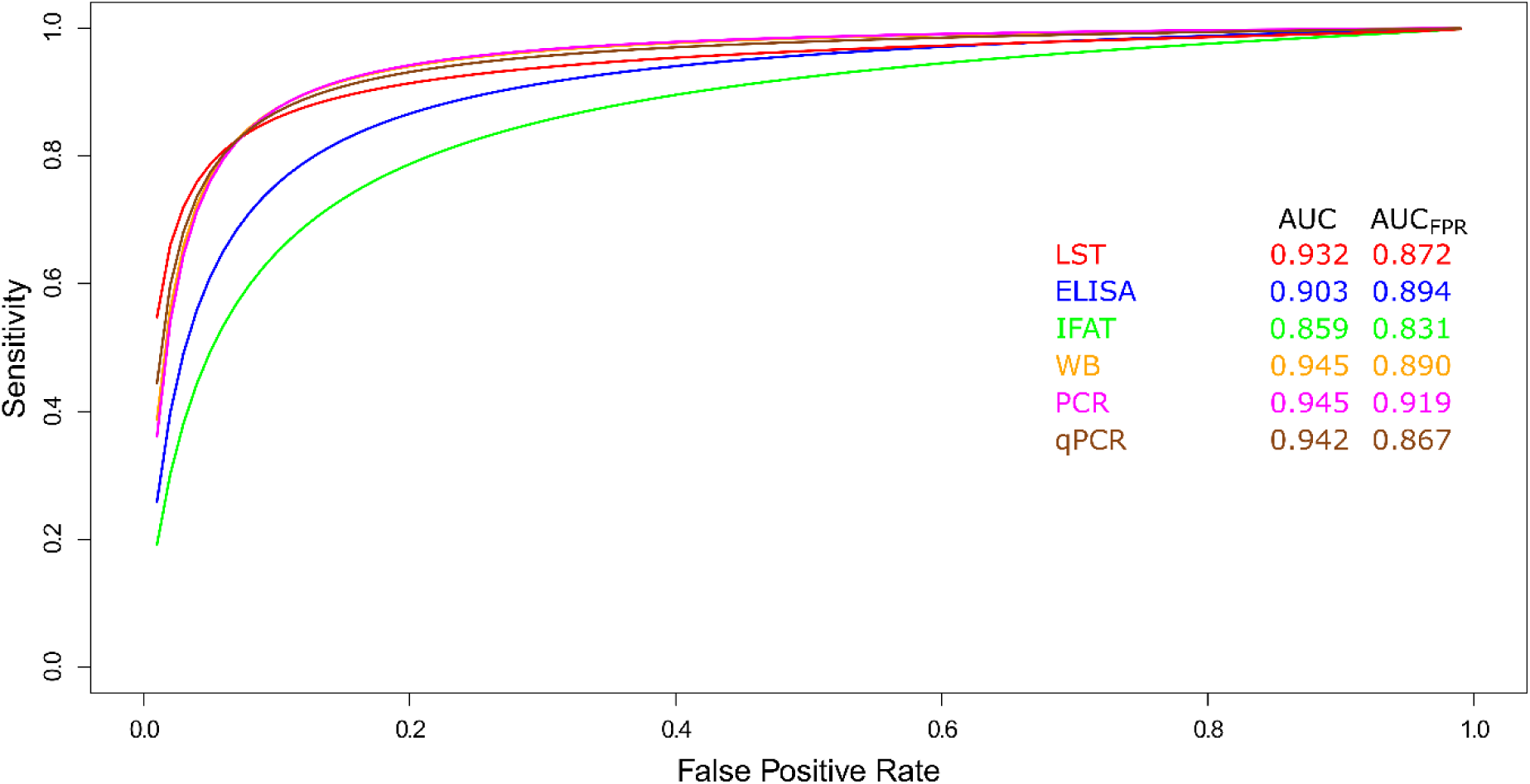
Meta-analysis of diagnostic test accuracy studies for tegumentary leishmaniasis. Summary receiver operating characteristic (sROC) curve of sensitivity (true positive rate) against false positive rate (1 - specificity). Comparison between LST, ELISA, IFAT, WB, PCR, and qPCR methods used in the diagnosis of tegumentary leishmaniasis.

### Visceral leishmaniasis Analyzed tests

Concerning diagnostic tests for VL, there were 23 included studies using DAT (50,96–139), 58 for ELISA (50,59,67,114–117,119–121,130,133,134,140–184), 12 for IFAT (50,96,114,115,123,126,130,169,185–188), 33 for RDT (96,100,114,115,120,123,125–128,130–133,135,138,139,144,156,178,182–187,189–195), 3 for LAMP (140,191,194), 22 for PCR (75,83,123,157,177,191,194–209), and 5 for qPCR (140,164,187,208,210). A detailed examination of the diagnostic performance of these tests in the included studies is shown in Table 2. DAT showed the most favorable accuracy profile, with a median sensitivity of 93.8%, a median specificity of 96.9%, and the highest AUC_FPR_ (0.966). ELISA and qPCR offered comparable sensitivities (92.9%) but lower specificities than DAT. LAMP, PCR, and qPCR provided the greatest specificities (> 96%). Lastly, DAT, ELISA, IFAT, and RDT displayed an inverse relation between sensitivity and false-positive rate.

**Table 2.**
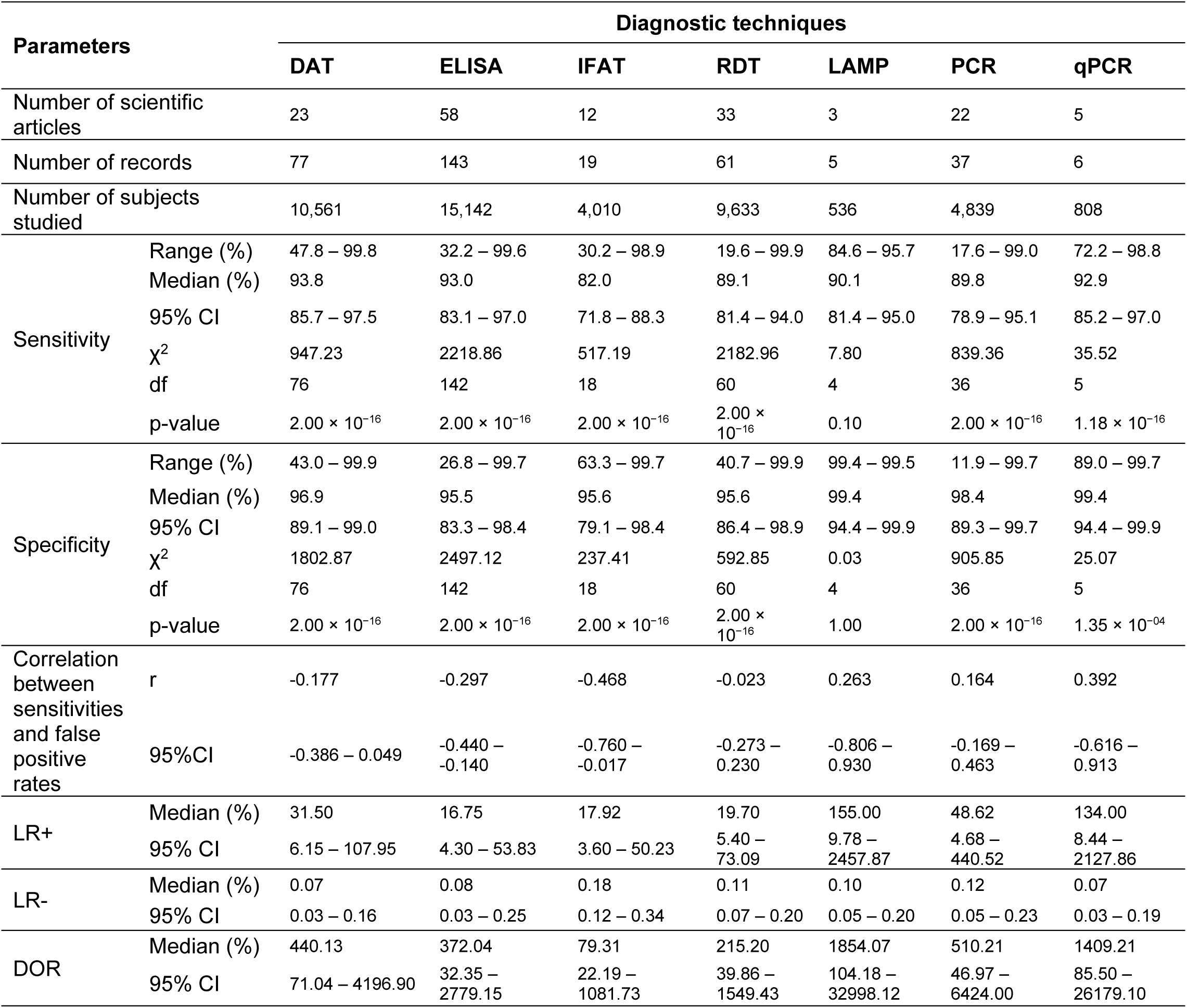
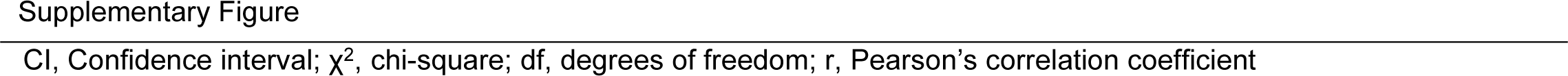
Meta-analysis of diagnostic tests for visceral leishmaniasis.

| Parameters |  | Diagnostic techniques |  |  |  |  |  |  |
| --- | --- | --- | --- | --- | --- | --- | --- | --- |
|  |  | DAT | ELISA | IFAT | RDT | LAMP | PCR | qPCR |
| Number of scientific articles |  | 23 | 58 | 12 | 33 | 3 | 22 | 5 |
| Number of records |  | 77 | 143 | 19 | 61 | 5 | 37 | 6 |
| Number of subjects studied |  | 10,561 | 15,142 | 4,010 | 9,633 | 536 | 4,839 | 808 |
| Sensitivity | Range (%) | 47.8 – 99.8 | 32.2 – 99.6 | 30.2 – 98.9 | 19.6 – 99.9 | 84.6 – 95.7 | 17.6 – 99.0 | 72.2 – 98.8 |
|  | Median (%) | 93.8 | 93.0 | 82.0 | 89.1 | 90.1 | 89.8 | 92.9 |
|  | 95% CI | 85.7 – 97.5 | 83.1 – 97.0 | 71.8 – 88.3 | 81.4 – 94.0 | 81.4 – 95.0 | 78.9 – 95.1 | 85.2 – 97.0 |
| | $\chi^2$ | 947.23 | 2218.86 | 517.19 | 2182.96 | 7.80 | 839.36 | 35.52 |
|  | df | 76 | 142 | 18 | 60 | 4 | 36 | 5 |
| | p-value | $2.00 \times 10^{-16}$ | $2.00 \times 10^{-16}$ | $2.00 \times 10^{-16}$ | $2.00 \times 10^{-16}$ | 0.10 | $2.00 \times 10^{-16}$ | $1.18 \times 10^{-16}$ |
| Specificity | Range (%) | 43.0 – 99.9 | 26.8 – 99.7 | 63.3 – 99.7 | 40.7 – 99.9 | 99.4 – 99.5 | 11.9 – 99.7 | 89.0 – 99.7 |
|  | Median (%) | 96.9 | 95.5 | 95.6 | 95.6 | 99.4 | 98.4 | 99.4 |
|  | 95% CI | 89.1 – 99.0 | 83.3 – 98.4 | 79.1 – 98.4 | 86.4 – 98.9 | 94.4 – 99.9 | 89.3 – 99.7 | 94.4 – 99.9 |
| | $\chi^2$ | 1802.87 | 2497.12 | 237.41 | 592.85 | 0.03 | 905.85 | 25.07 |
|  | df | 76 | 142 | 18 | 60 | 4 | 36 | 5 |
| | p-value | $2.00 \times 10^{-16}$ | $2.00 \times 10^{-16}$ | $2.00 \times 10^{-16}$ | $2.00 \times 10^{-16}$ | 1.00 | $2.00 \times 10^{-16}$ | $1.35 \times 10^{-04}$ |
| Correlation between sensitivities and false positive rates | r | -0.177 | -0.297 | -0.468 | -0.023 | 0.263 | 0.164 | 0.392 |
|  | 95%CI | -0.386 – 0.049 | -0.440 – -0.140 | -0.760 – -0.017 | -0.273 – 0.230 | -0.806 – 0.930 | -0.169 – 0.463 | -0.616 – 0.913 |
| LR+ | Median (%) | 31.50 | 16.75 | 17.92 | 19.70 | 155.00 | 48.62 | 134.00 |
|  | 95% CI | 6.15 – 107.95 | 4.30 – 53.83 | 3.60 – 50.23 | 5.40 – 73.09 | 9.78 – 2457.87 | 4.68 – 440.52 | 8.44 – 2127.86 |
| LR- | Median (%) | 0.07 | 0.08 | 0.18 | 0.11 | 0.10 | 0.12 | 0.07 |
|  | 95% CI | 0.03 – 0.16 | 0.03 – 0.25 | 0.12 – 0.34 | 0.07 – 0.20 | 0.05 – 0.20 | 0.05 – 0.23 | 0.03 – 0.19 |
| DOR | Median (%) | 440.13 | 372.04 | 79.31 | 215.20 | 1854.07 | 510.21 | 1409.21 |
|  | 95% CI | 71.04 – 4196.90 | 32.35 – 2779.15 | 22.19 – 1081.73 | 39.86 – 1549.43 | 104.18 – 32998.12 | 46.97 – 6424.00 | 85.50 – 26179.10 |

### Other diagnostic tests

Two studies, one concerning WB (59) and one on IHA (185), were selected for VL diagnosis. According to the established criteria, at least 5 studies with a p-value < 0.05 were required for the analysis. As a result, no analysis of these diagnostic tests could be conducted.

### Summary ROC curves (sROC)

Data for VL diagnostic tests (DAT, ELISA, IFAT, RDT, LAMP, PCR, and qPCR) were compared using a sROC curve analysis (Fig. 5). All diagnostic tests showed an overall good performance (AUC > 0.9, Fig. 5), with LAMP standing out (AUC = 0.990, Fig. 5). Yet, when the AUC was limited to the observed false positive rate, LAMP showed lower accuracy (AUC_FPR_ = 0.499, Fig. 5) than the other tests (AUC_FPR_ range between 0.899 and 0.966, Fig. 5). This discrepancy arises because the AUC_FPR_ isolates the test’s ability to minimize false positives, revealing a potential weakness of LAMP in distinguishing between diseased and non-diseased individuals, despite overall good performance. On the other hand, DAT performed properly for VL (AUC = 0.975 and AUC_FPR_ = 0.966; Fig. 5), and its results were comparable to those of PCR tests (AUC = 0.967 and AUC_FPR_ = 0.965; Fig. 5).

**Fig 5.**
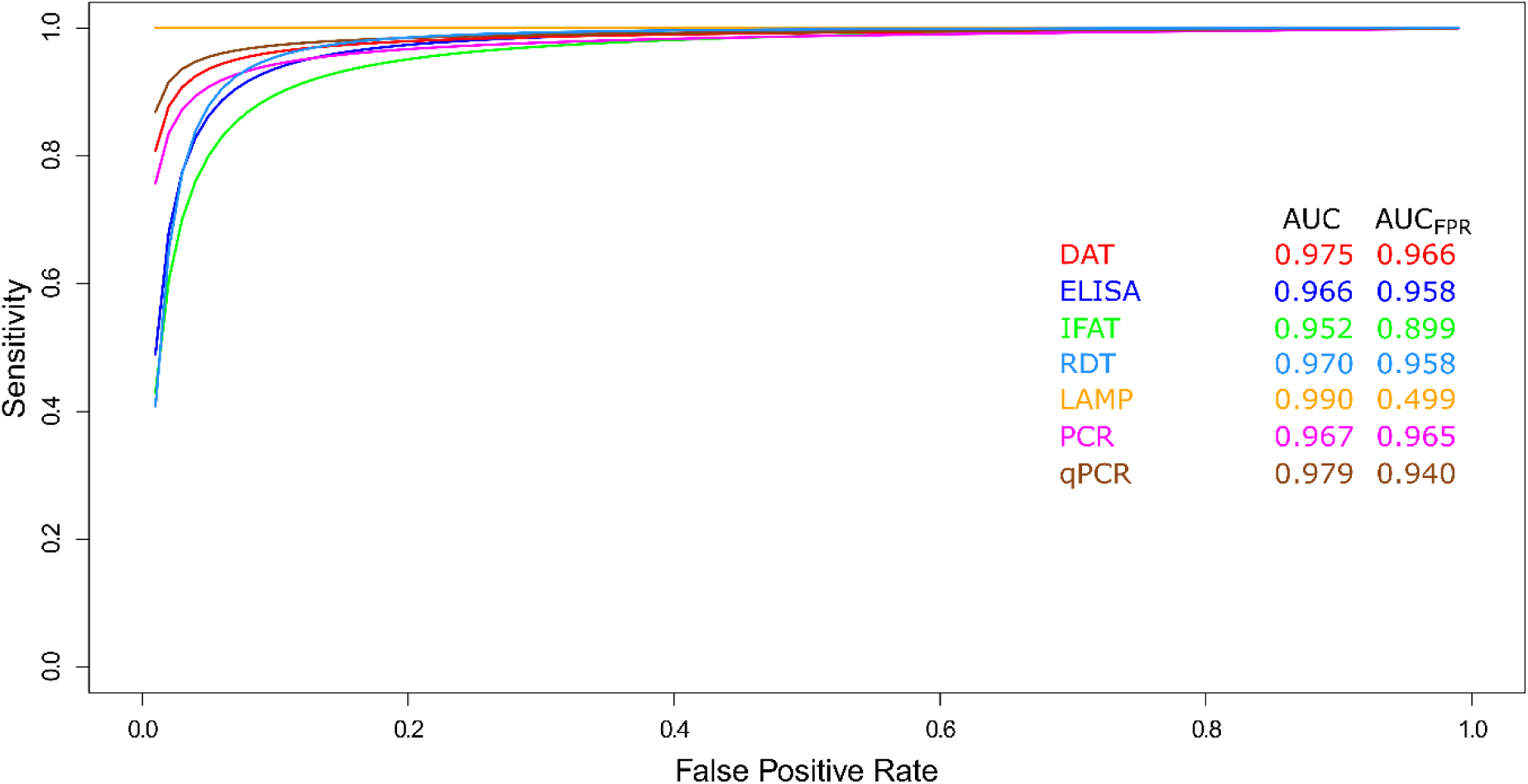
Meta-analysis of diagnostic test accuracy studies for visceral leishmaniasis. Summary receiver operating characteristic (sROC) curve of sensitivity (true positive rate) against false positive rate (1 - specificity). Comparison between DAT, ELISA, IFAT, LAMP, PCR, qPCR, and RDT methods used in the diagnosis of visceral leishmaniasis.

## Discussion

According to the WHO, early diagnosis and prompt treatment for leishmaniasis reduce disease prevalence, prevent disabilities and death, help lower transmission, and facilitate the estimation of disease burden and the development of interventions for disease control (5). In this context, the WHO Roadmap for Neglected Tropical Diseases 2021-2030 highlights diagnosis as a key pillar in the efforts to control infectious diseases such as leishmaniasis, emphasizing the need to strengthen diagnostic capacity to achieve the proposed objectives (211). To this end, the implementation of accurate and accessible molecular and/or immunological diagnostic tests is crucial for improving the early detection and clinical management of leishmaniasis cases, thereby contributing to global efforts towards the effective control and eventual elimination of leishmaniasis as a public health problem.

### Summary of main findings

This study constitutes a comprehensive systematic review and meta-analysis that evaluates the global landscape of molecular and immunological diagnostic tests for human leishmaniasis. The findings reveal a complex diagnostic scenario, characterized by significant geographical and methodological heterogeneity, as well as clear trends in test performance depending on the clinical form of the disease.

For TL, the results demonstrate that molecular tests, particularly conventional PCR and qPCR, exhibit the best diagnostic performance profile. PCR demonstrated exceptional median sensitivity (90.6%) and high specificity (95.1%), as supported by an AUC in the sROC analysis, confirming its robustness. Although evaluated in a smaller number of studies, qPCR showed the highest reported specificity (98.3%), positioning it as an ideal confirmatory tool in contexts where specificity is paramount, such as in areas with co-endemic diseases (212). In contrast, serological tests such as ELISA and IFAT, although widely used, have shown inferior discriminatory ability (DOR) compared to molecular techniques, suggesting their utility may be more limited to initial screening studies or in resource-limited settings where PCR is not available.

In the case of VL, the findings are different. Serological tests demonstrated outstanding performance. DAT emerged as the most accurate test, with median sensitivity and specificity of 93.8% and 96.9%, respectively, and an AUC_FPR_ of 0.966, indicating an excellent ability to minimize false positives. ELISA and RDT also demonstrated high sensitivities (∼93%), reinforcing the central role of serology in VL diagnosis, particularly in the field where rapid results are required. Although molecular techniques like LAMP showed a near-perfect overall AUC (0.990), its AUC_FPR_ was significantly lower (0.499). This discrepancy suggests that, despite its high potential, the LAMP technique may present challenges in terms of specificity in real-world practice, as observed in the included studies.

### Interpretation of findings in the context of existing literature

The diagnostic performance of molecular and immunological tests for leishmaniasis has been a subject of intense research in biomedical literature. Various studies have highlighted that in immunocompromised patients, serological tests tend to exhibit reduced sensitivity due to the inability of these individuals to mount an effective immune response against *Leishmania* parasites (213). This often leads to false-negative results in early stages of the disease or in cases harboring low parasite loads. By contrast, PCR has been recognized as a more reliable test, capable of directly detecting the parasite’s genetic material, even in low-level infections, in human host tissue samples (14). However, the applicability of PCR in resource-limited settings remains a challenge due to its requirement for specialized equipment and trained personnel, which limits its use in rural areas or countries with underdeveloped healthcare infrastructures.

### Strengths and limitations

This review’s main strength is the thorough examination of the published literature, which made it possible to assess the accuracy of leishmaniasis diagnostic tests across various endemic geographic regions. The use of PRISMA guidelines for conducting the systematic review ensured transparency and reproducibility of the procedures, reinforcing the reliability of the findings (214).

Additionally, prospectively registering the protocol in INPLASY allowed for a standardized methodology, ensuring the review maintains its relevance over time (215). The inclusion of diverse diagnostic techniques—both serological and molecular tests—enriched the discussion, offering a broad perspective on the diagnostic performance of available tests across different populations and settings. This strengthens the validity of the conclusions and their applicability to real-world diagnostic challenges in leishmaniasis.

There are important limitations of this review. The inherent heterogeneity among the included studies—evidenced by differences in population study designs (acute/chronic stages of disease, New/Old World clinical forms), sampling methods, controls used, composite reference standards, and reference tests (microscopy/PCR/culture) used—hindered direct comparisons and prevented the assessment of diagnostic test performance according to specific clinical-epidemiological contexts. A second limitation relates to the issue of possible publication bias. For the selection of studies included in this review, we considered journal articles published in English as an eligibility criterion. In addition, there is potential underrepresentation of studies with negative findings. Under-reporting is also possible since we found a scarcity of data from endemic regions with a weak publication record (e.g., sub-Saharan Africa, rural areas of Asia). While we did not formally test for publication bias, we believe that the overall estimation of the diagnostic test accuracy itself was not directly affected. However, we issue a warning regarding the conclusions of this review’s transregional generalizability.

### Recommendations for standardization and future research

To improve the accuracy of leishmaniasis diagnostic tests, it is essential to standardize methods and result interpretation criteria. Multicenter prospective studies are recommended, including a variety of *Leishmania* species and different biological samples to provide a more comprehensive assessment of test performance (216). Furthermore, the development of standardized reference panels would allow for objective comparisons between diagnostic tests.

### Health policy perspectives and technological development

The findings of this review could influence public health policy decisions, guiding investments in research and the development of new diagnostic technologies. Global health policies, such as those proposed by the WHO, should promote the integration of rapid and accessible diagnostic tests into leishmaniasis control programs, particularly in rural areas and countries with limited infrastructure. Investment in research on new tests, such as low-cost PCR-based methods, could be key to improving the diagnosis and treatment of this neglected disease.

### Conclusions

The results showed that while serological assays (ELISA, IFAT) represent an accessible alternative in resource-limited settings, their variable sensitivity compromises their reliability for the early diagnosis of leishmaniasis. In contrast, molecular methods (PCR) demonstrate superior sensitivity (>90%) and a critical capacity to detect low parasite loads, establishing themselves as indispensable pillars for timely clinical management. It is crucial to consider that in recent years, the SARS-CoV-2 pandemic has driven the global acquisition of equipment and infrastructure for molecular testing, such as PCR, an advance that could mitigate historical operational barriers in remote endemic areas. Nevertheless, major challenges persist, such as methodological heterogeneity and the lack of global standardization. These findings reinforce the public health imperative to develop integrated strategies—combining serology for screening and PCR for confirmation—alongside policies that capitalize on this new diagnostic capacity, drive accessible technological innovation, and prioritize equity. Only then can the WHO’s goal of controlling this neglected disease be realized, transforming technical precision into better human outcomes for the most vulnerable communities.

## Author Contributions

Conceptualization: M.A.C.-P. and M.A.C.-F.; data curation: M.A.C.-P., B.M.R.-P., L.Y.M.-L., and L.P.-R.; formal analysis: M.A.C.-P. and M.A.C.-F.; funding acquisition: M.A.C.-P. and M.A.C.-F.; investigation: V.A., A.S.G., R.C.G., R.A.M.-d.-Á., and E.A.F.C.; methodology: M.A.C.-P. and M.A.C.-F.; writing—review and editing: V.A., A.S.G., R.C.G., R.A.M.-d.-Á., E.A.F.C., and M.A.C.-F. All authors have read and agreed to the published version of the manuscript.

## Institutional Review Board Statement

Not applicable.

## Informed Consent Statement

Not applicable.

## Data Availability Statement

Not applicable.

## Acknowledgments

Not applicable.

## Supplementary Figure legends

**S1 Fig.**
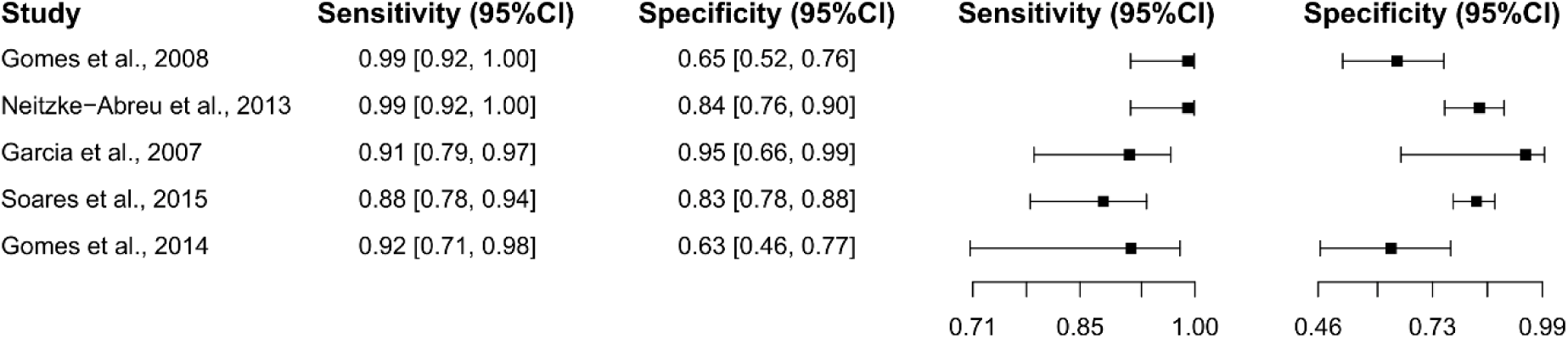
Study data and paired forest plot of the sensitivity and specificity of the leishmanin skin test (LST) in tegumentary leishmaniasis diagnosis. Data from each included study (44–48) are summarized. Sensitivity and specificity are reported with a mean (95% confidence limits). The forest plot depicts the estimated sensitivity and specificity (black squares) and their 95% confidence limits (horizontal black line).

**S2 Fig.**
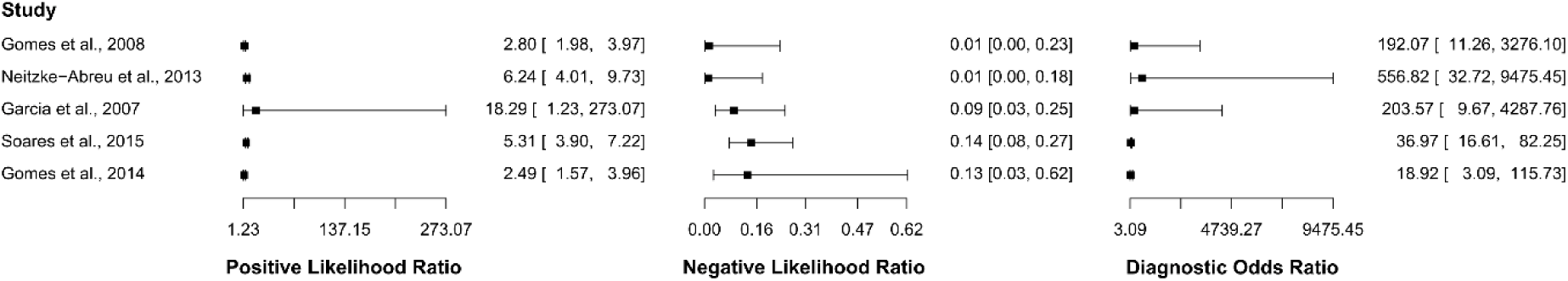
Study data and paired forest plot of the positive likelihood ratio, negative likelihood ratio, and diagnostic odds ratio of the leishmanin skin test (LST) in the diagnosis of tegumentary leishmaniasis. The positive likelihood ratio, negative likelihood ratio, and diagnostic odds ratio are reported with a mean (95% confidence limits) for the included studies (44–48).

**S3 Fig.**
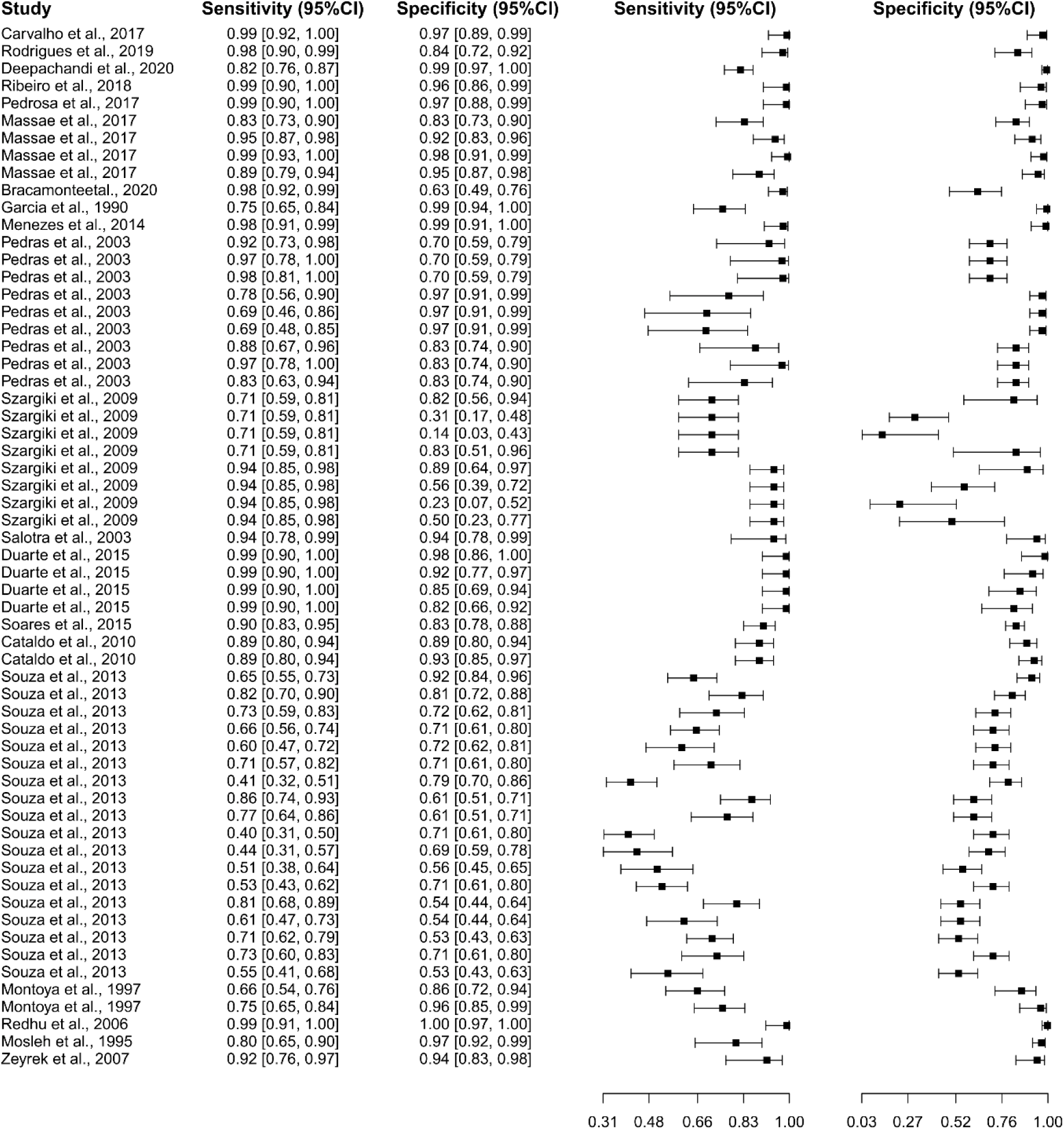
Study data and paired forest plot of the sensitivity and specificity of enzyme-linked immunosorbent assay (ELISA) in tegumentary leishmaniasis diagnosis. Data from each included study (45,49–67) are summarized. Sensitivity and specificity are reported with a mean (95% confidence limits). The forest plot depicts the estimated sensitivity and specificity (black squares) and their 95% confidence limits (horizontal black line).

**S4 Fig.**
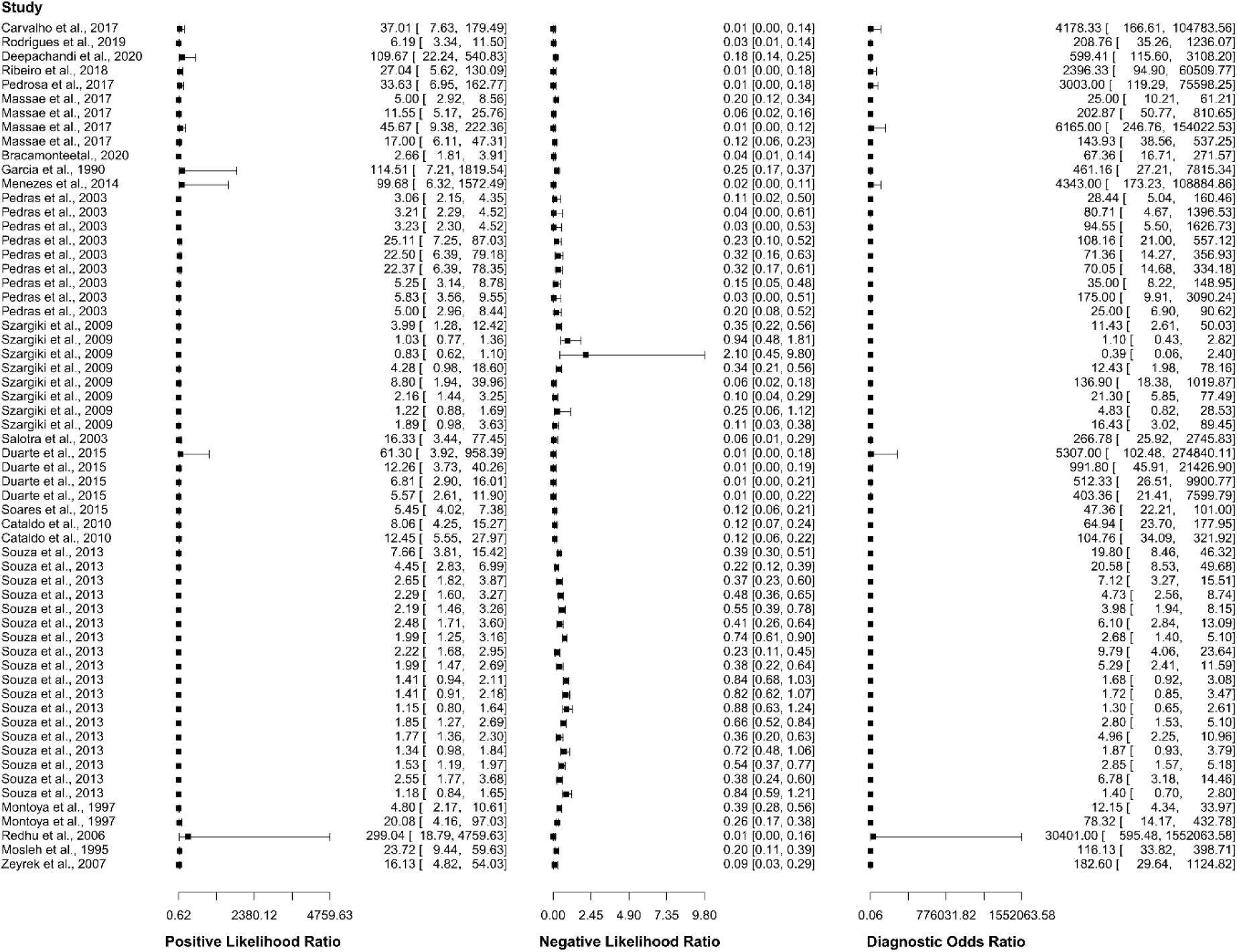
Study data and paired forest plot of the positive likelihood ratio, negative likelihood ratio, and diagnostic odds ratio of enzyme-linked immunosorbent assay (ELISA) in the diagnosis of tegumentary leishmaniasis. The positive likelihood ratio, negative likelihood ratio, and diagnostic odds ratio are reported with a mean (95% confidence limits) for the included studies (45,49–67).

**S5 Fig.**
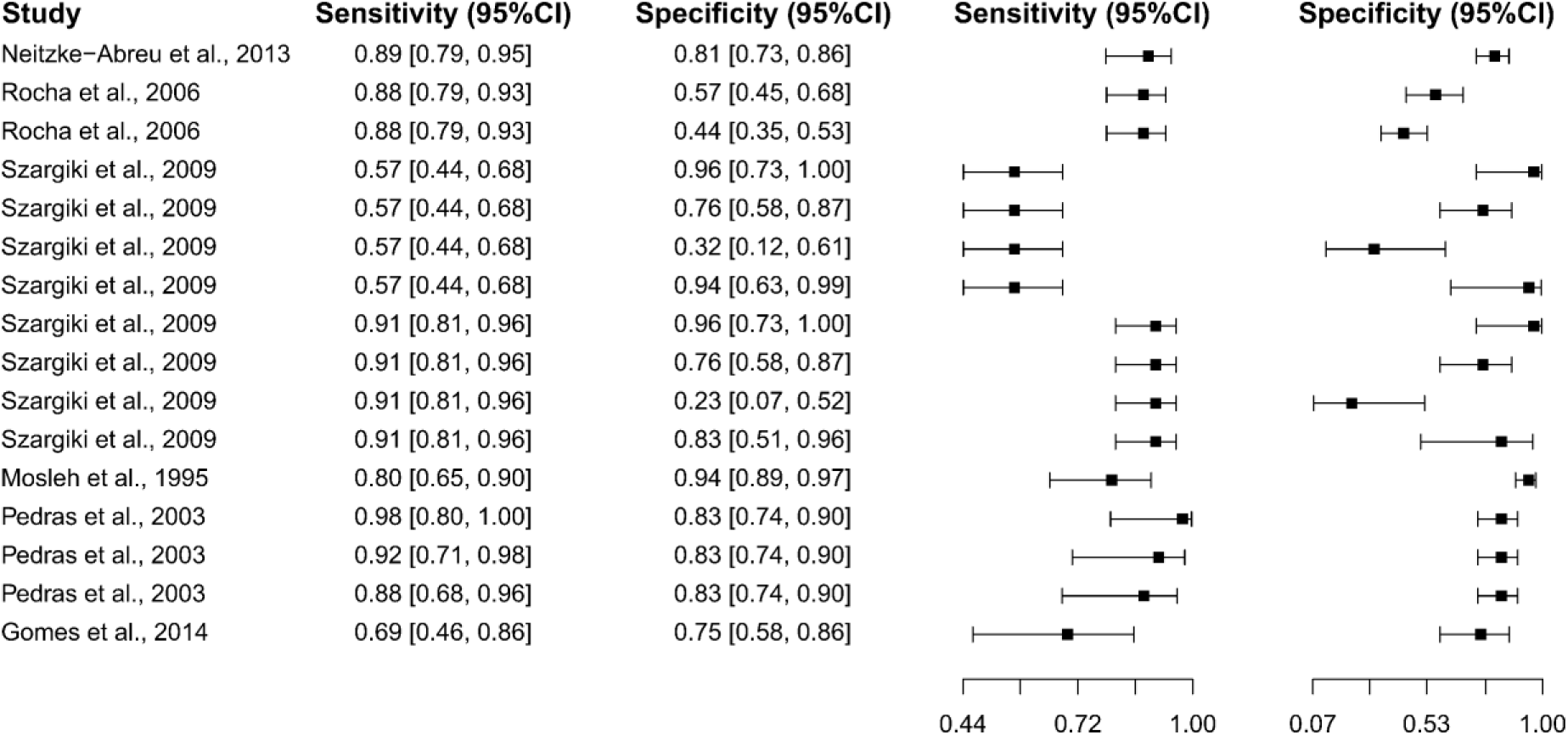
Study data and paired forest plot of the sensitivity and specificity of indirect fluorescent antibody test (IFAT) in tegumentary leishmaniasis diagnosis. Data from each included study (44,47,51,58,68,69) are summarized. Sensitivity and specificity are reported with a mean (95% confidence limits). The forest plot depicts the estimated sensitivity and specificity (black squares) and their 95% confidence limits (horizontal black line).

**S6 Fig.**
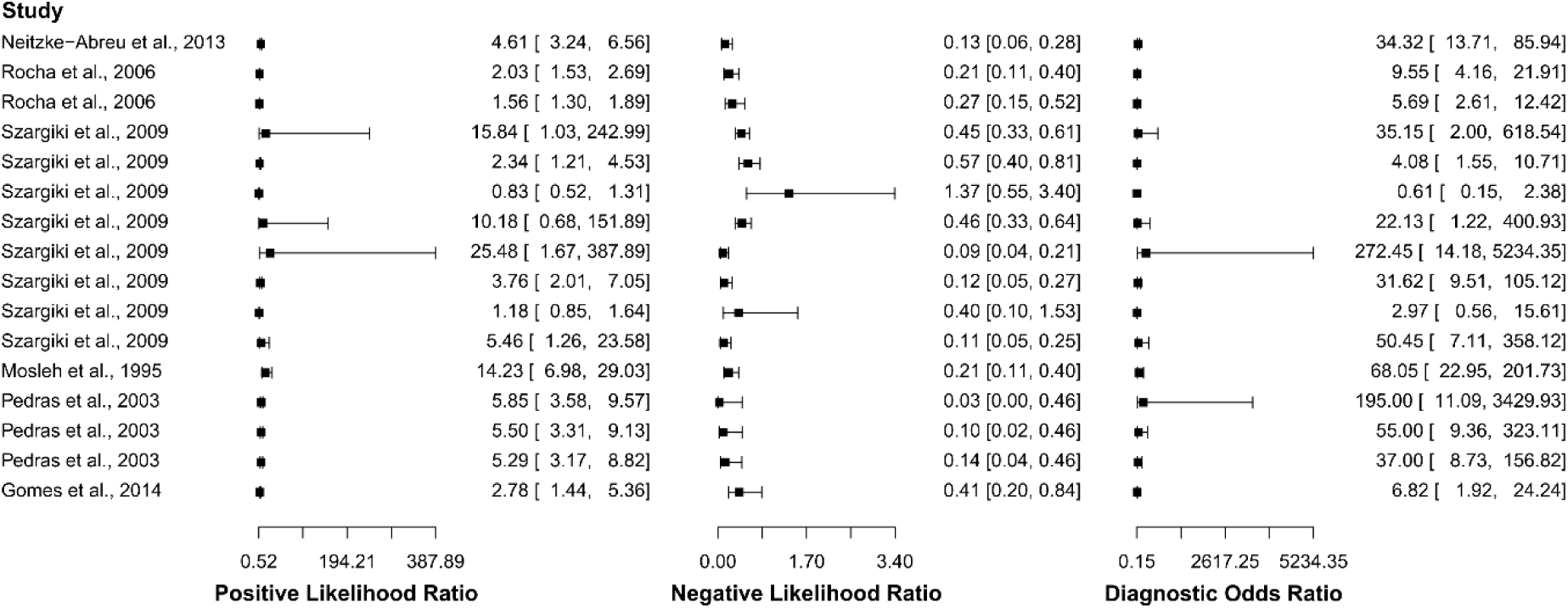
Study data and paired forest plot of the positive likelihood ratio, negative likelihood ratio, and diagnostic odds ratio of indirect fluorescent antibody test (IFAT) in the diagnosis of tegumentary leishmaniasis. The positive likelihood ratio, negative likelihood ratio, and diagnostic odds ratio are reported with a mean (95% confidence limits) for the included studies (44,47,51,58,68,69).

**S7 Fig.**
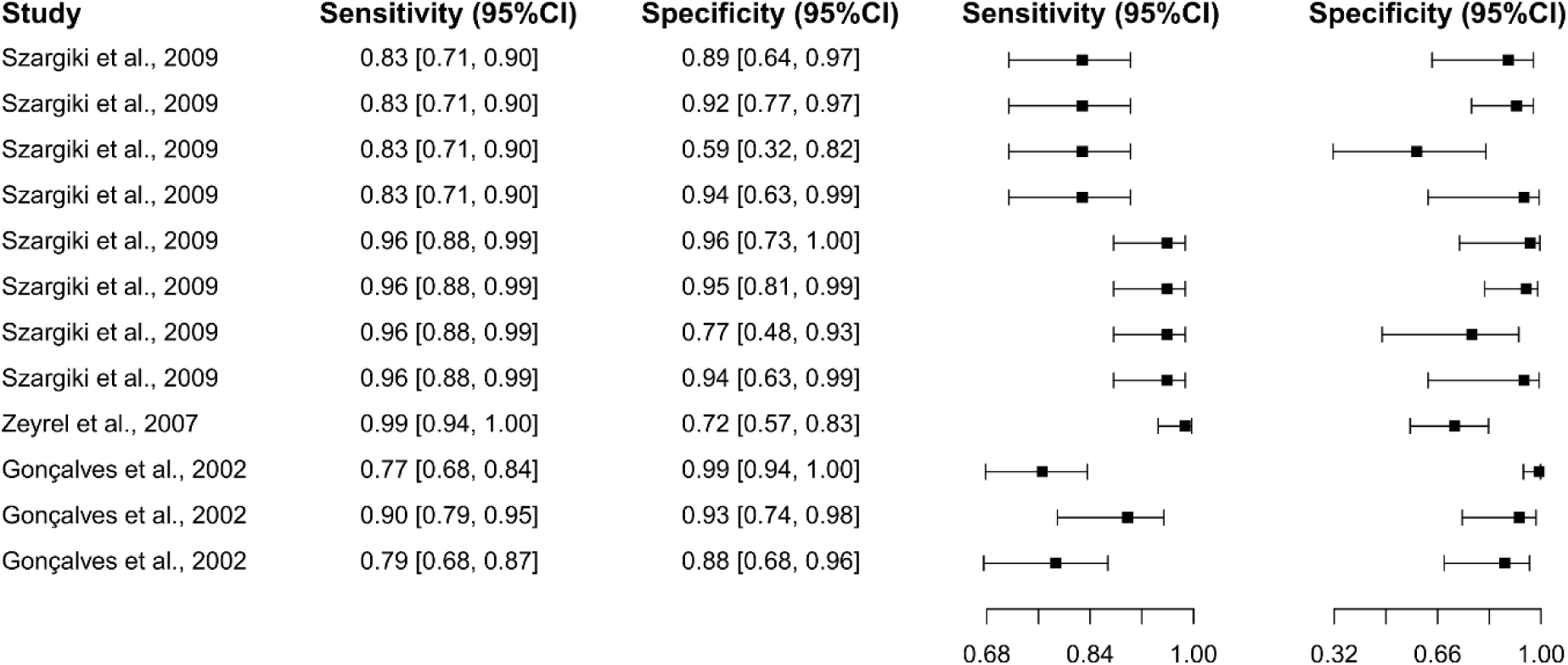
Study data and paired forest plot of the sensitivity and specificity of Western Blot (WB) in tegumentary leishmaniasis diagnosis. Data from each included study (51,59,70) are summarized. Sensitivity and specificity are reported with a mean (95% confidence limits). The forest plot depicts the estimated sensitivity and specificity (black squares) and their 95% confidence limits (horizontal black line).

**S8 Fig.**
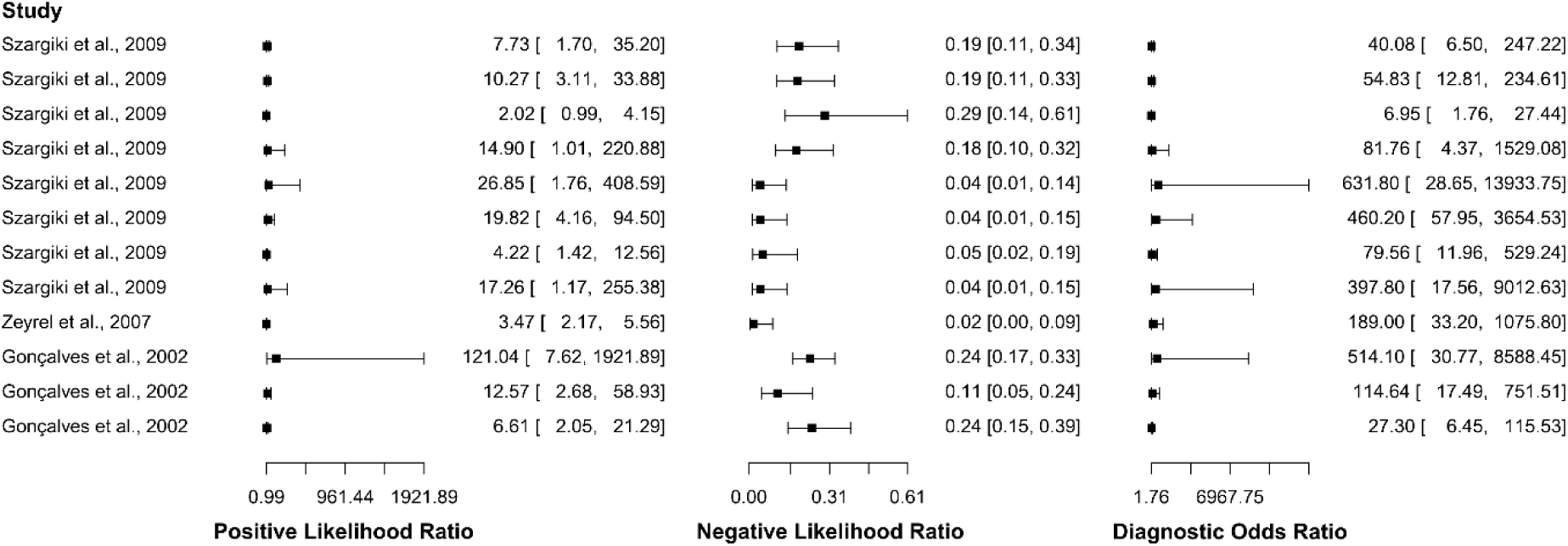
Study data and paired forest plot of the positive likelihood ratio, negative likelihood ratio, and diagnostic odds ratio of Western Blot (WB) in the diagnosis of tegumentary leishmaniasis. The positive likelihood ratio, negative likelihood ratio, and diagnostic odds ratio are reported with a mean (95% confidence limits) for the included studies (51,59,70).

**S9 Fig.**
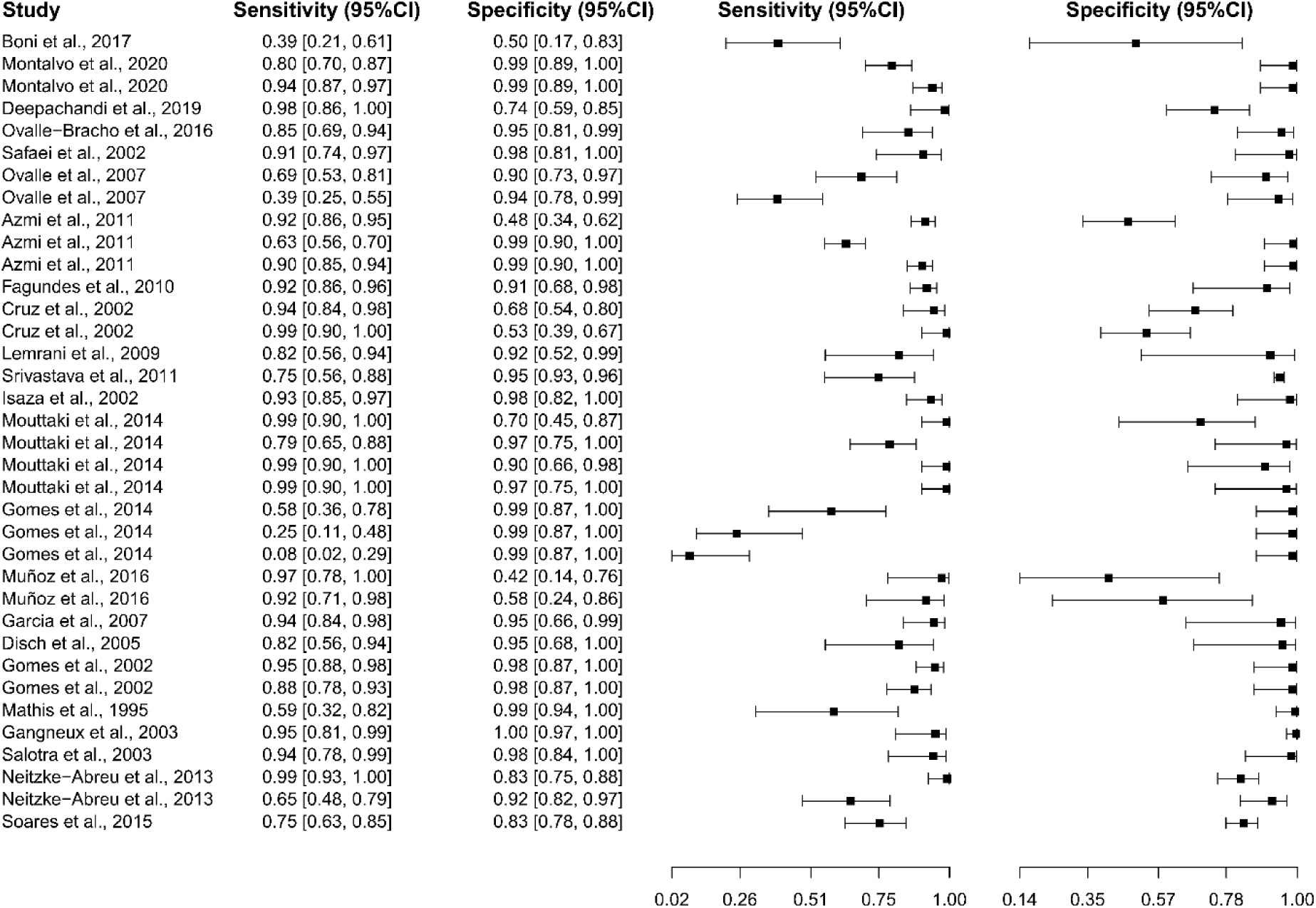
Study data and paired forest plot of the sensitivity and specificity of polymerase chain reaction (PCR) in tegumentary leishmaniasis diagnosis. Data from each included study (44–47,71–89) are summarized. Sensitivity and specificity are reported with a mean (95% confidence limits). The forest plot depicts the estimated sensitivity and specificity (black squares) and their 95% confidence limits (horizontal black line).

**S10 Fig.**
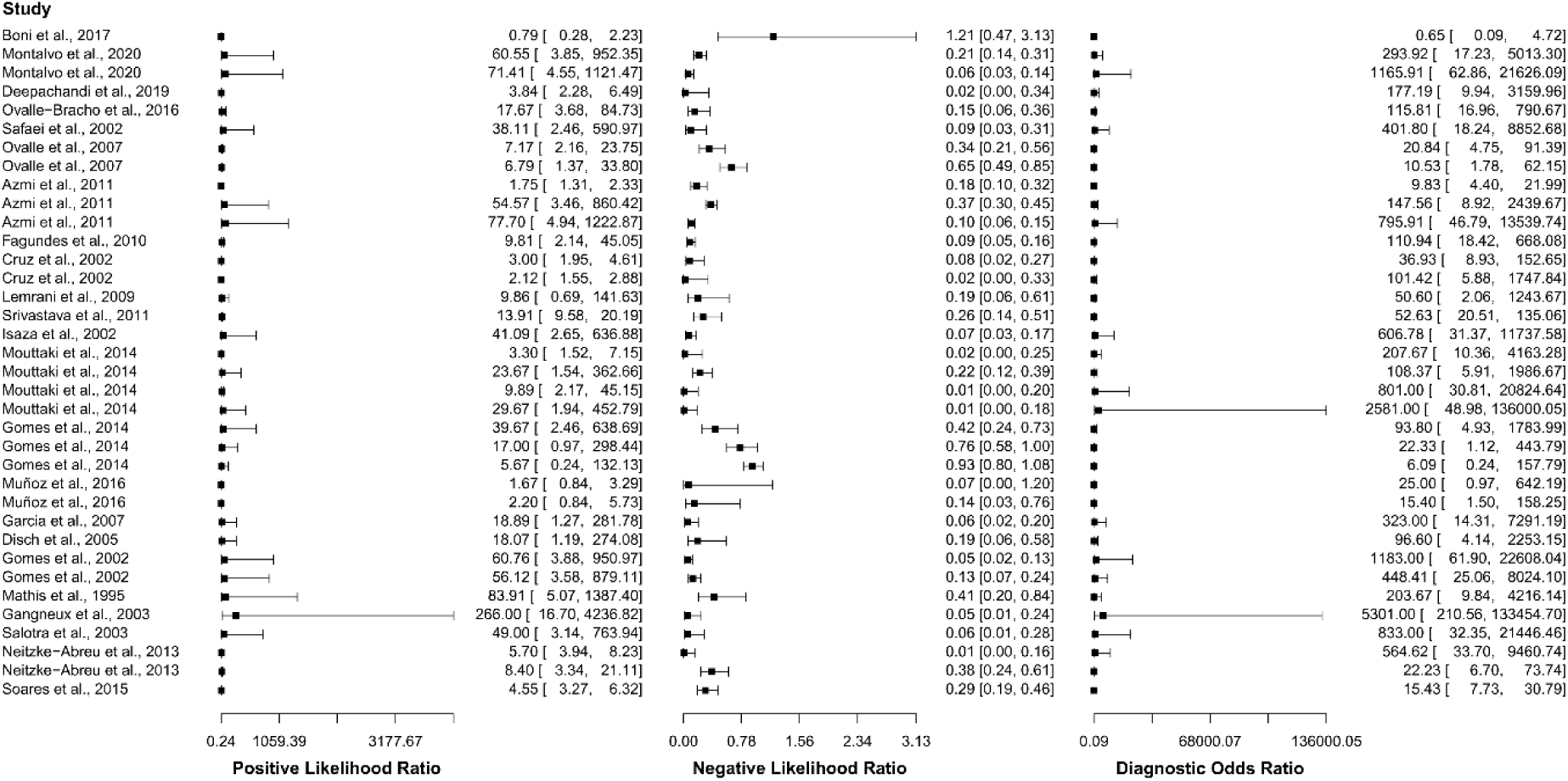
Study data and paired forest plot of the positive likelihood ratio, negative likelihood ratio, and diagnostic odds ratio of polymerase chain reaction (PCR) in the diagnosis of tegumentary leishmaniasis. The positive likelihood ratio, negative likelihood ratio, and diagnostic odds ratio are reported with a mean (95% confidence limits) for the included studies (44–47,71–89).

**S11 Fig.**
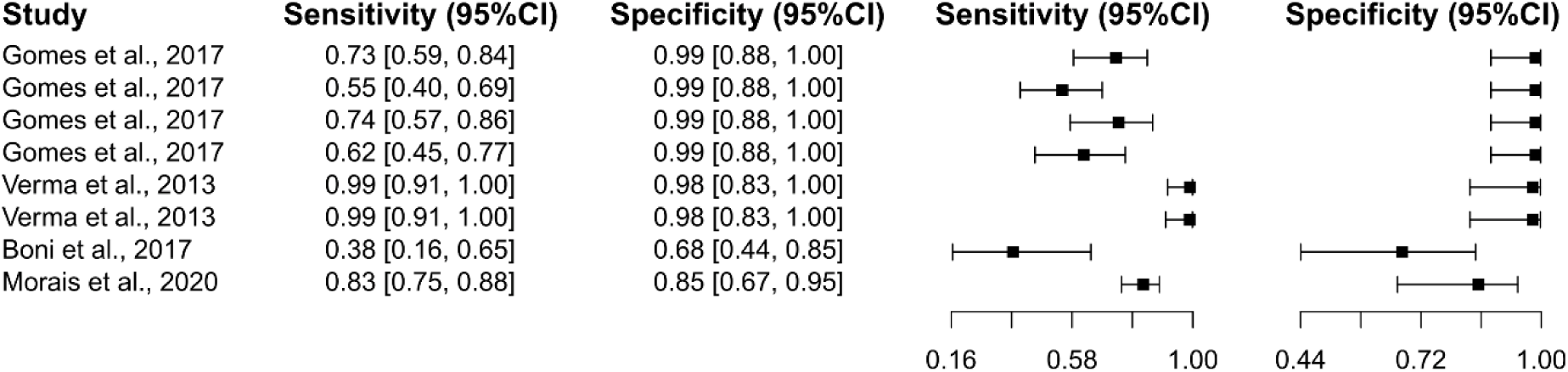
Study data and paired forest plot of the sensitivity and specificity of real-time polymerase chain reaction (qPCR) in tegumentary leishmaniasis diagnosis. Data from each included study (84,90–92) are summarized. Sensitivity and specificity are reported with a mean (95% confidence limits). The forest plot depicts the estimated sensitivity and specificity (black squares) and their 95% confidence limits (horizontal black line).

**S12 Fig.**
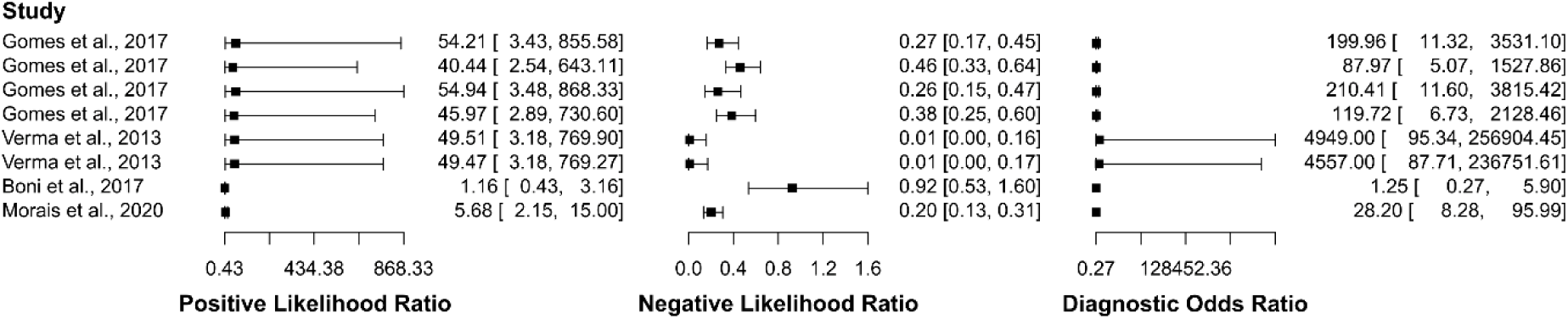
Study data and paired forest plot of the positive likelihood ratio, negative likelihood ratio, and diagnostic odds ratio of real-time polymerase chain reaction (qPCR) in the diagnosis of tegumentary leishmaniasis. The positive likelihood ratio, negative likelihood ratio, and diagnostic odds ratio are reported with a mean (95% confidence limits) for the included studies (84,90–92).

**S13 Fig.**
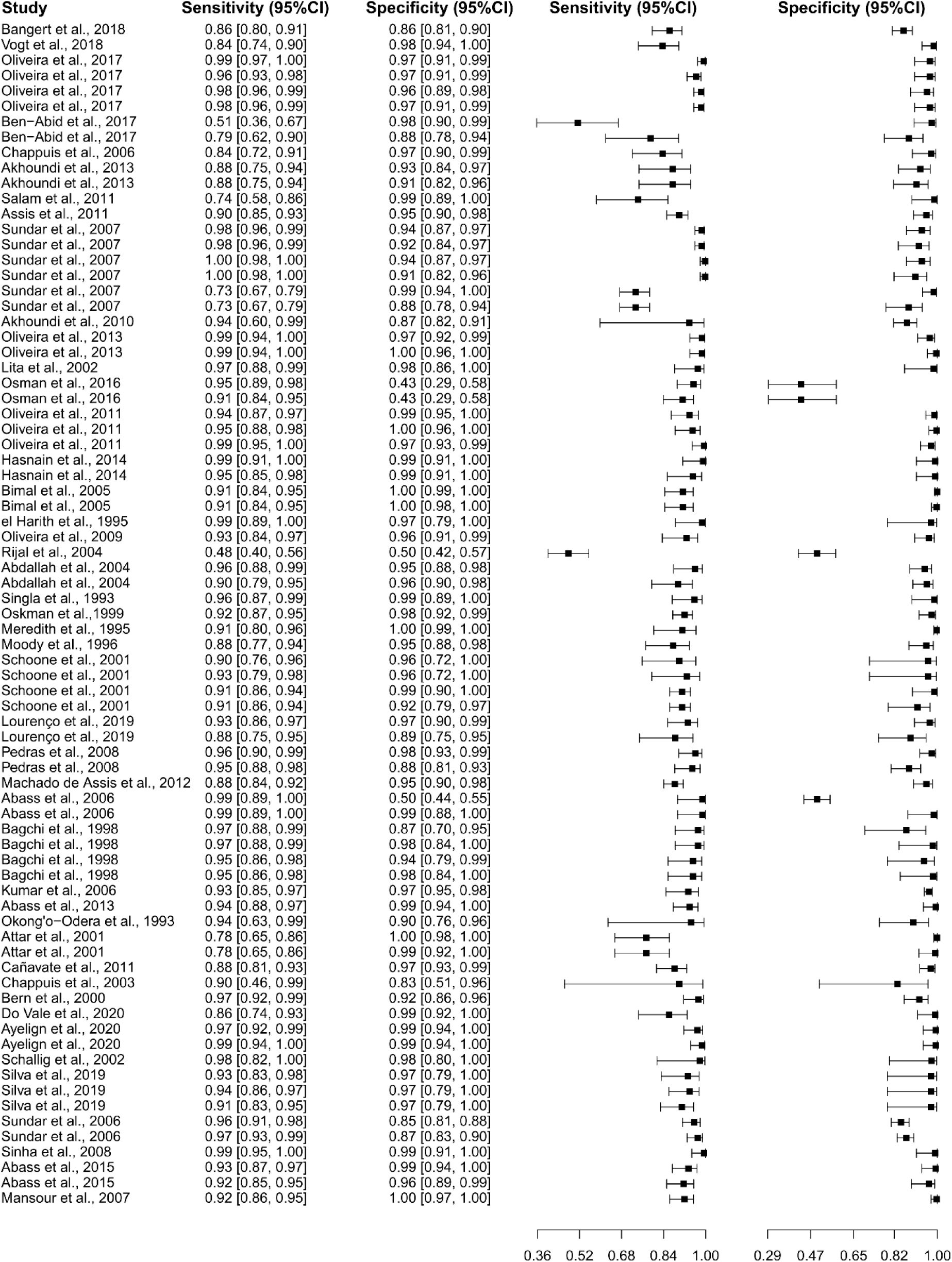
Study data and paired forest plot of the sensitivity and specificity of the direct agglutination test (DAT) in visceral leishmaniasis diagnosis. Data from each included study (50,96–139) are summarized. Sensitivity and specificity are reported with a mean (95% confidence limits). The forest plot depicts the estimated sensitivity and specificity (black squares) and their 95% confidence limits (horizontal black line).

**S14 Fig.**
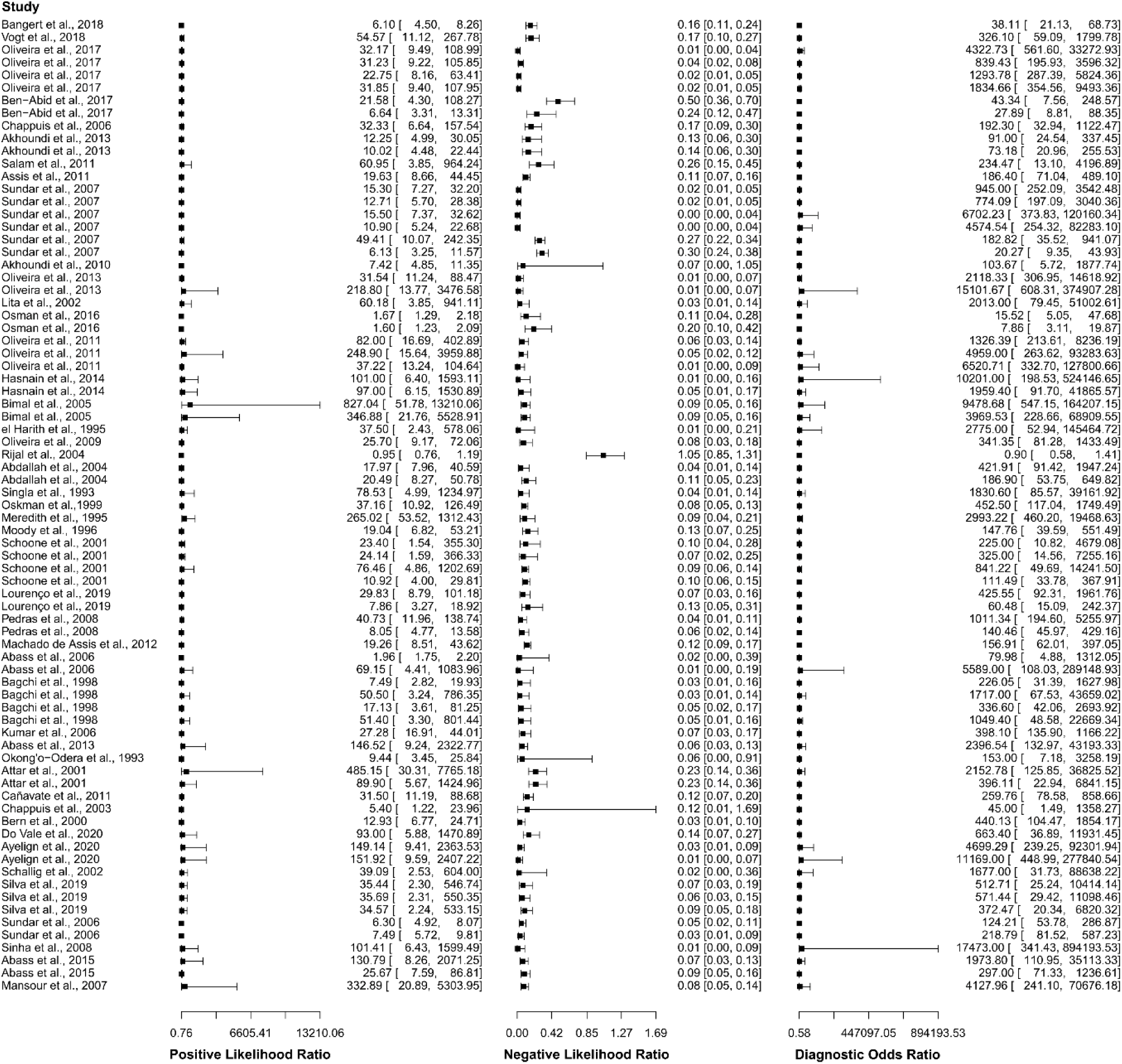
Study data and paired forest plot of the positive likelihood ratio, negative likelihood ratio, and diagnostic odds ratio of the direct agglutination test (DAT) in the diagnosis of visceral leishmaniasis. The positive likelihood ratio, negative likelihood ratio, and diagnostic odds ratio are reported with a mean (95% confidence limits) for the included studies (50,96–139).

**S15 Fig.**
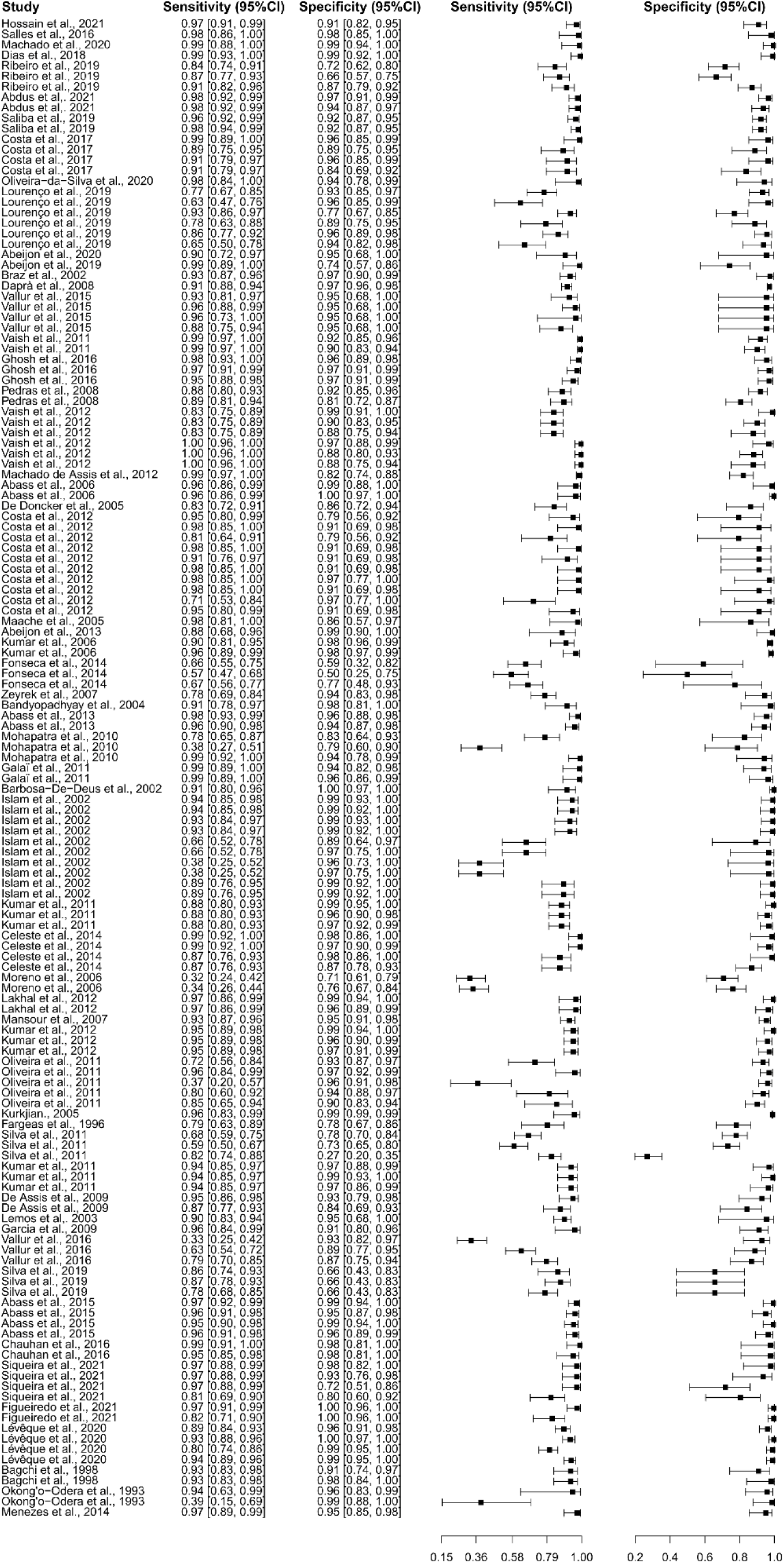
Study data and paired forest plot of the sensitivity and specificity of enzyme-linked immunosorbent assay (ELISA) in visceral leishmaniasis diagnosis. Data from each included study (50,59,67,114–117,119–121,130,133,134,140–184) are summarized. Sensitivity and specificity are reported with a mean (95% confidence limits). The forest plot depicts the estimated sensitivity and specificity (black squares) and their 95% confidence limits (horizontal black line).

**S16 Fig.**
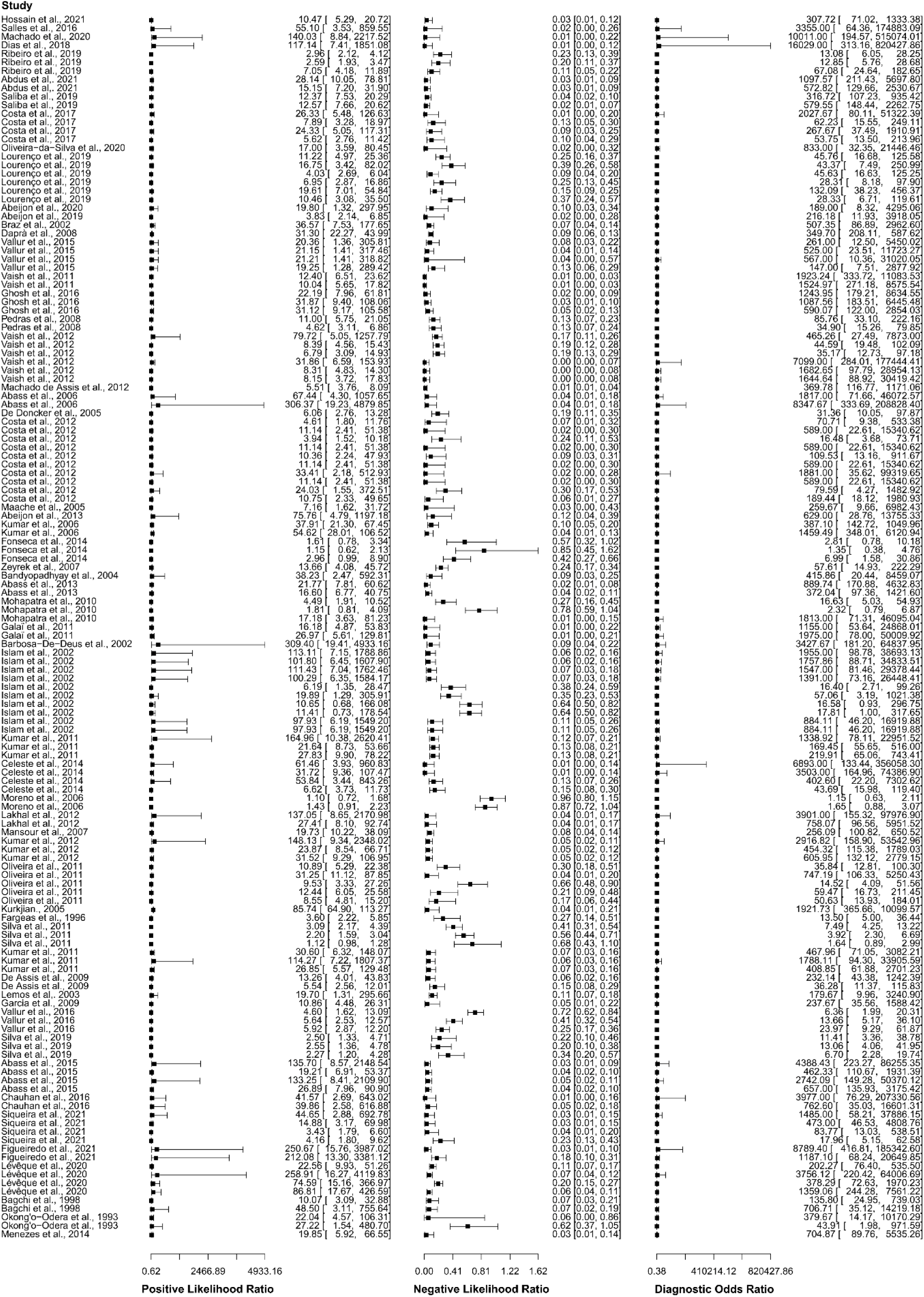
Study data and paired forest plot of the positive likelihood ratio, negative likelihood ratio, and diagnostic odds ratio of enzyme-linked immunosorbent assay (ELISA) in the diagnosis of visceral leishmaniasis. The positive likelihood ratio, negative likelihood ratio, and diagnostic odds ratio are reported with a mean (95% confidence limits) for the included studies (50,59,67,114–117,119–121,130,133,134,140–184).

**S17 Fig.**
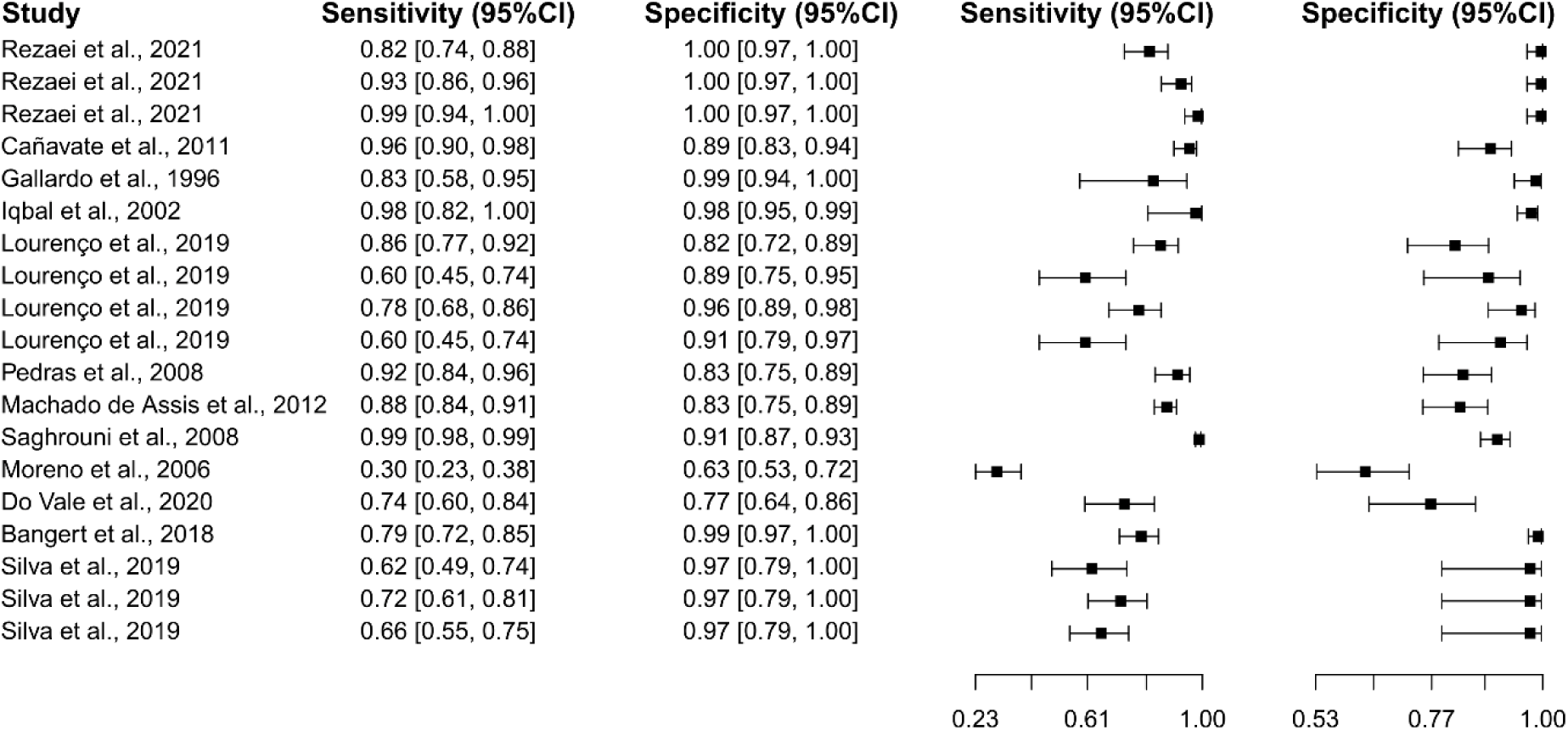
Study data and paired forest plot of the sensitivity and specificity of indirect fluorescent antibody test (IFAT) in visceral leishmaniasis diagnosis. Data from each included study (50,96,114,115,123,126,130,169,185–188) are summarized. Sensitivity and specificity are reported with a mean (95% confidence limits). The forest plot depicts the estimated sensitivity and specificity (black squares) and their 95% confidence limits (horizontal black line).

**S18 Fig.**
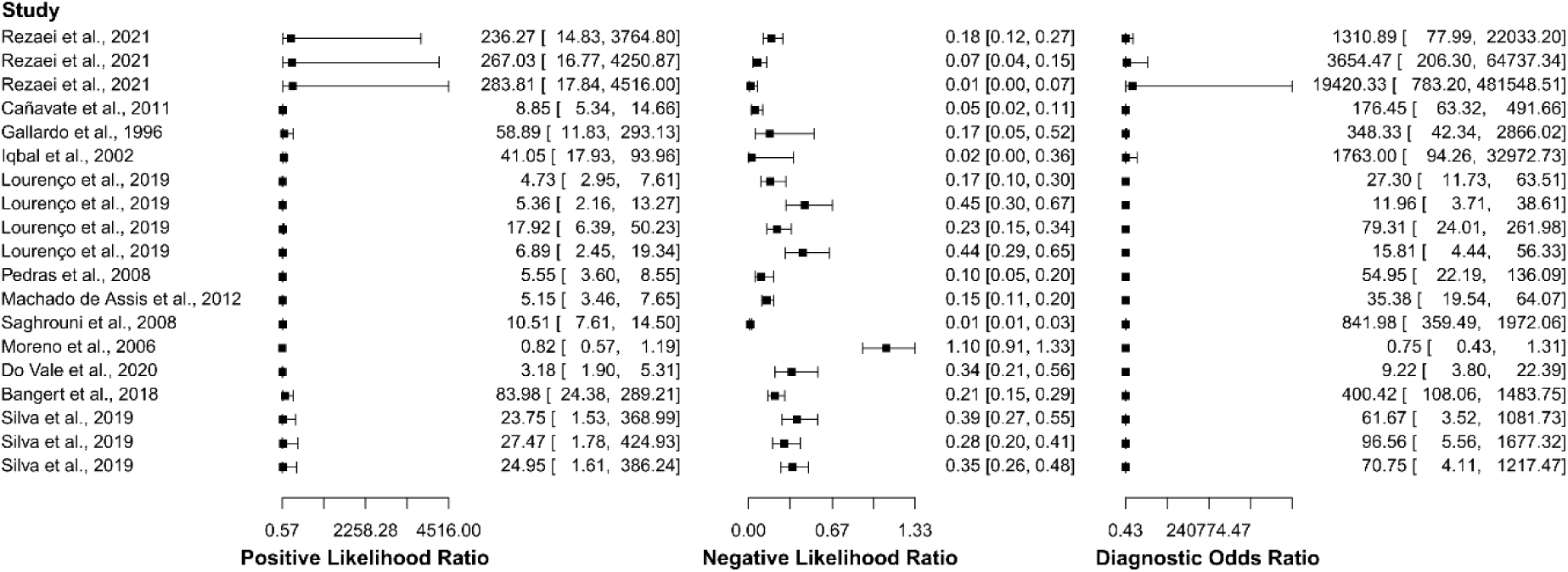
Study data and paired forest plots of the positive likelihood ratio, negative likelihood ratio, and diagnostic odds ratio of indirect fluorescent antibody test (IFAT) in the diagnosis of visceral leishmaniasis. The positive likelihood ratio, negative likelihood ratio, and diagnostic odds ratio are reported with a mean (95% confidence limits) for the included studies (50,96,114,115,123,126,130,169,185–188).

**S19 Fig.**
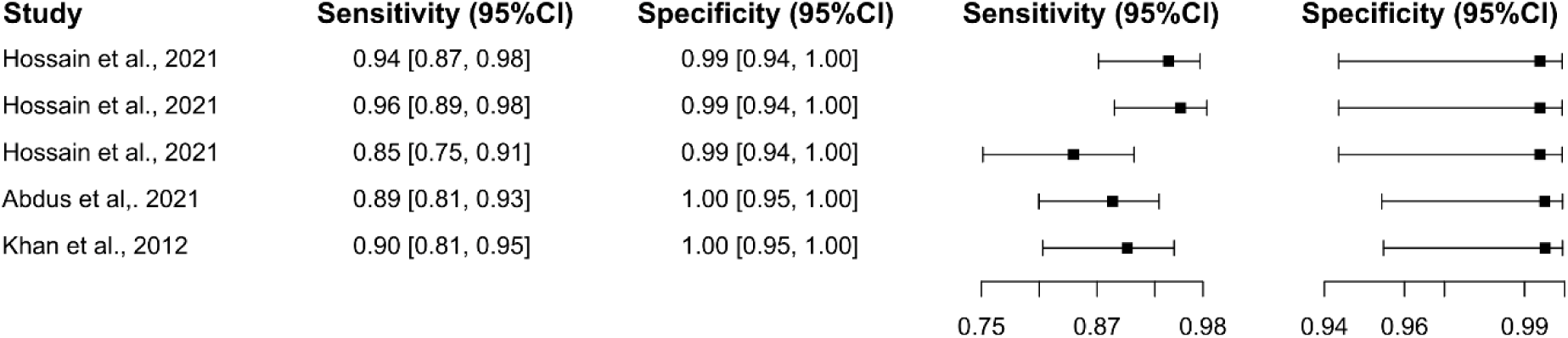
Study data and paired forest plot of the sensitivity and specificity of loop-mediated isothermal amplification (LAMP) in visceral leishmaniasis diagnosis. Data from each included study (140,191,194) are summarized. Sensitivity and specificity are reported with a mean (95% confidence limits). The forest plot depicts the estimated sensitivity and specificity (black squares) and their 95% confidence limits (horizontal black line).

**S20 Fig.**
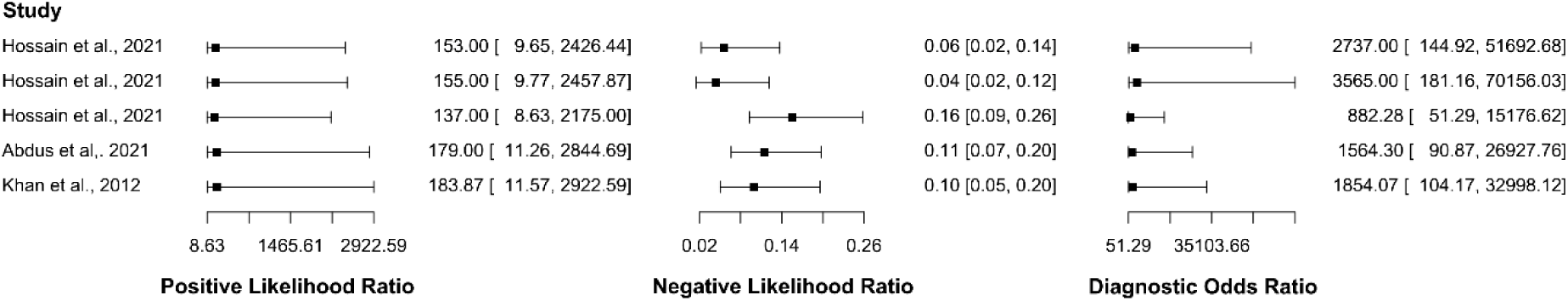
Study data and paired forest plot of the positive likelihood ratio, negative likelihood ratio, and diagnostic odds ratio of loop-mediated isothermal amplification (LAMP) in the diagnosis of visceral leishmaniasis. The positive likelihood ratio, negative likelihood ratio, and diagnostic odds ratio are reported with a mean (95% confidence limits) for the included studies (140,191,194).

**S21 Fig.**
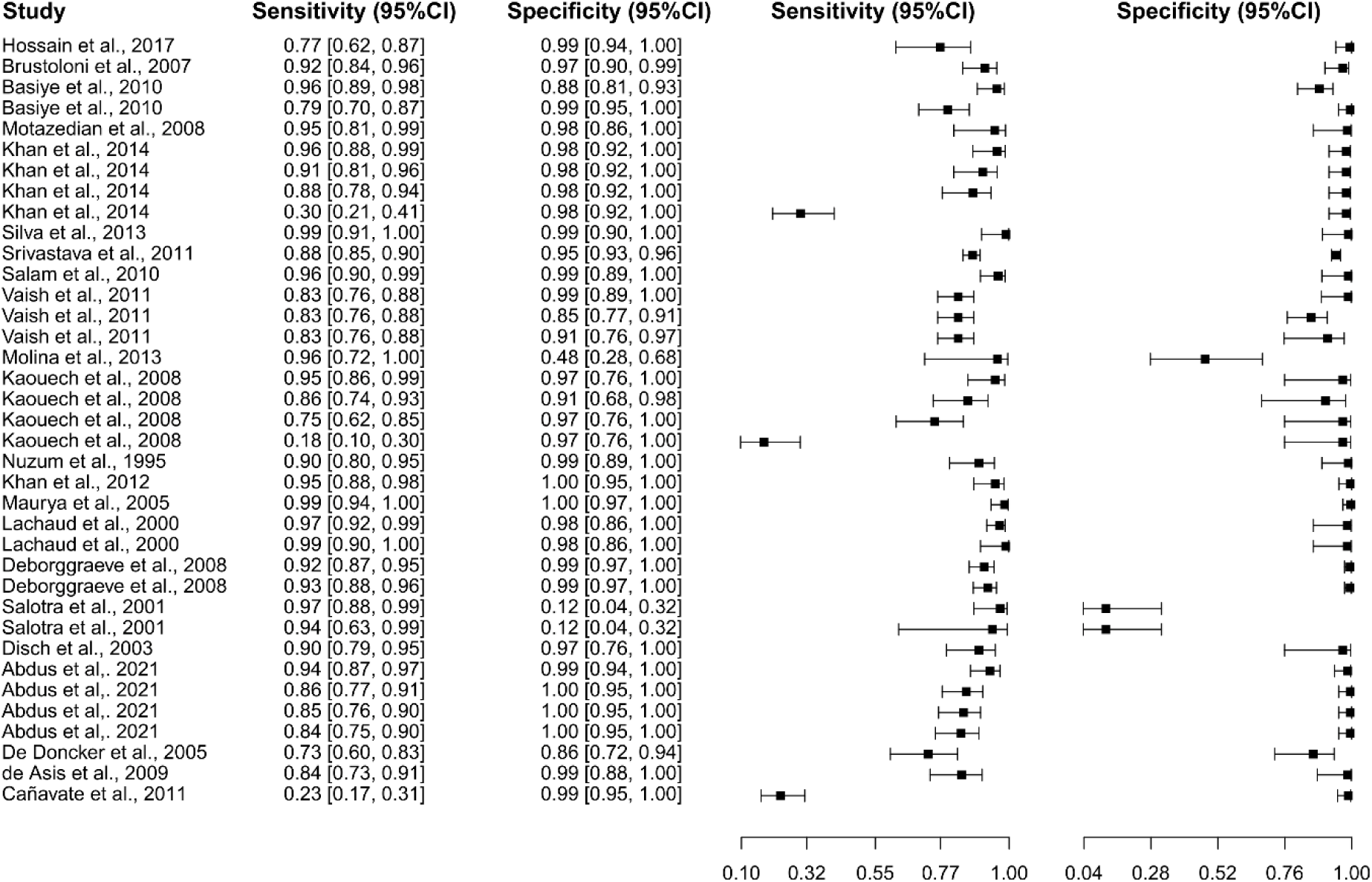
Study data and paired forest plot of the sensitivity and specificity of polymerase chain reaction (PCR) in visceral leishmaniasis diagnosis. Data from each included study (75,83,123,157,177,191,194–209) are summarized. Sensitivity and specificity are reported with a mean (95% confidence limits). The forest plot depicts the estimated sensitivity and specificity (black squares) and their 95% confidence limits (horizontal black line).

**S22 Fig.**
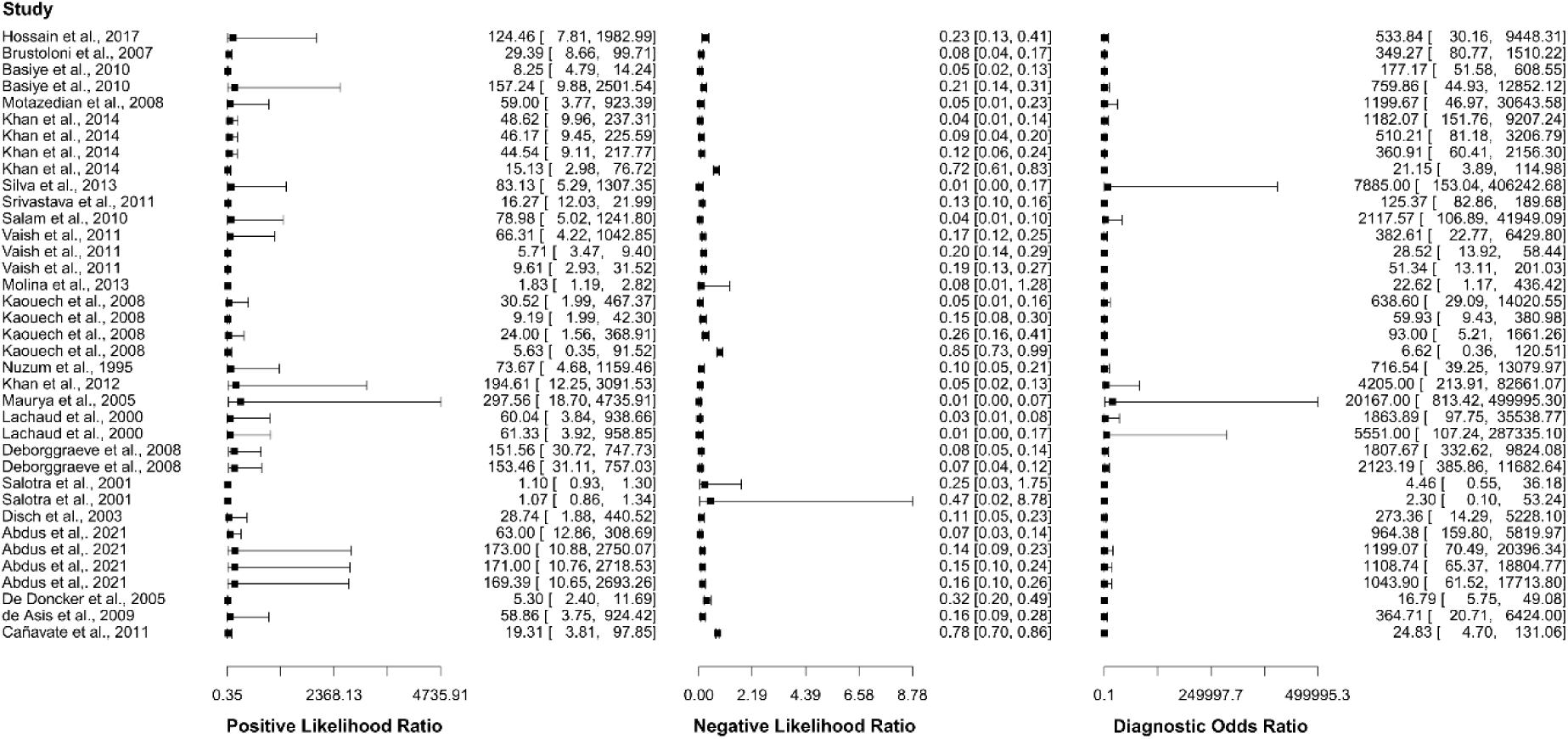
Study data and paired forest plot of the positive likelihood ratio, negative likelihood ratio, and diagnostic odds ratio of polymerase chain reaction (PCR) in the diagnosis of visceral leishmaniasis. The positive likelihood ratio, negative likelihood ratio, and diagnostic odds ratio are reported with a mean (95% confidence limits) for the included studies (75,83,123,157,177,191,194–209).

**S23 Fig.**
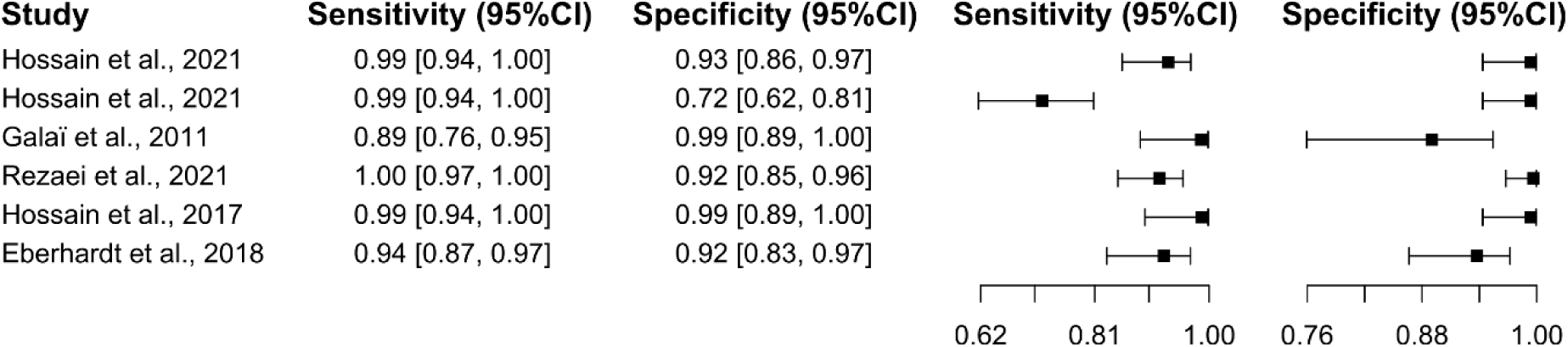
Study data and paired forest plot of the sensitivity and specificity of real-time polymerase chain reaction (qPCR) in visceral leishmaniasis diagnosis. Data from each included study (140,164,187,208,210) are summarized. Sensitivity and specificity are reported with a mean (95% confidence limits). The forest plot depicts the estimated sensitivity and specificity (black squares) and their 95% confidence limits (horizontal black line).

**S24 Fig.**
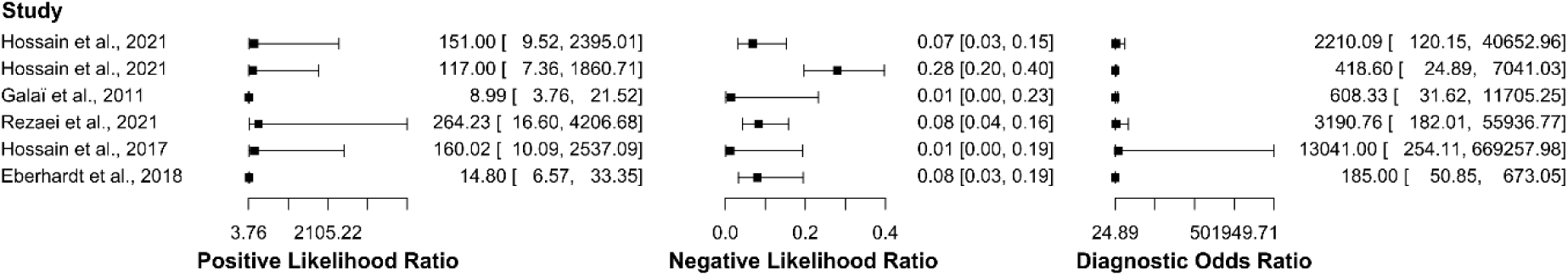
Study data and paired forest plot of the positive likelihood ratio, negative likelihood ratio, and diagnostic odds ratio of real-time polymerase chain reaction (qPCR) in the diagnosis of visceral leishmaniasis. The positive likelihood ratio, negative likelihood ratio, and diagnostic odds ratio are reported with a mean (95% confidence limits) for the included studies (140,164,187,208,210).

**S25 Fig.**
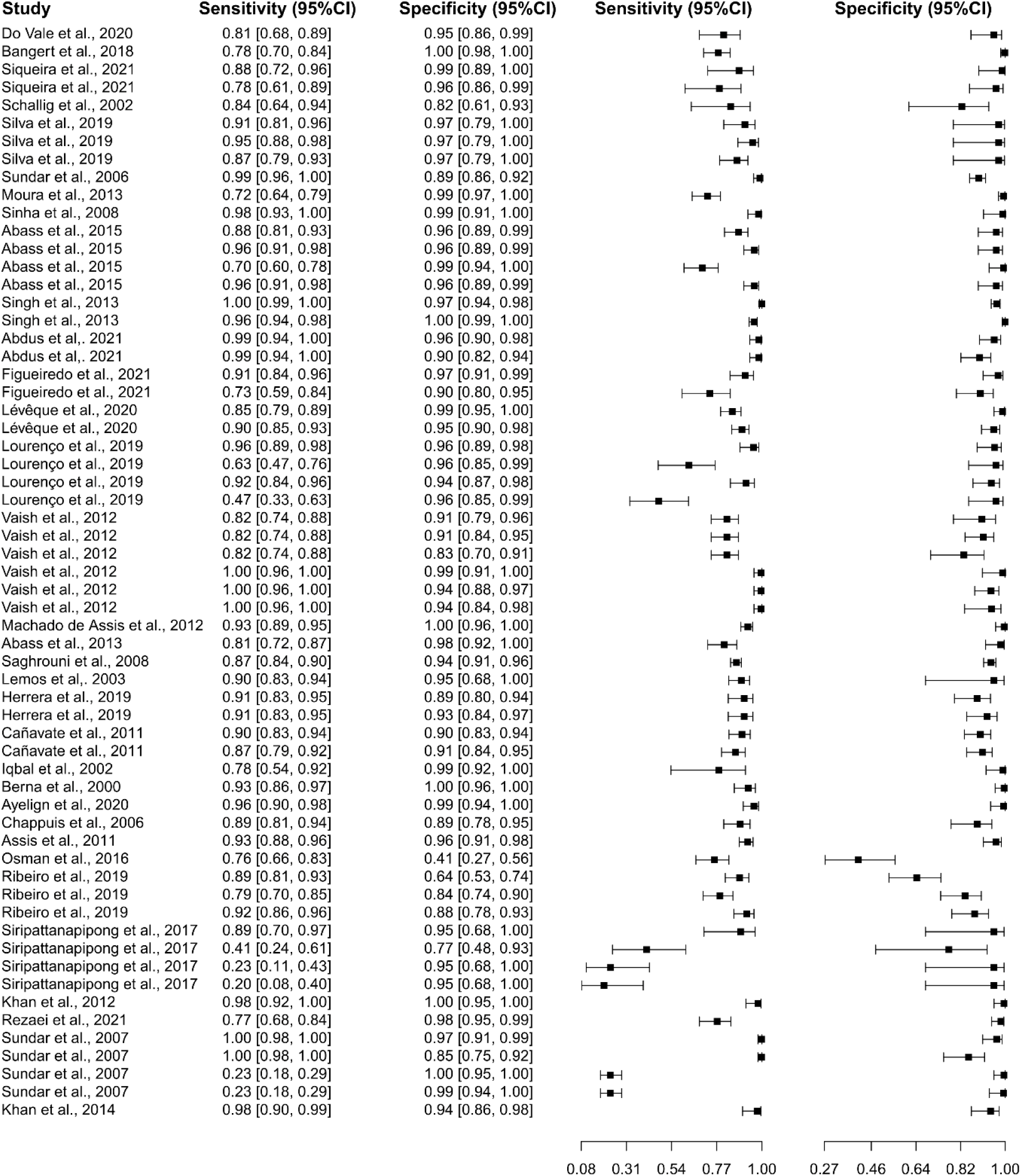
Study data and paired forest plots of the sensitivity and specificity of rapid diagnostic tests (RDT) in visceral leishmaniasis diagnosis. Data from each included study (96,100,114,115,120,123,125–128,130–133,135,138,139,144,156,178,182–187,189–195) are summarized. Sensitivity and specificity are reported with a mean (95% confidence limits). The forest plot depicts the estimated sensitivity and specificity (black squares) and their 95% confidence limits (horizontal black line).

**S26 Fig.**
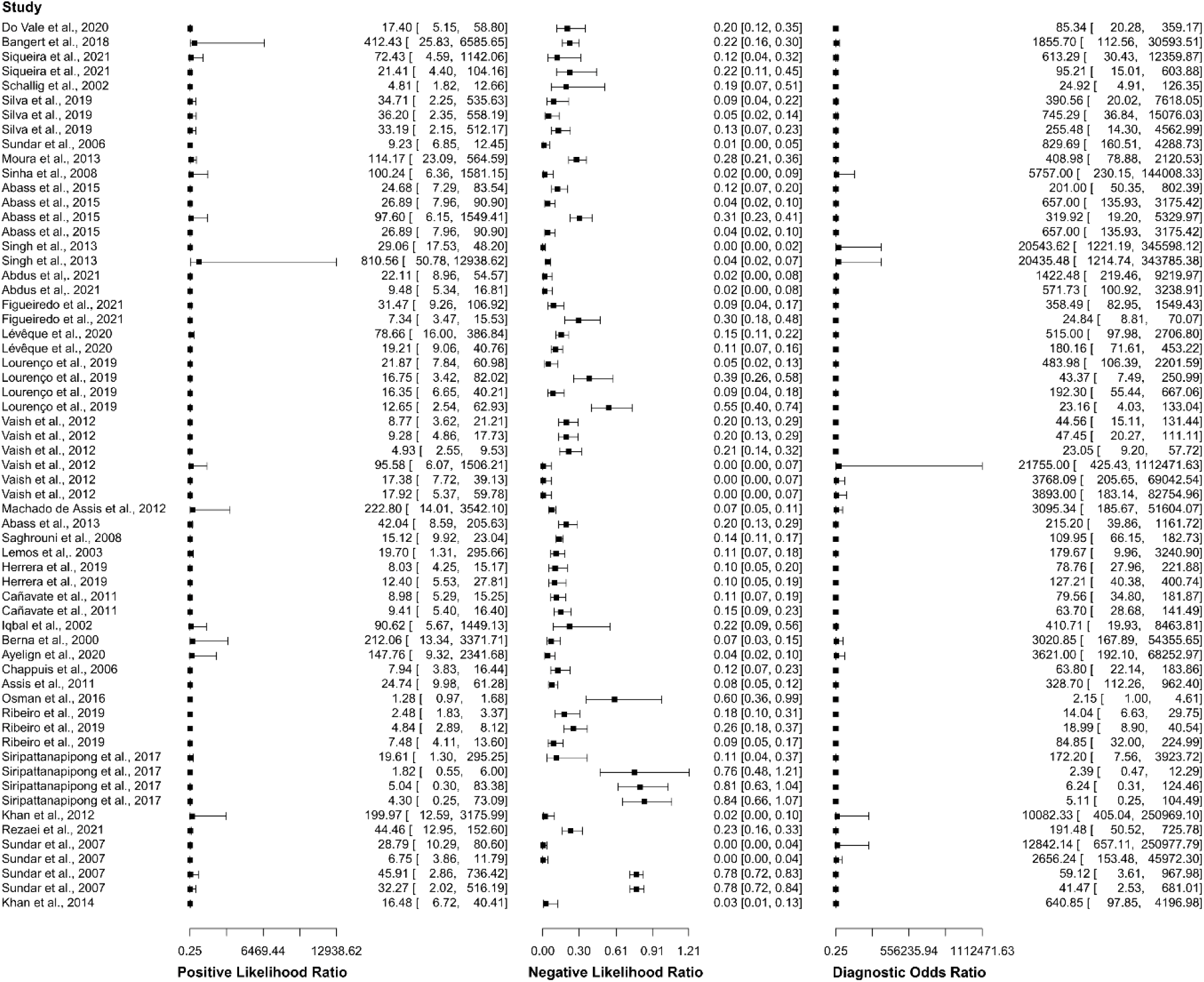
Study data and paired forest plots of the positive likelihood ratio, negative likelihood ratio, and diagnostic odds ratio of rapid diagnostic tests (RDT) in the diagnosis of visceral leishmaniasis. The positive likelihood ratio, negative likelihood ratio, and diagnostic odds ratio are reported with a mean (95% confidence limits) for the included studies (96,100,114,115,120,123,125–128,130–133,135,138,139,144,156,178,182–187,189–195).

**S1 Table.**
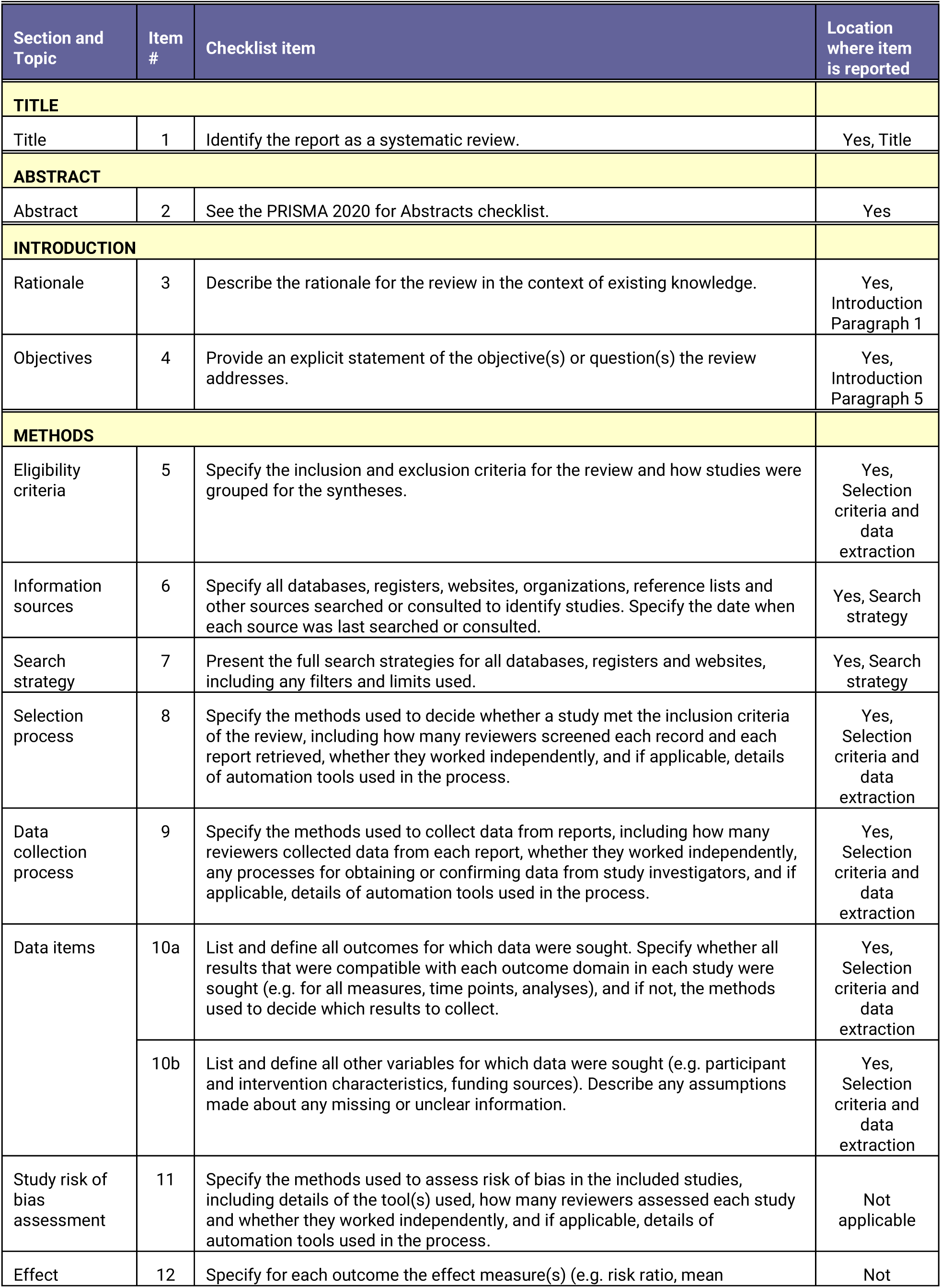
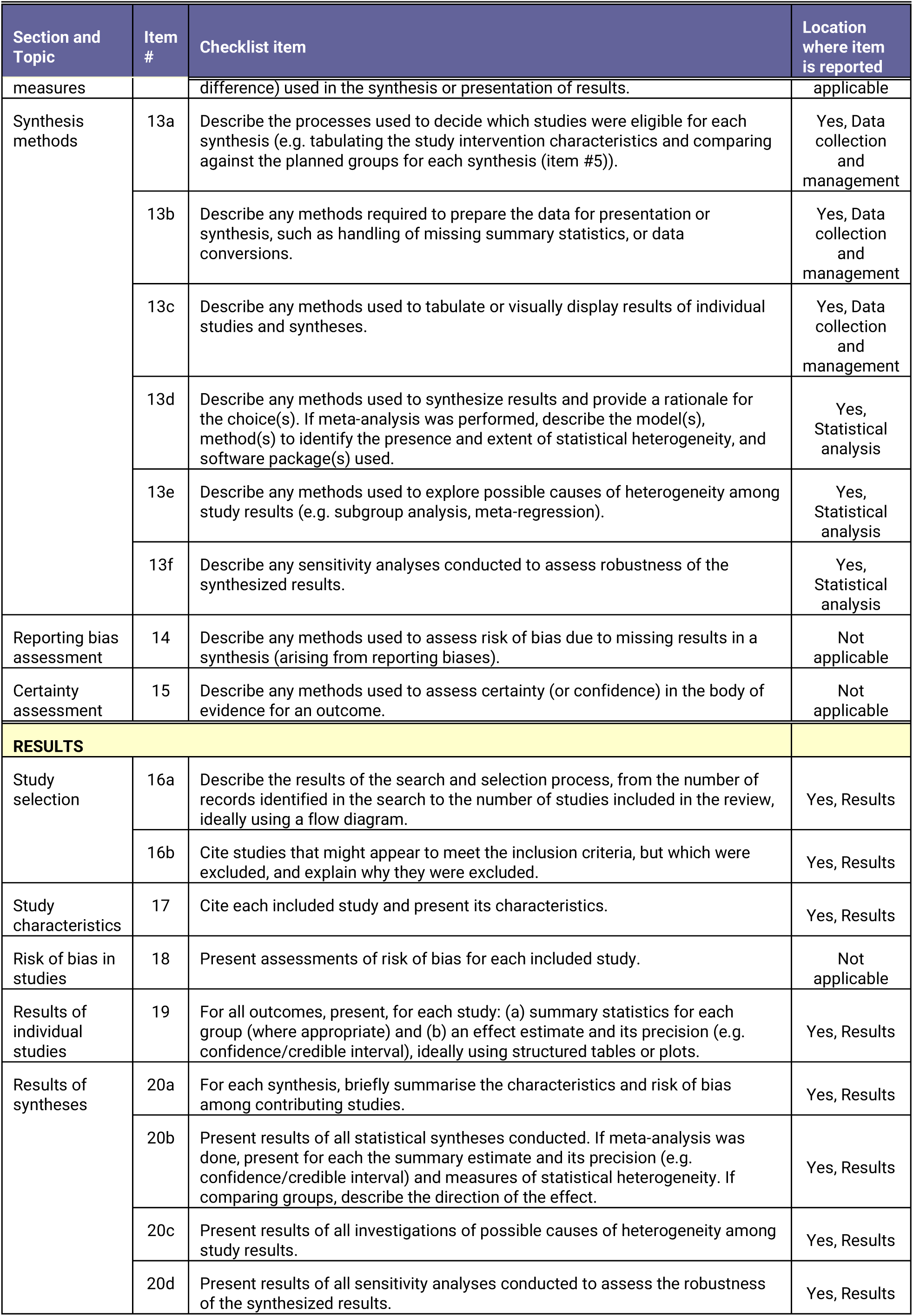
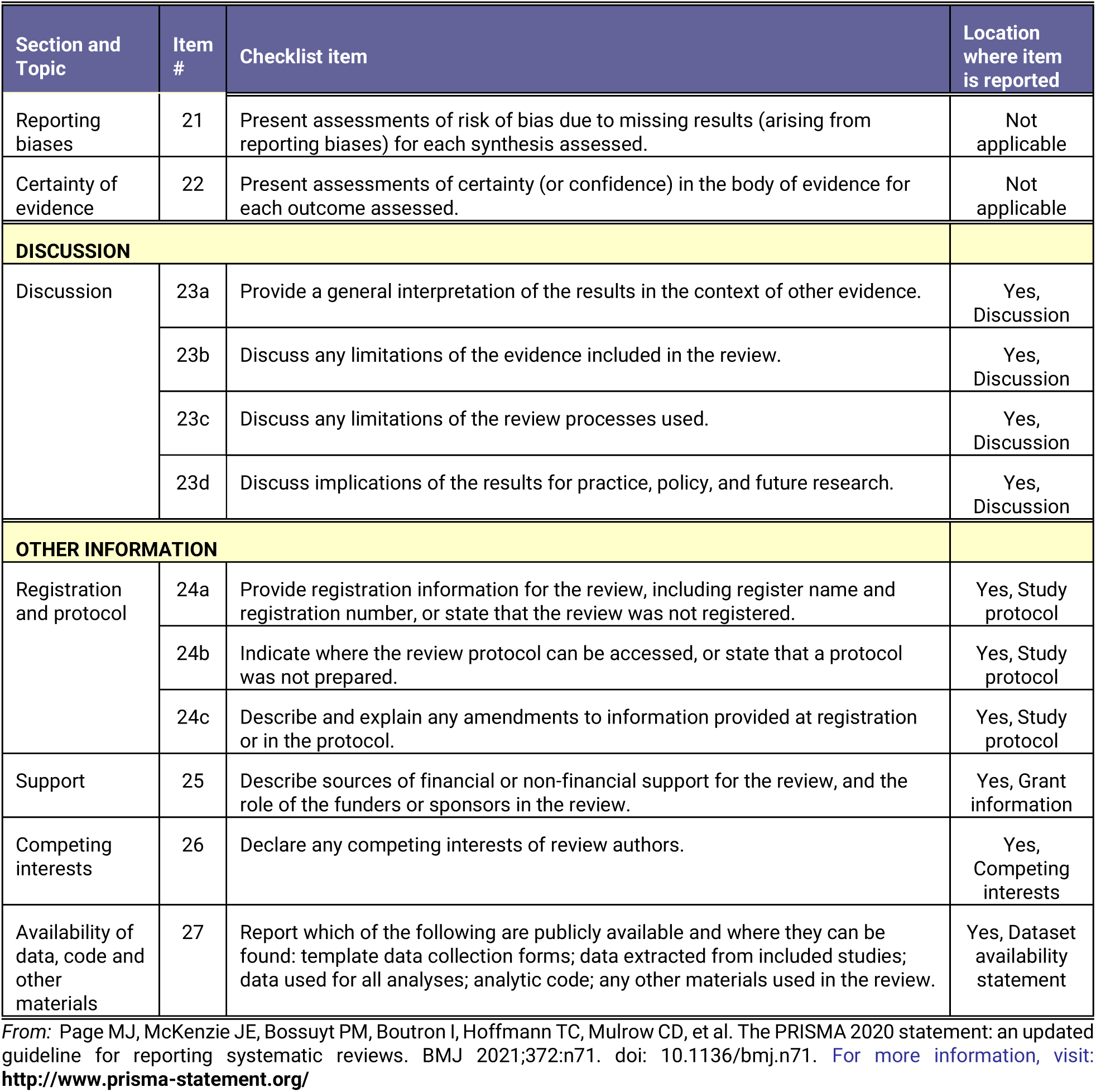
PRISMA checklist. PRISMA, Preferred Reporting Items for Systematic Reviews and Meta-analyses.

**S2 Table.**
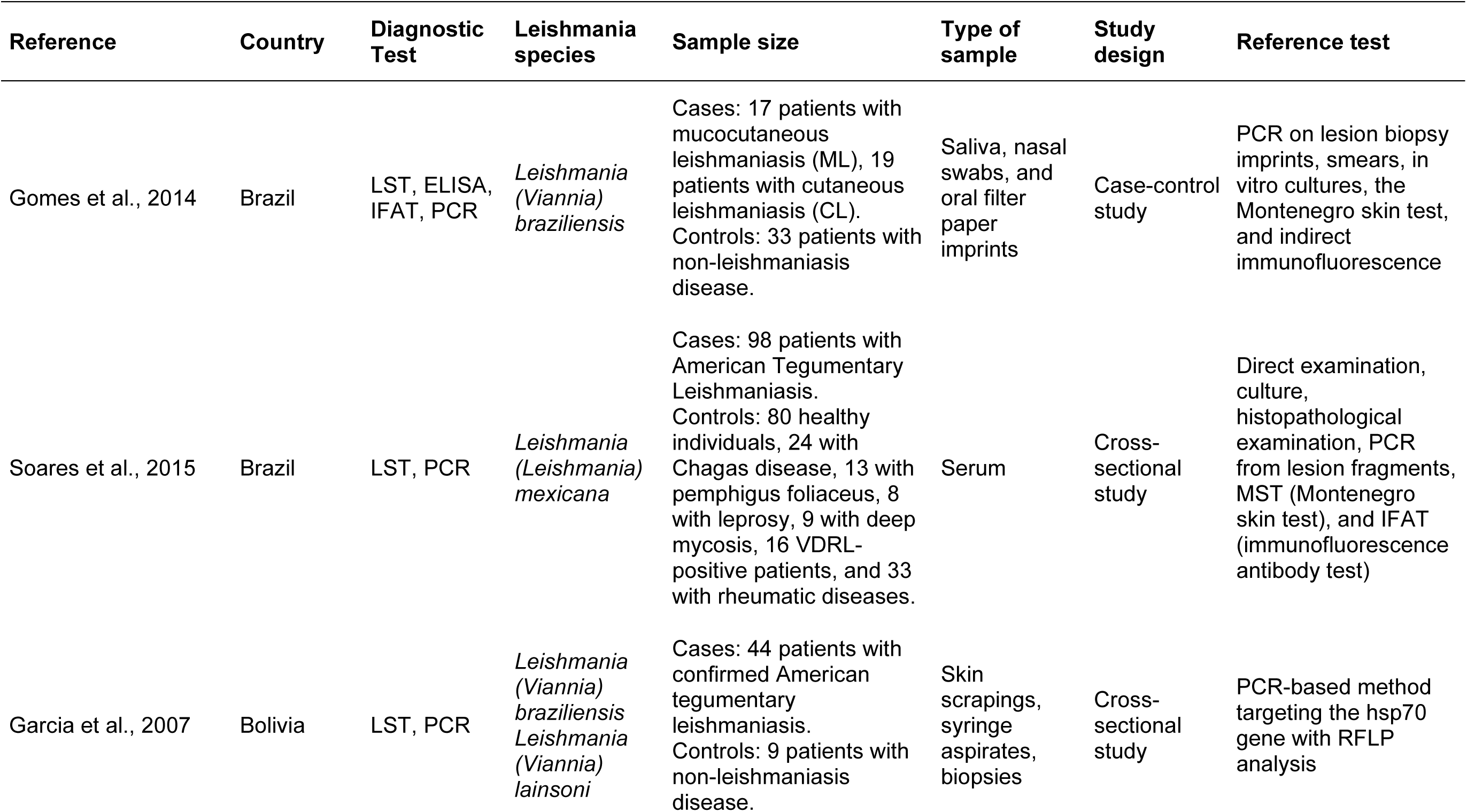
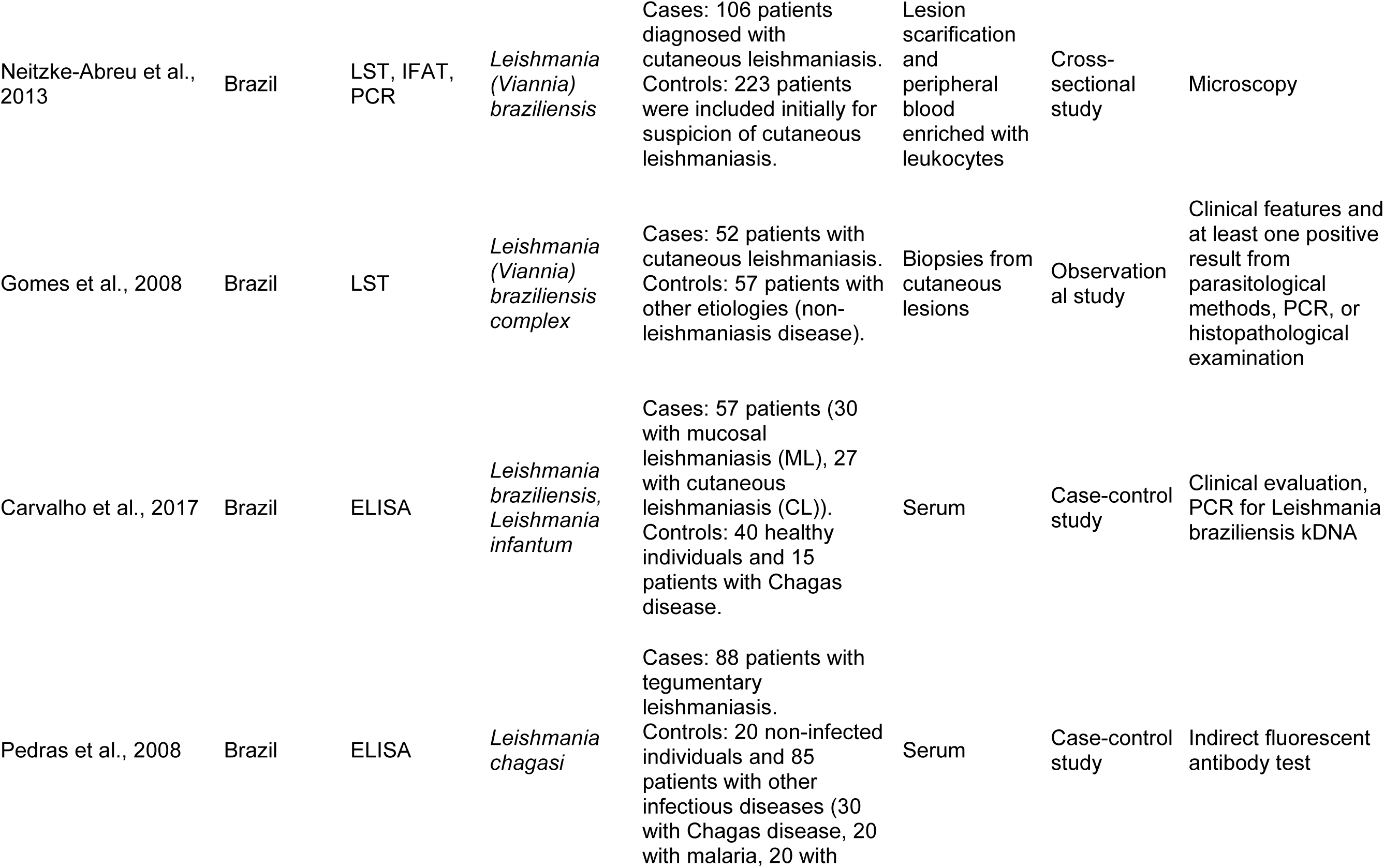
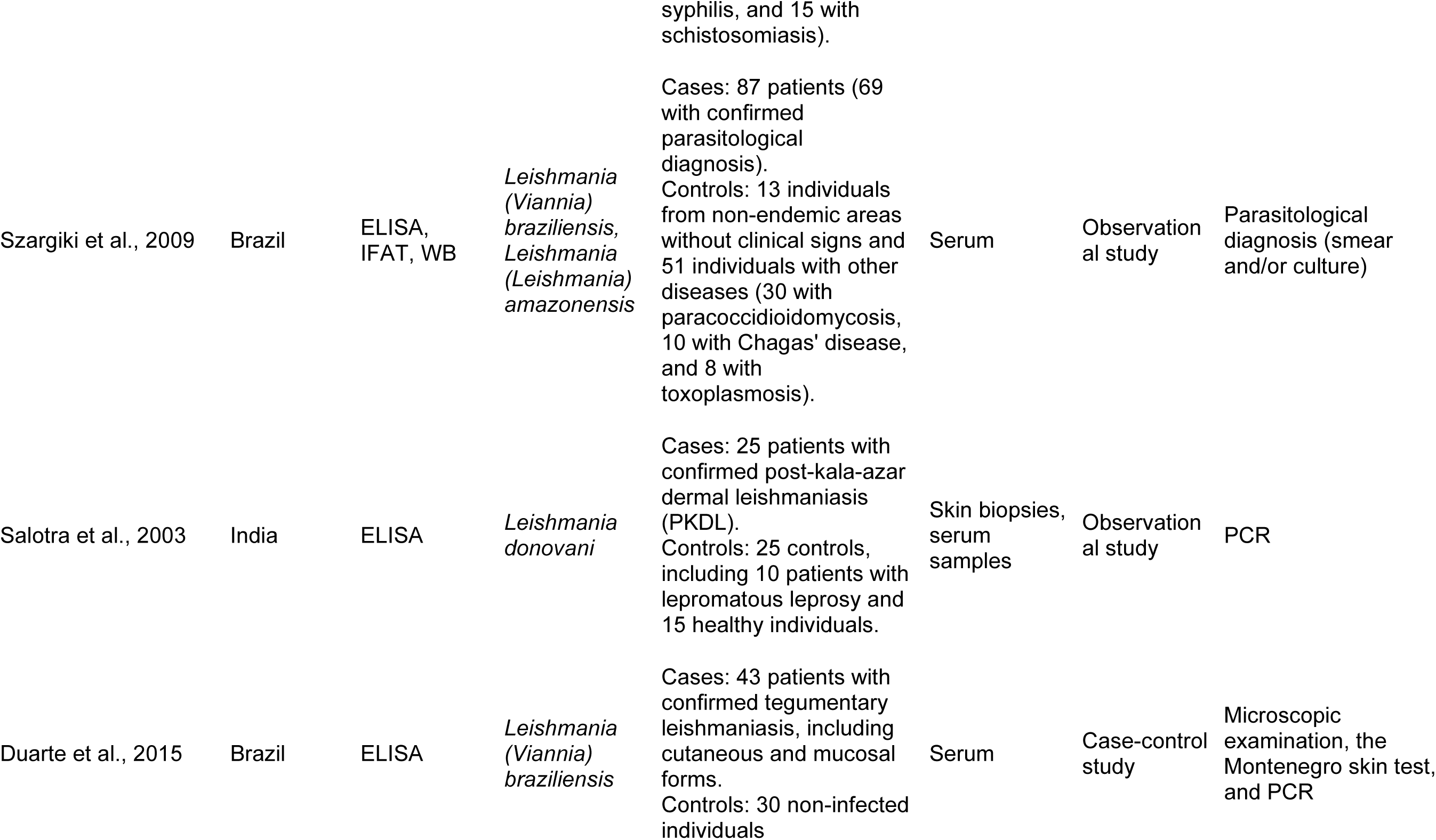
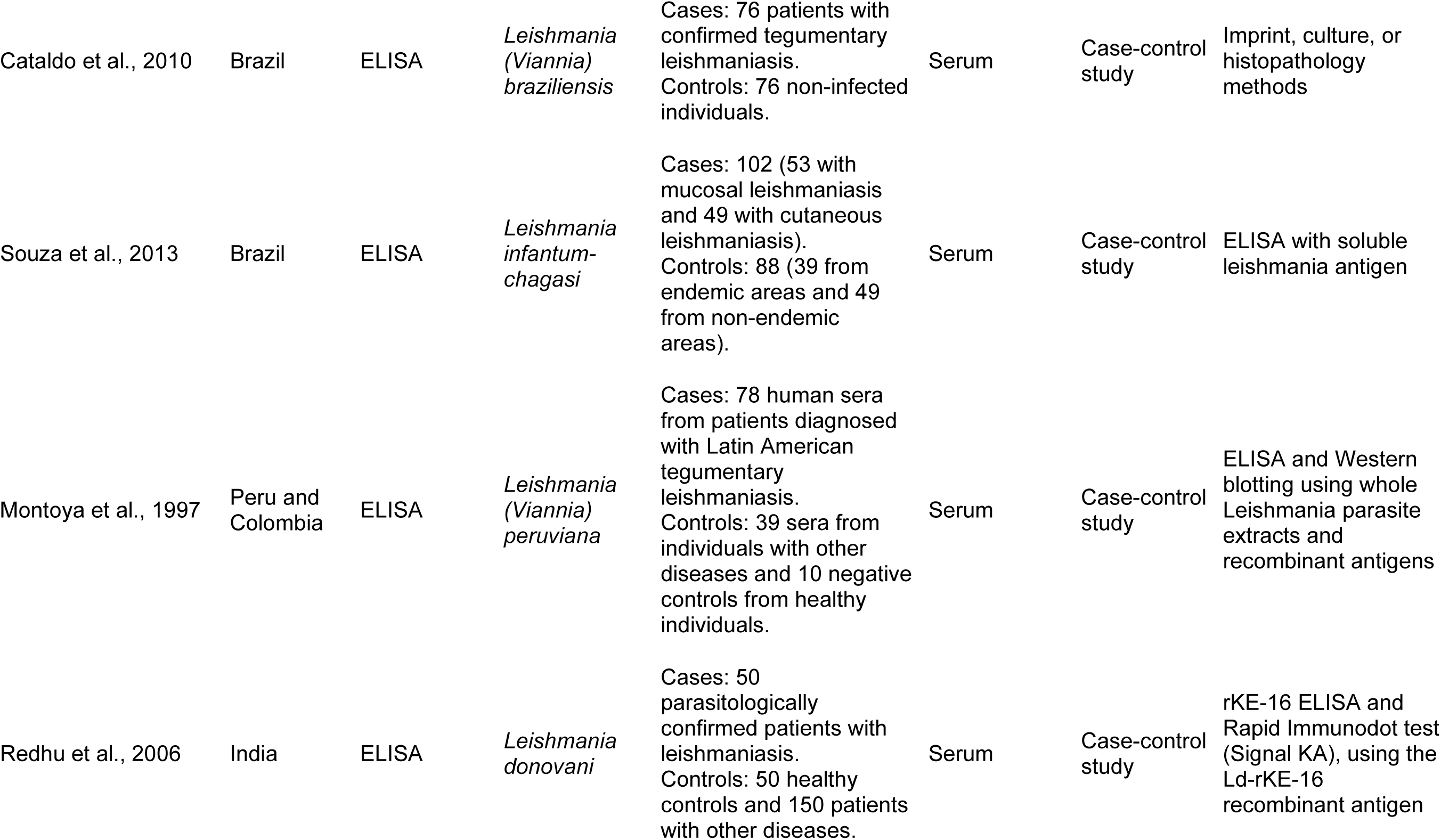
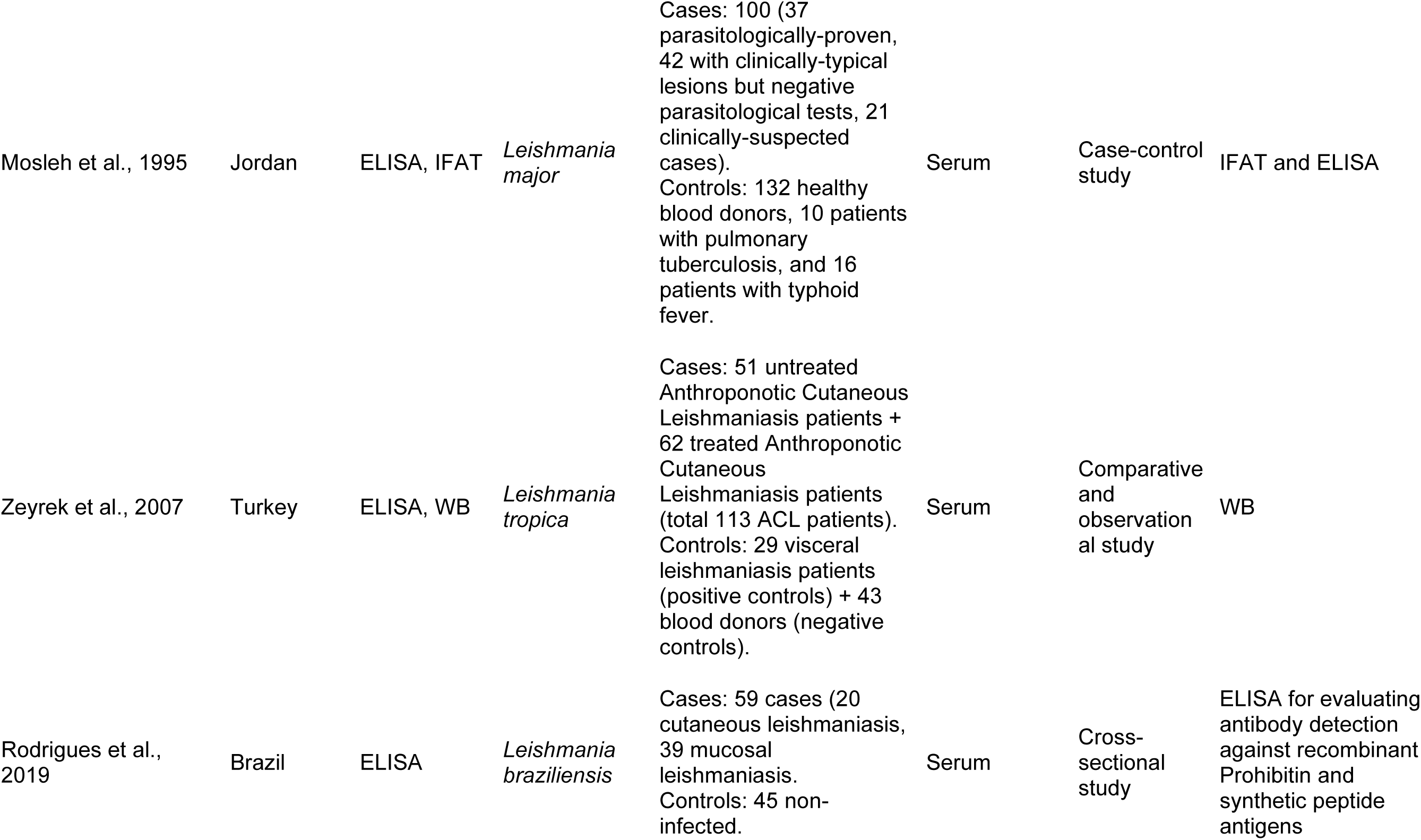
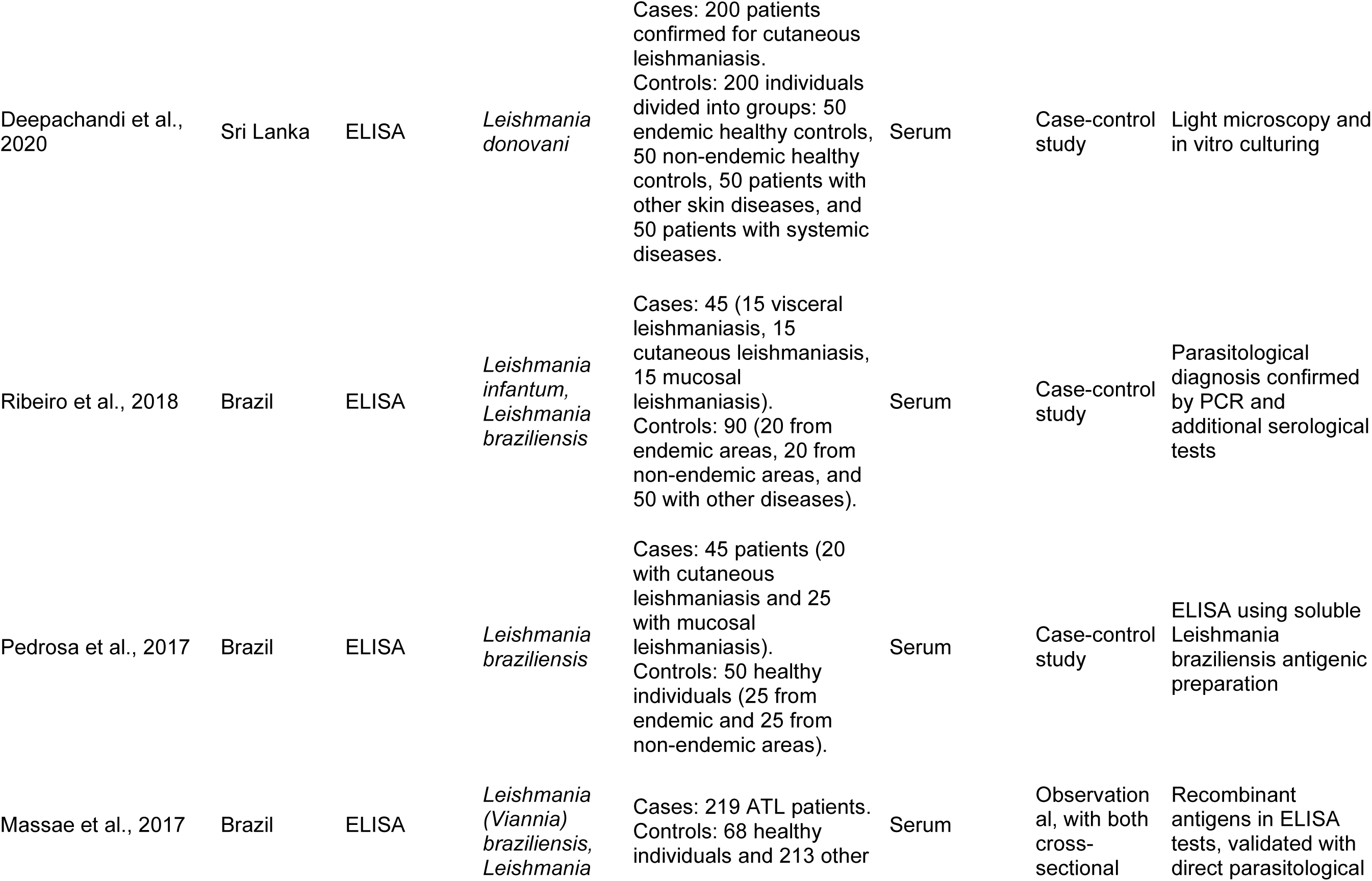
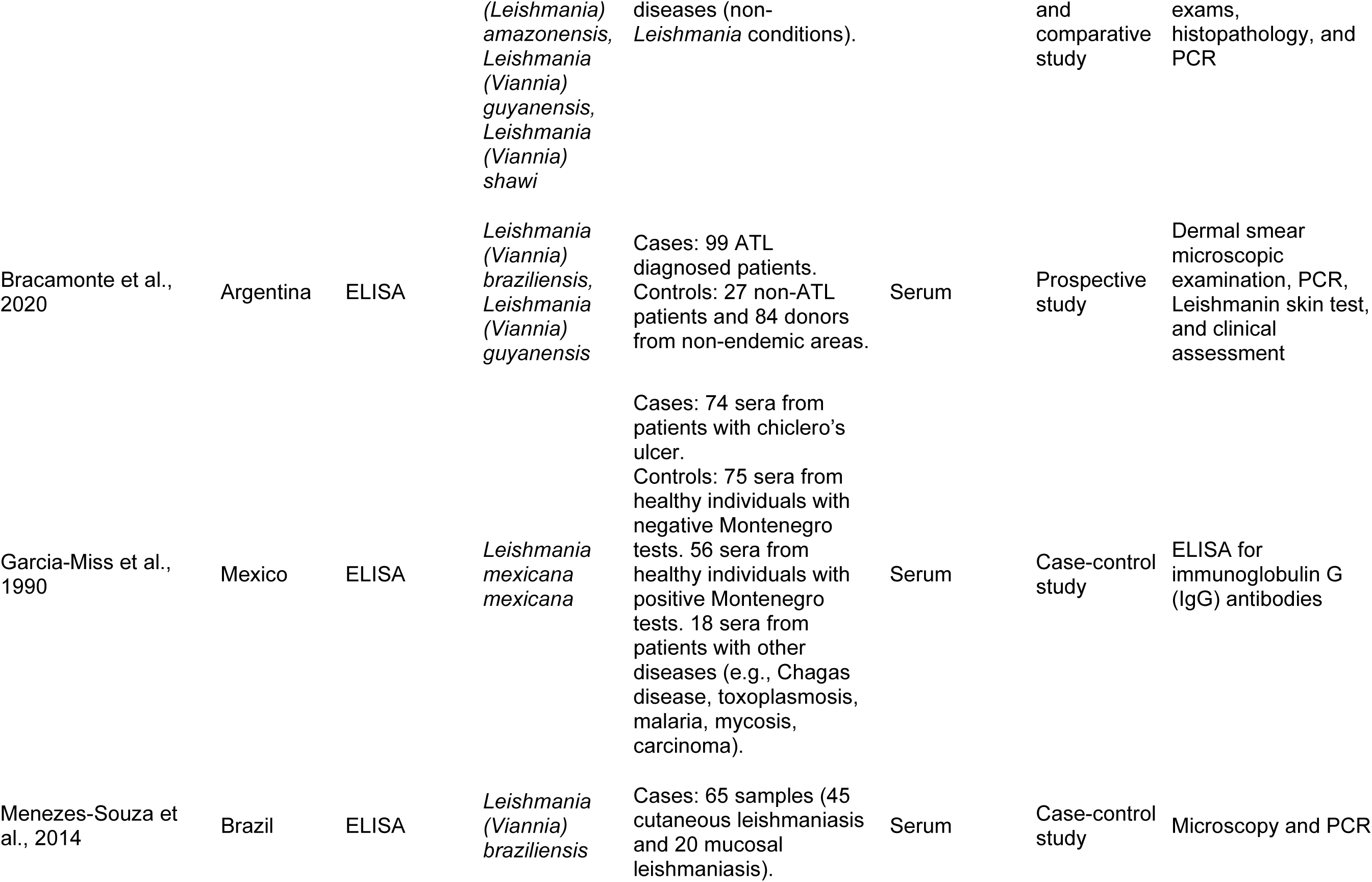
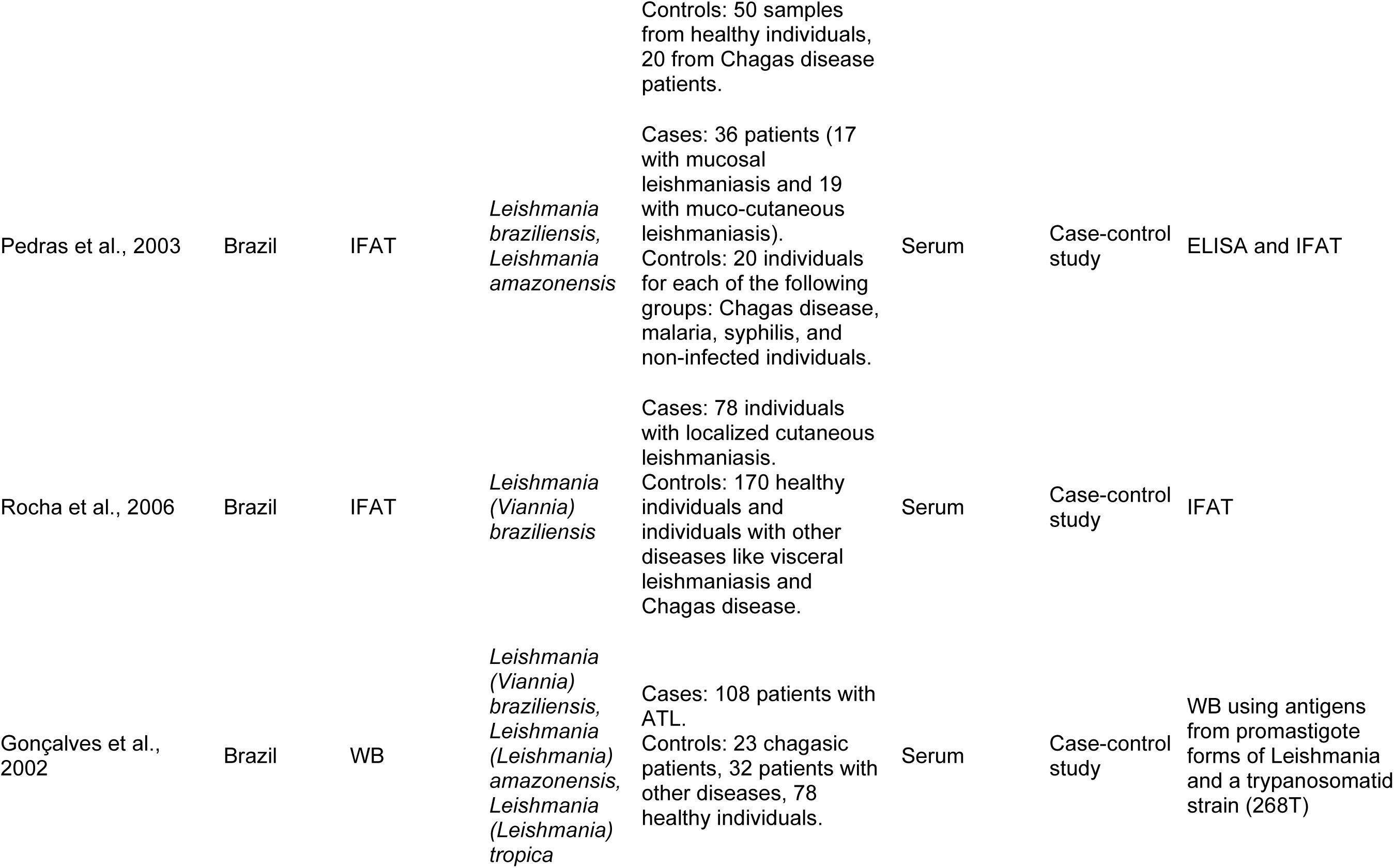
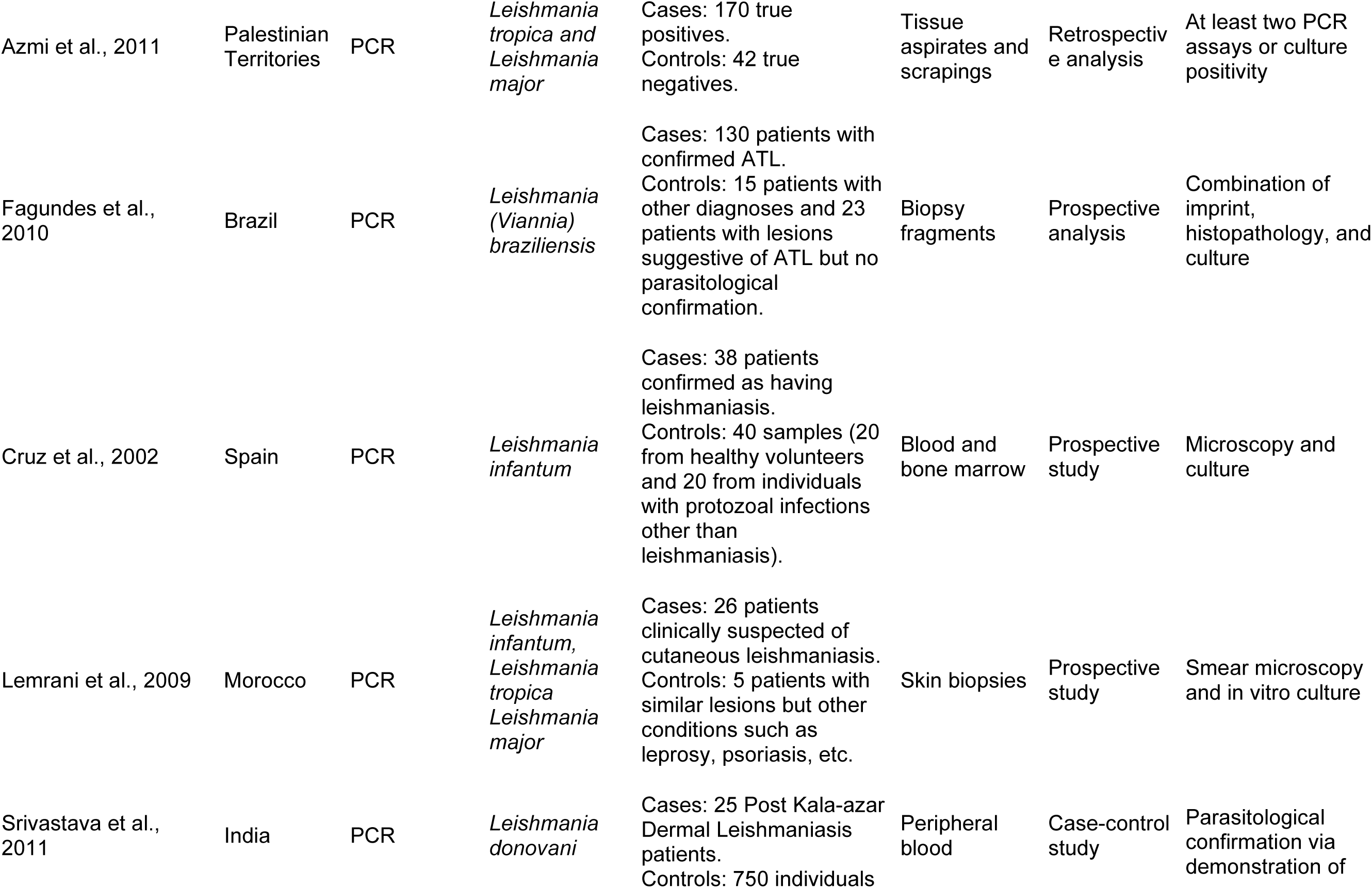
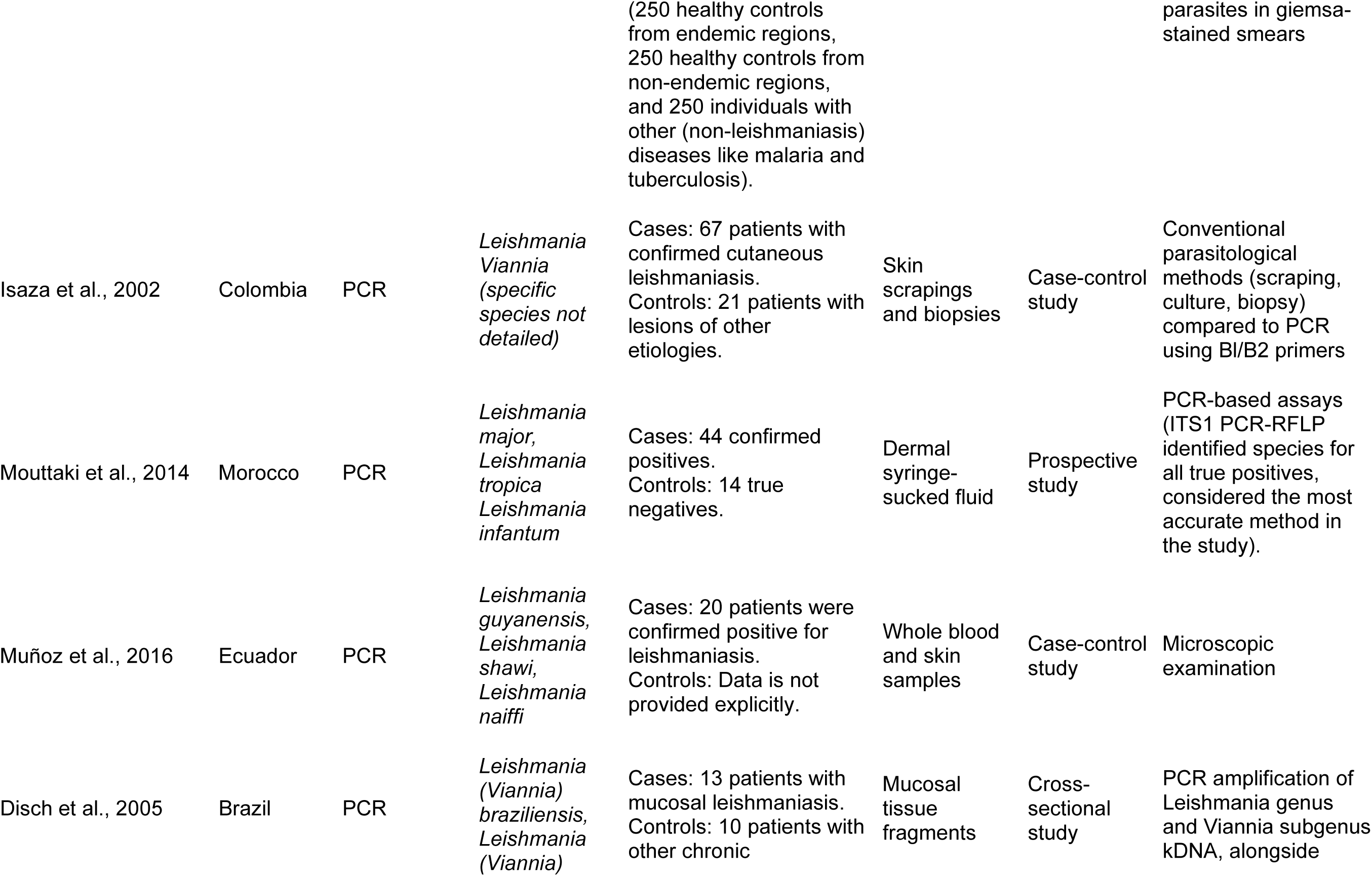
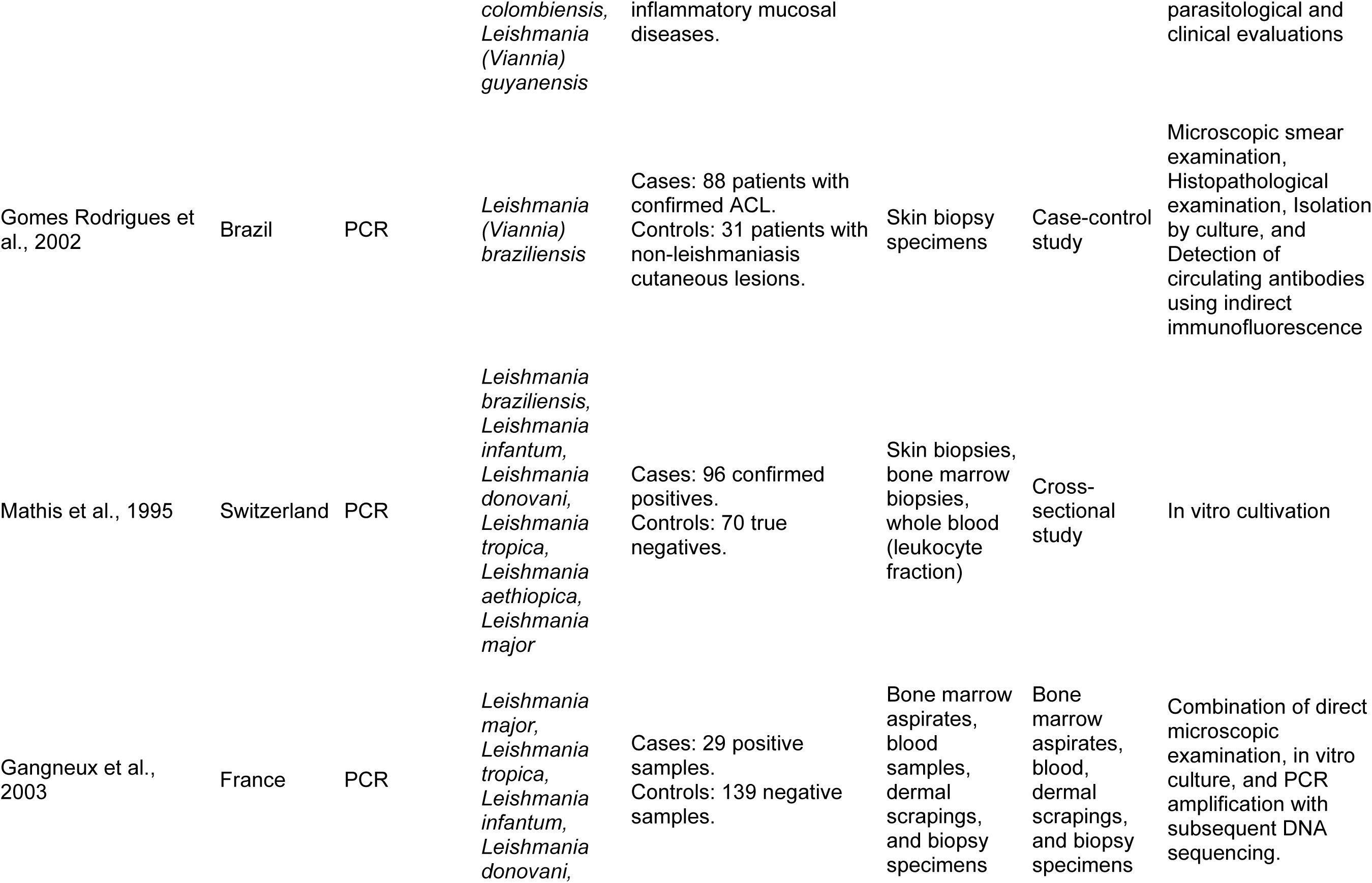
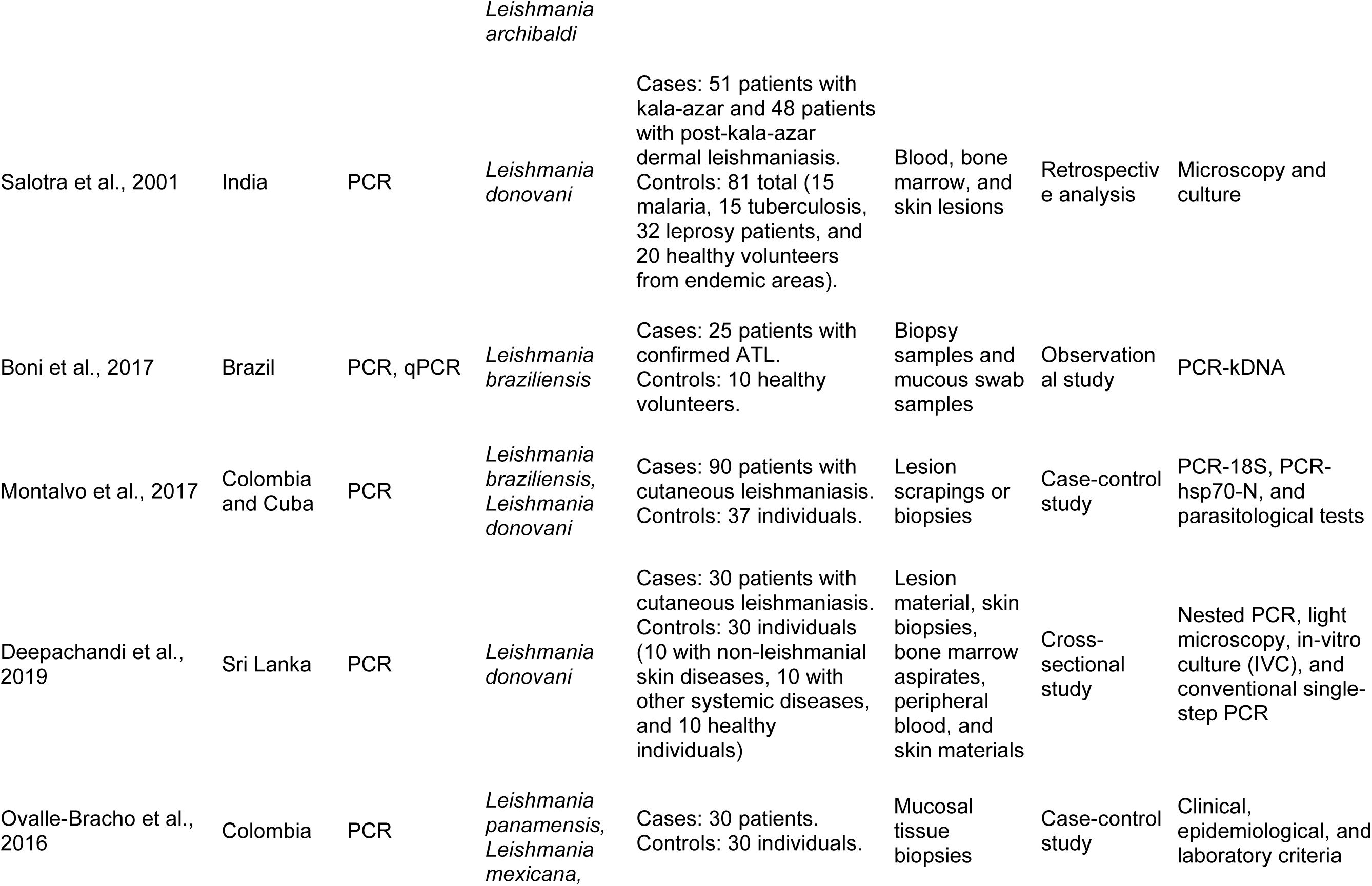
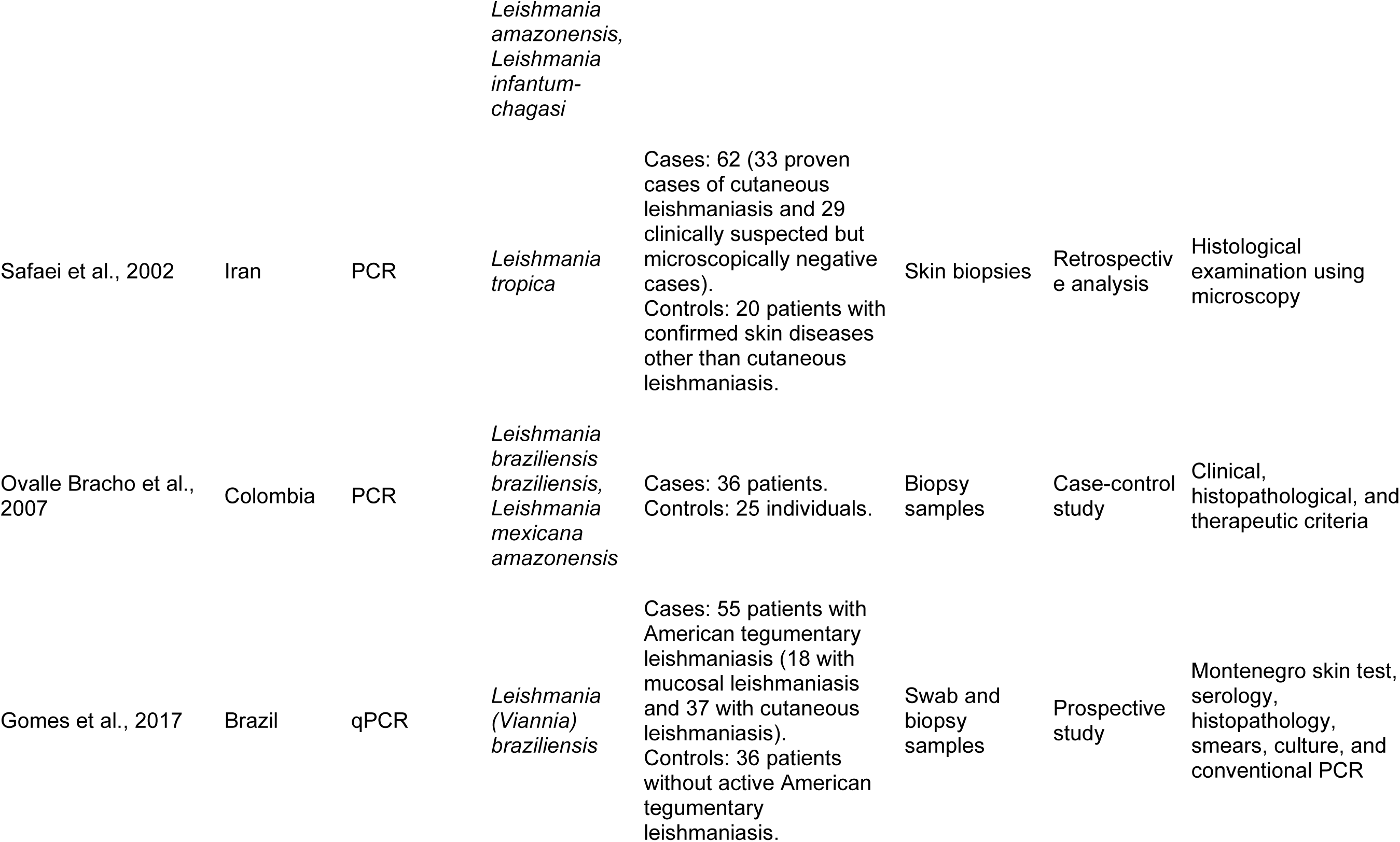
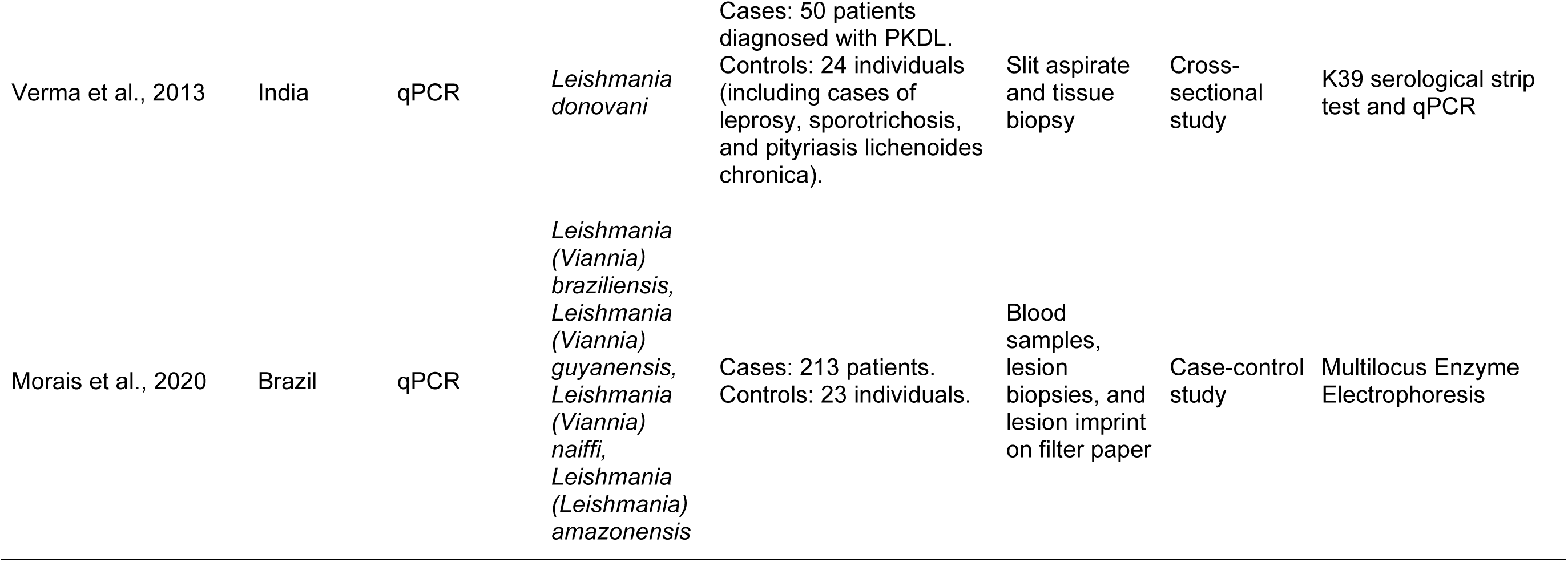
Main methodological aspects of studies on tegumentary leishmaniasis.

**S3 Table.**
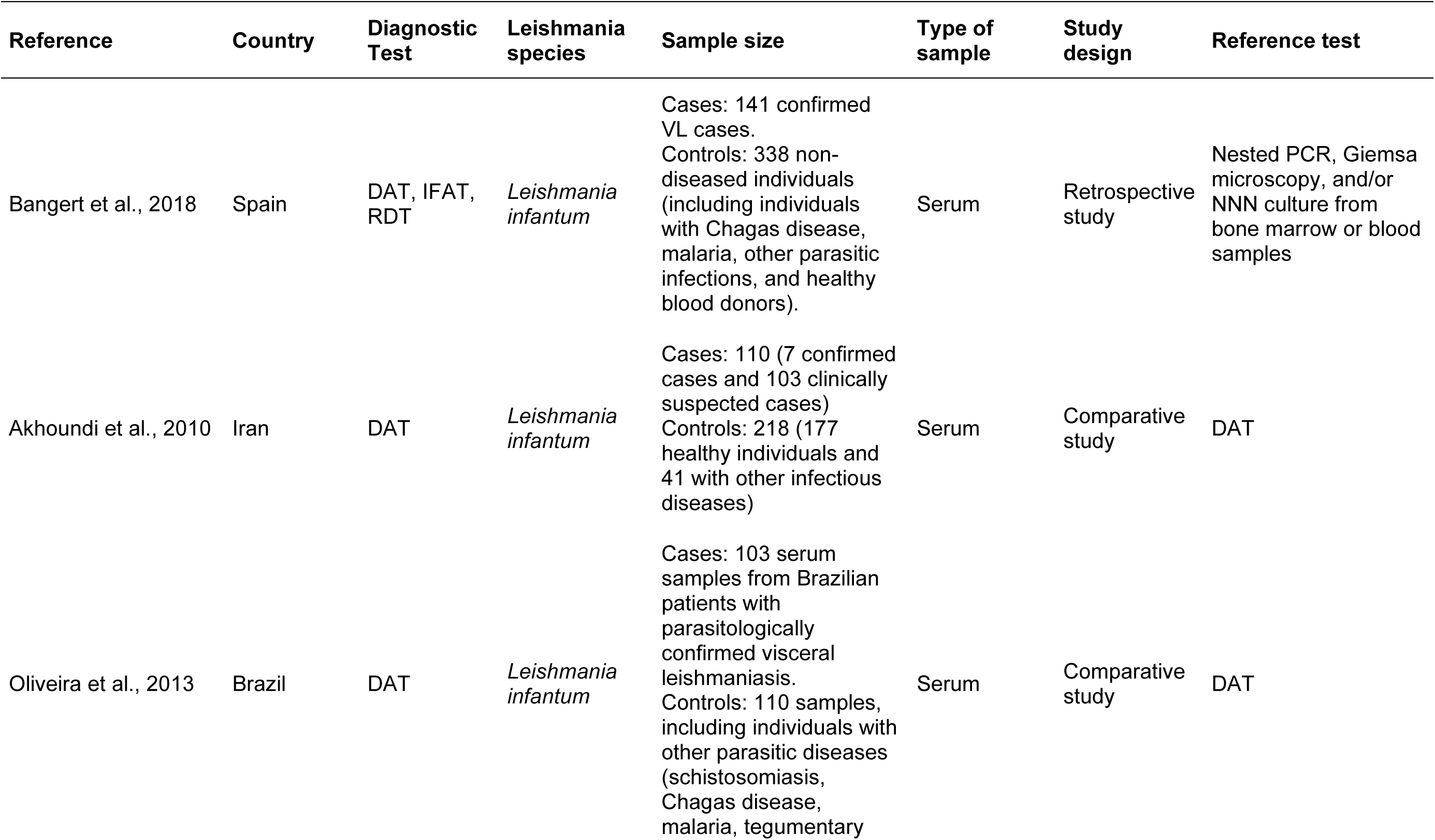
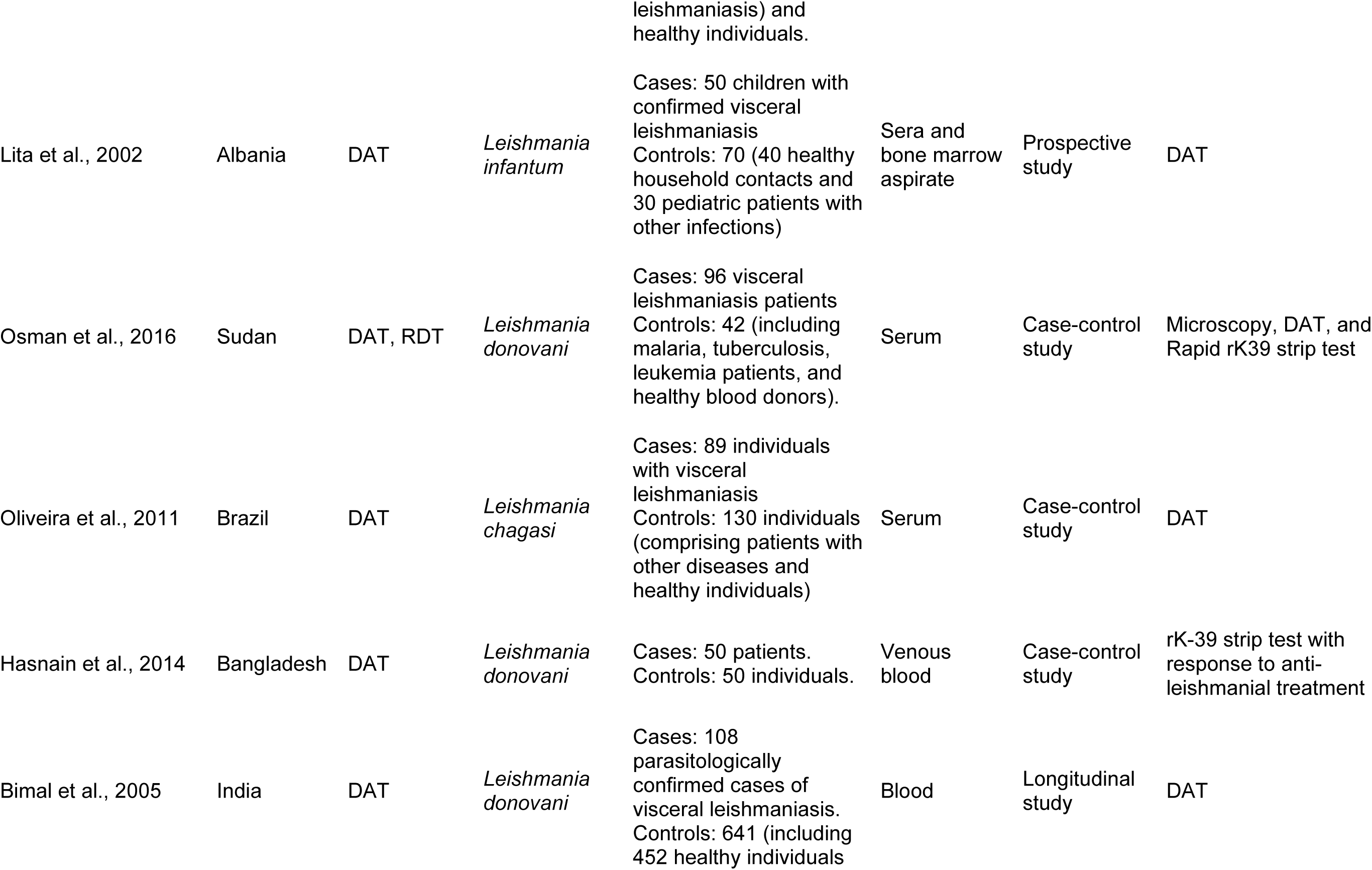
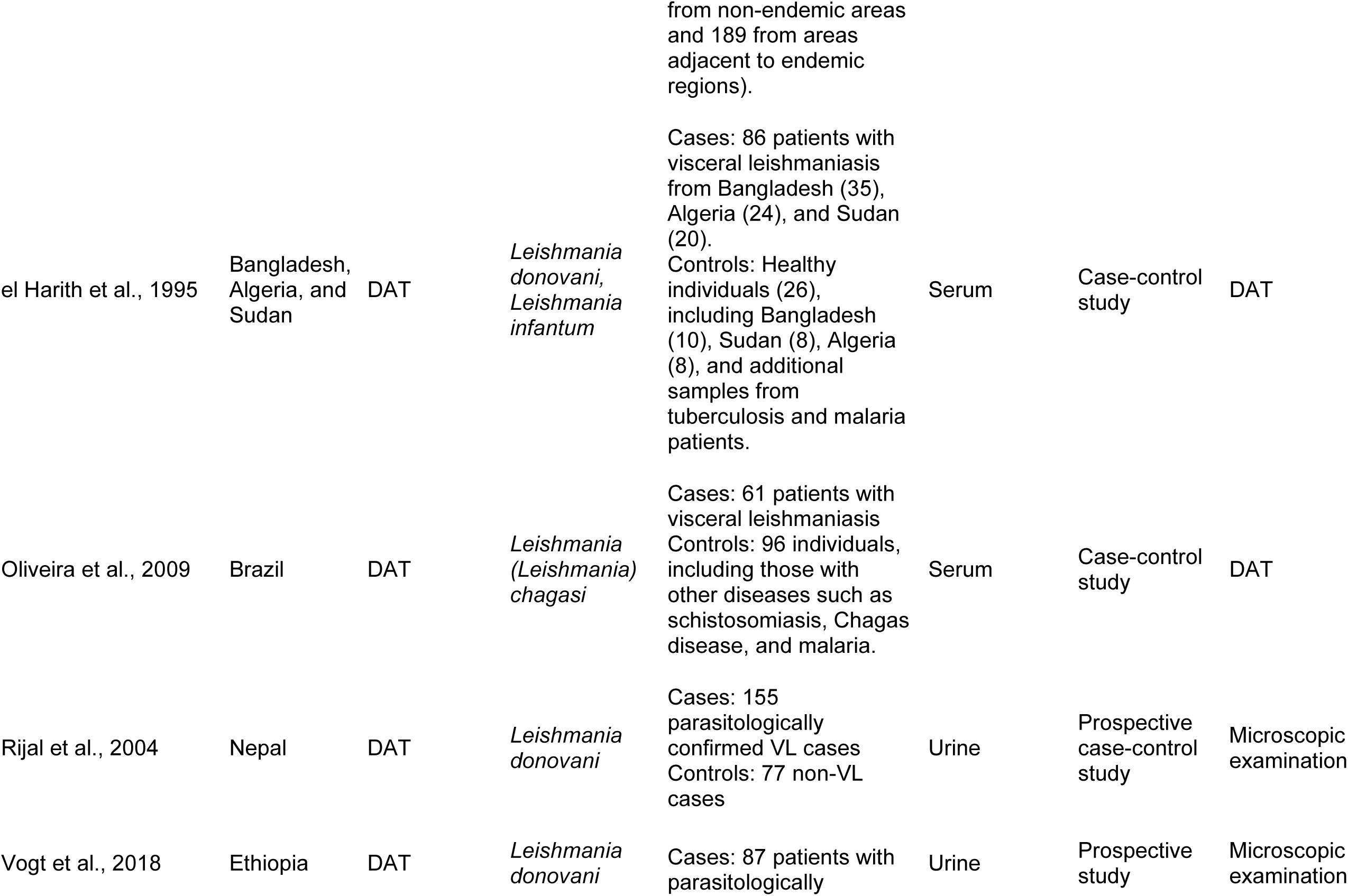
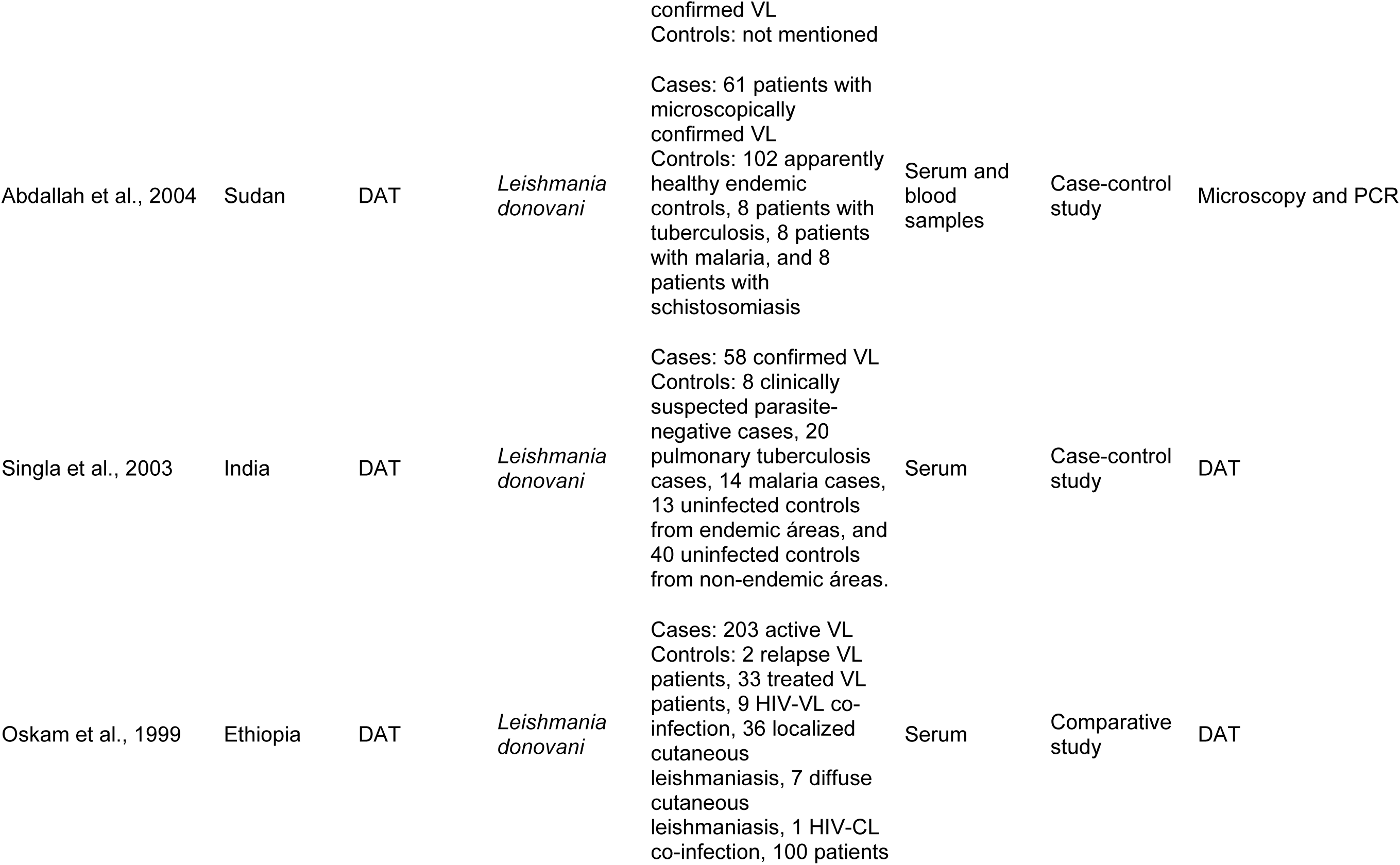
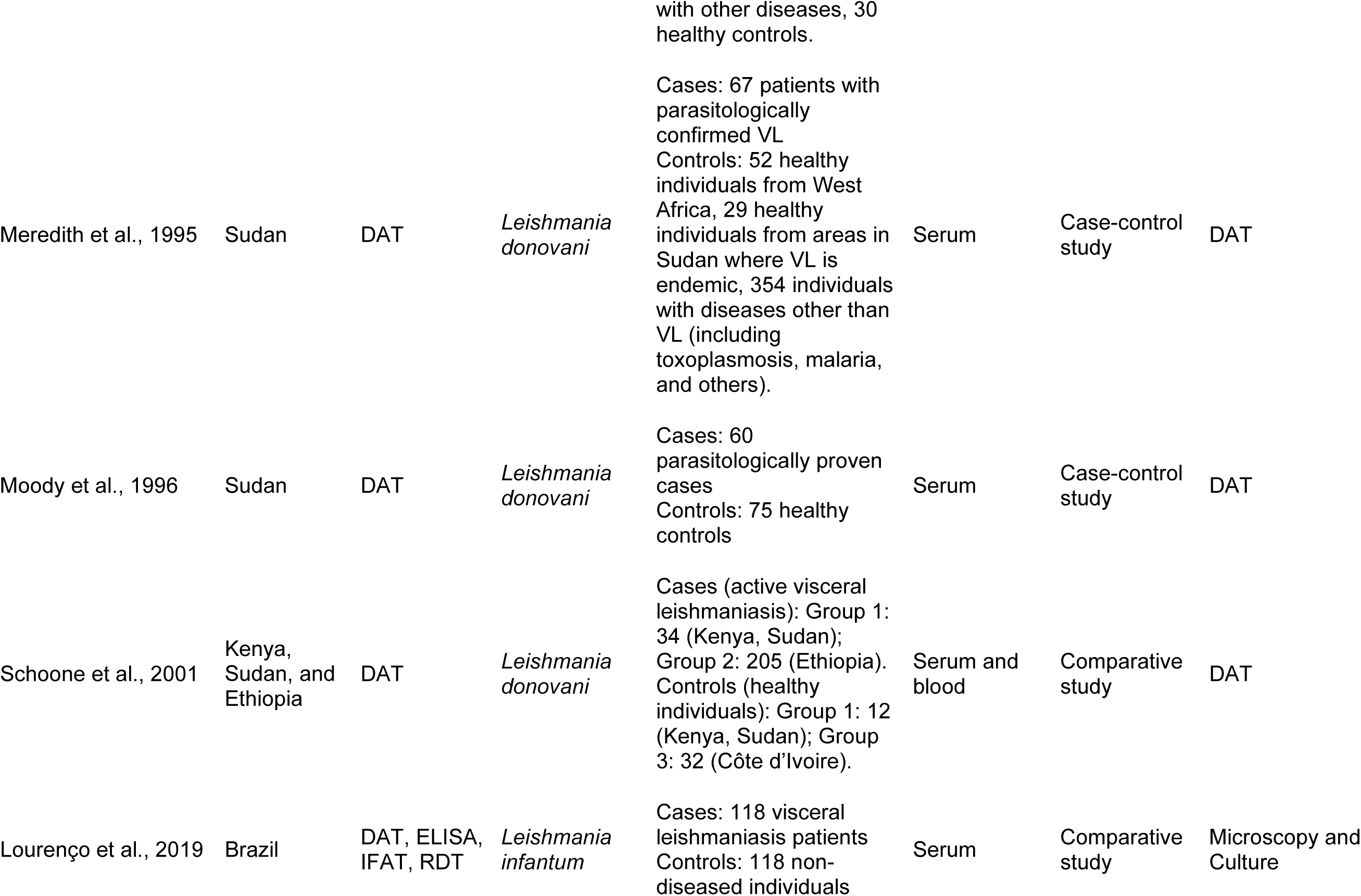
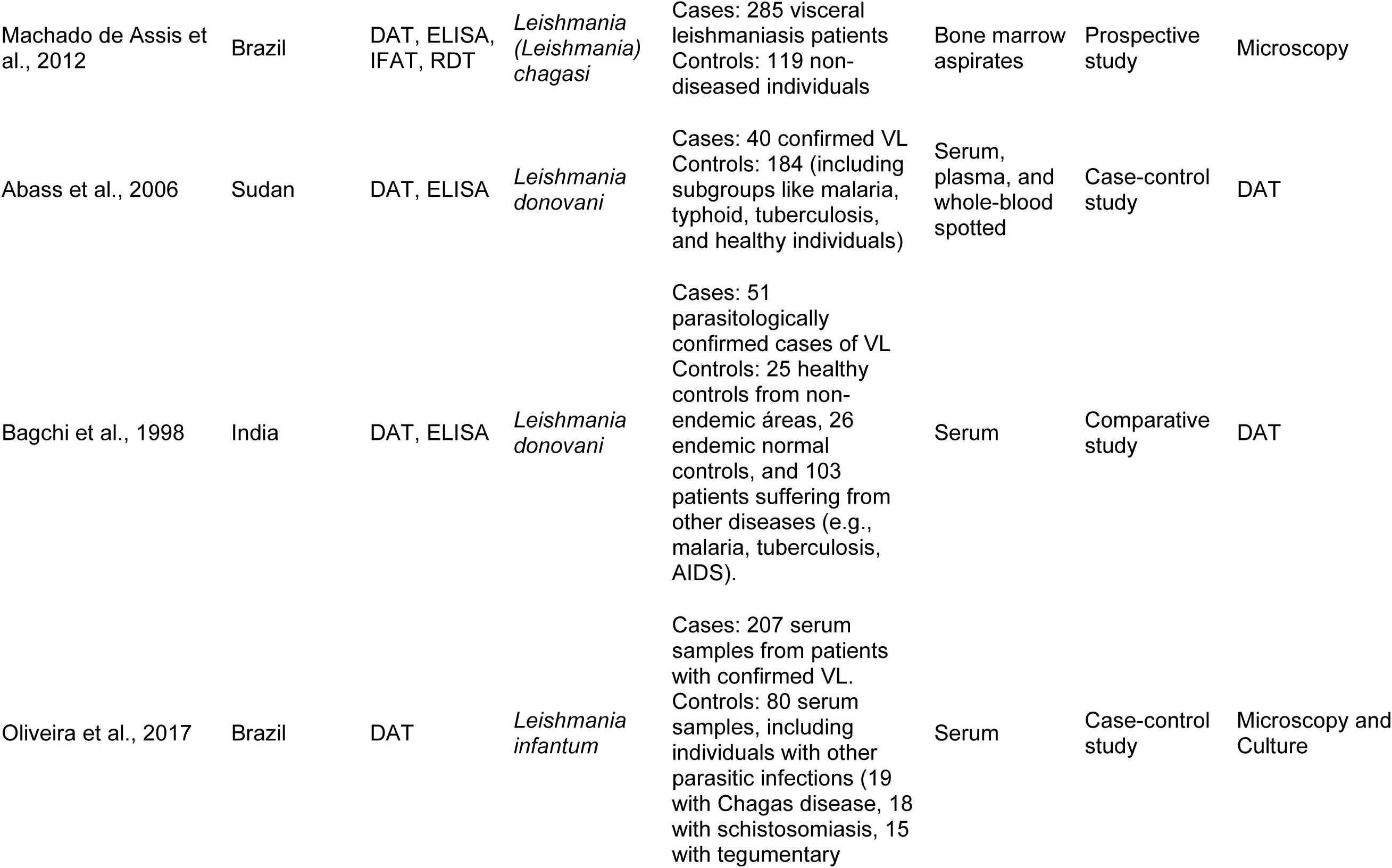
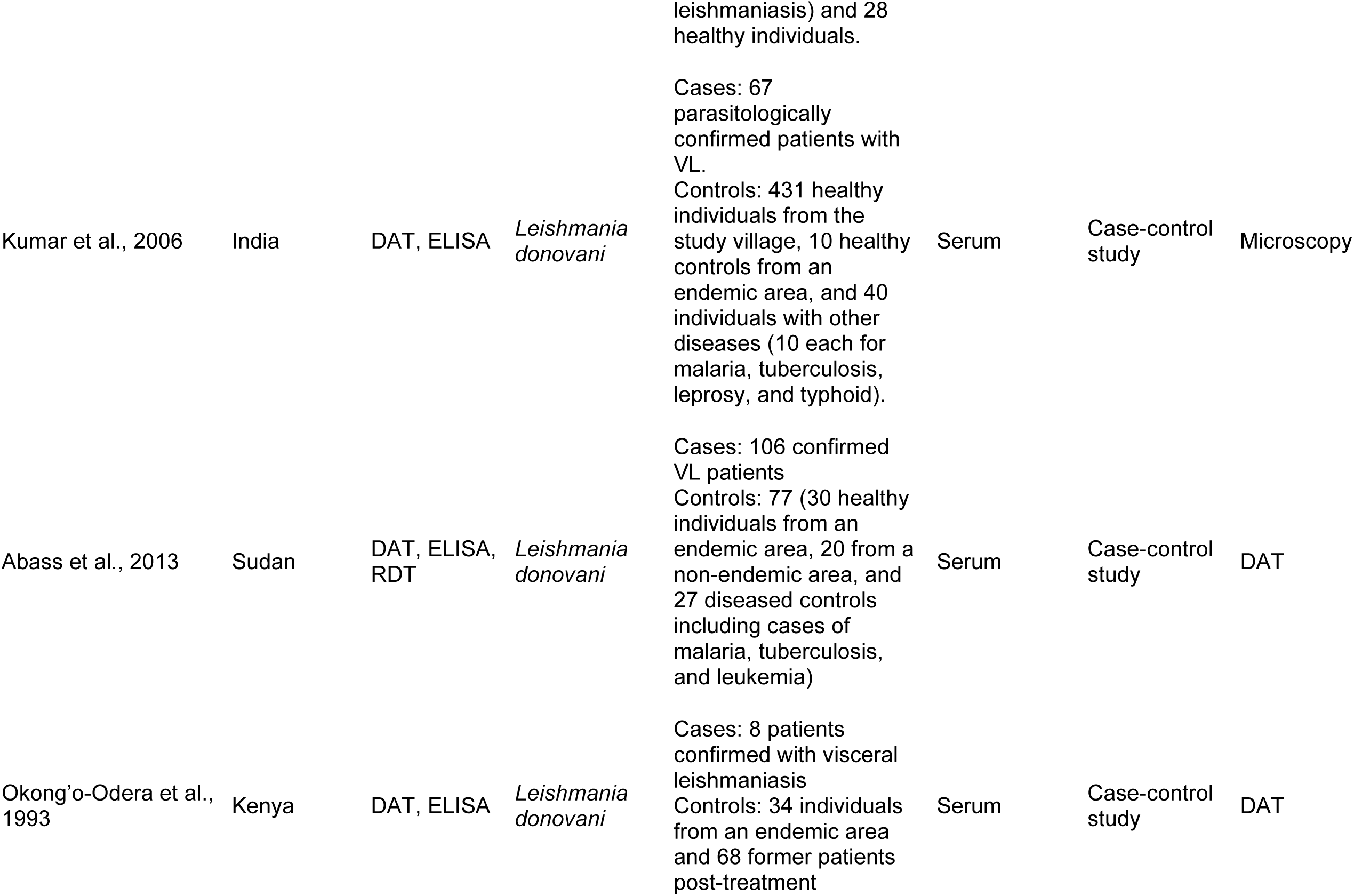
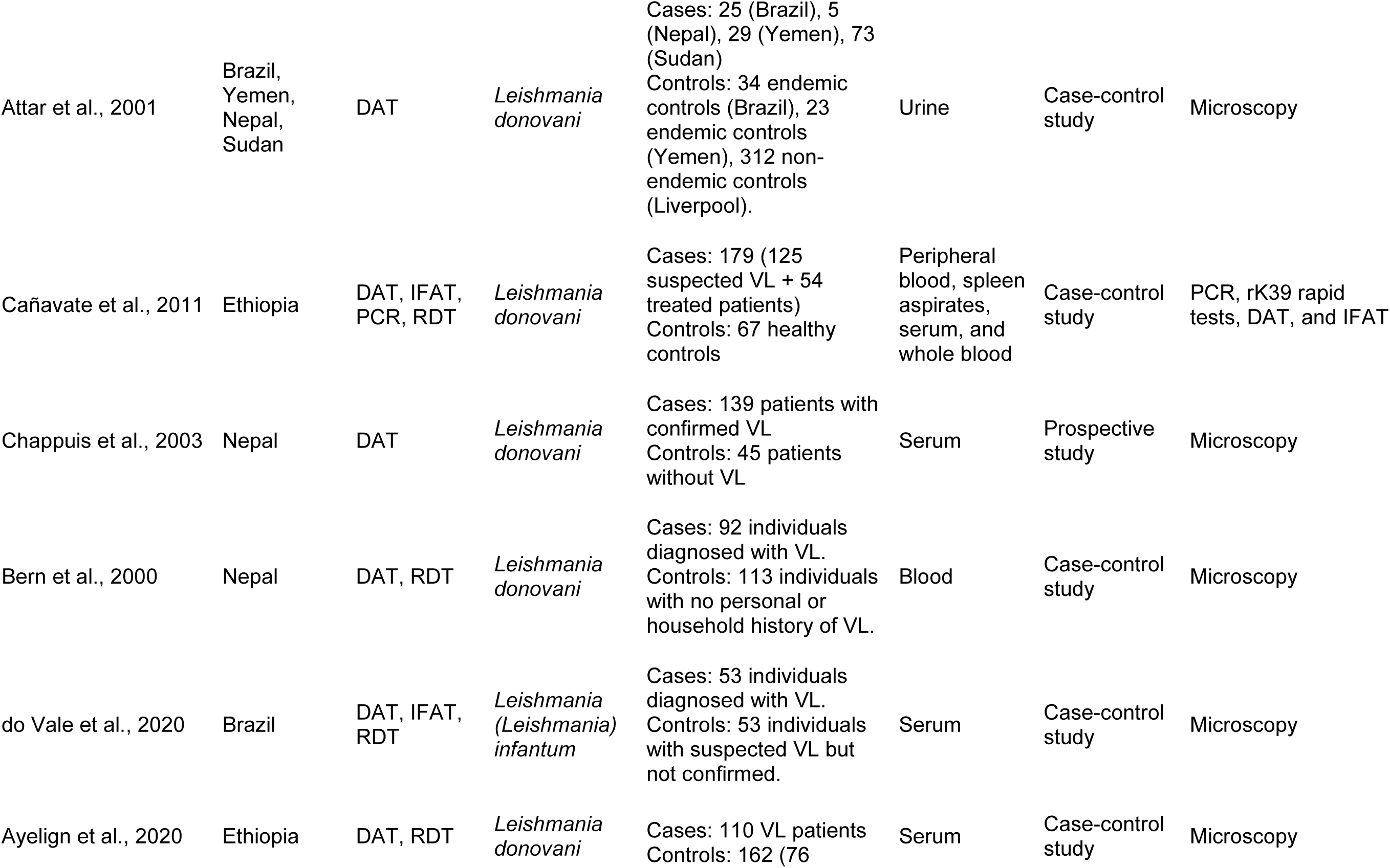
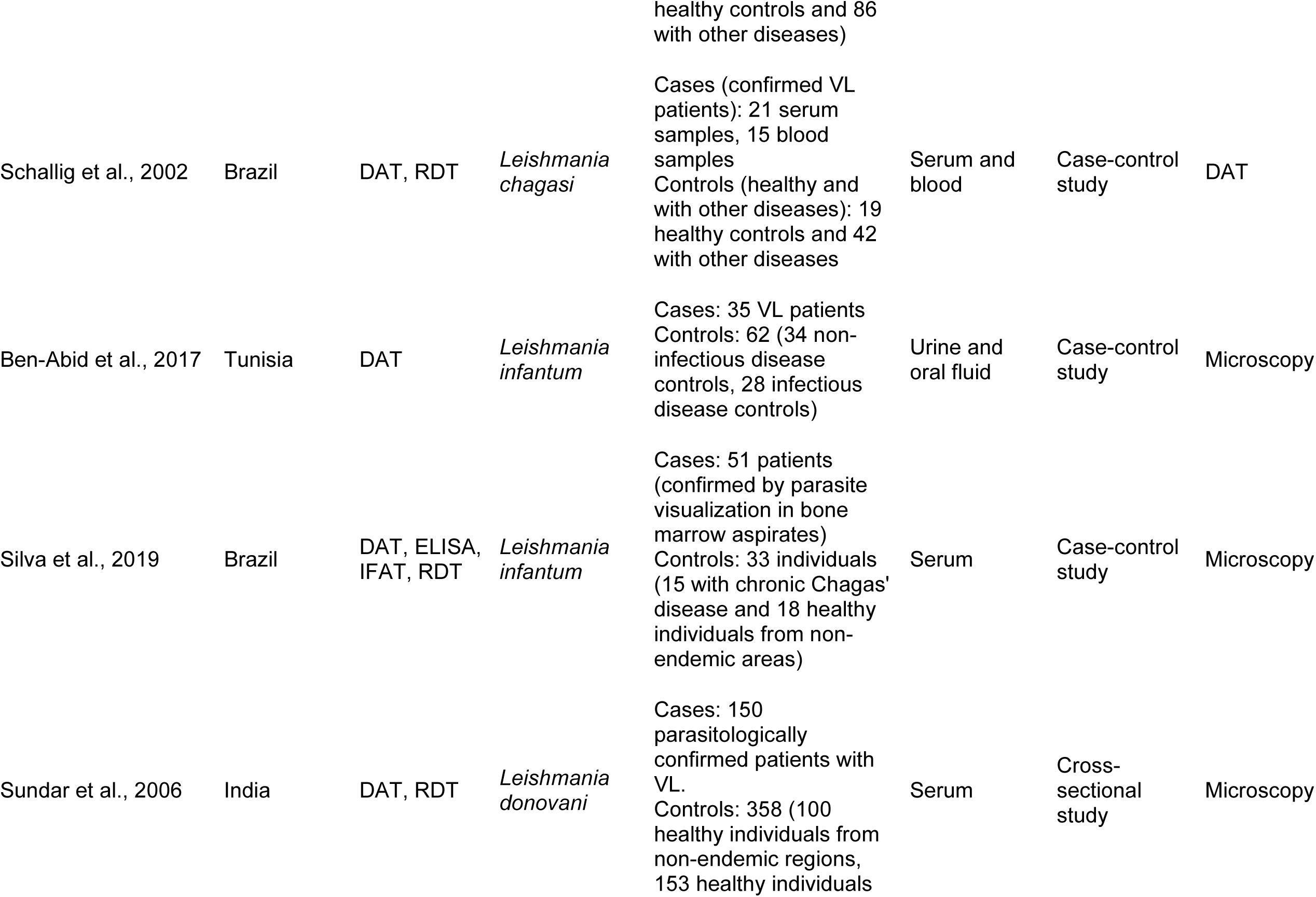
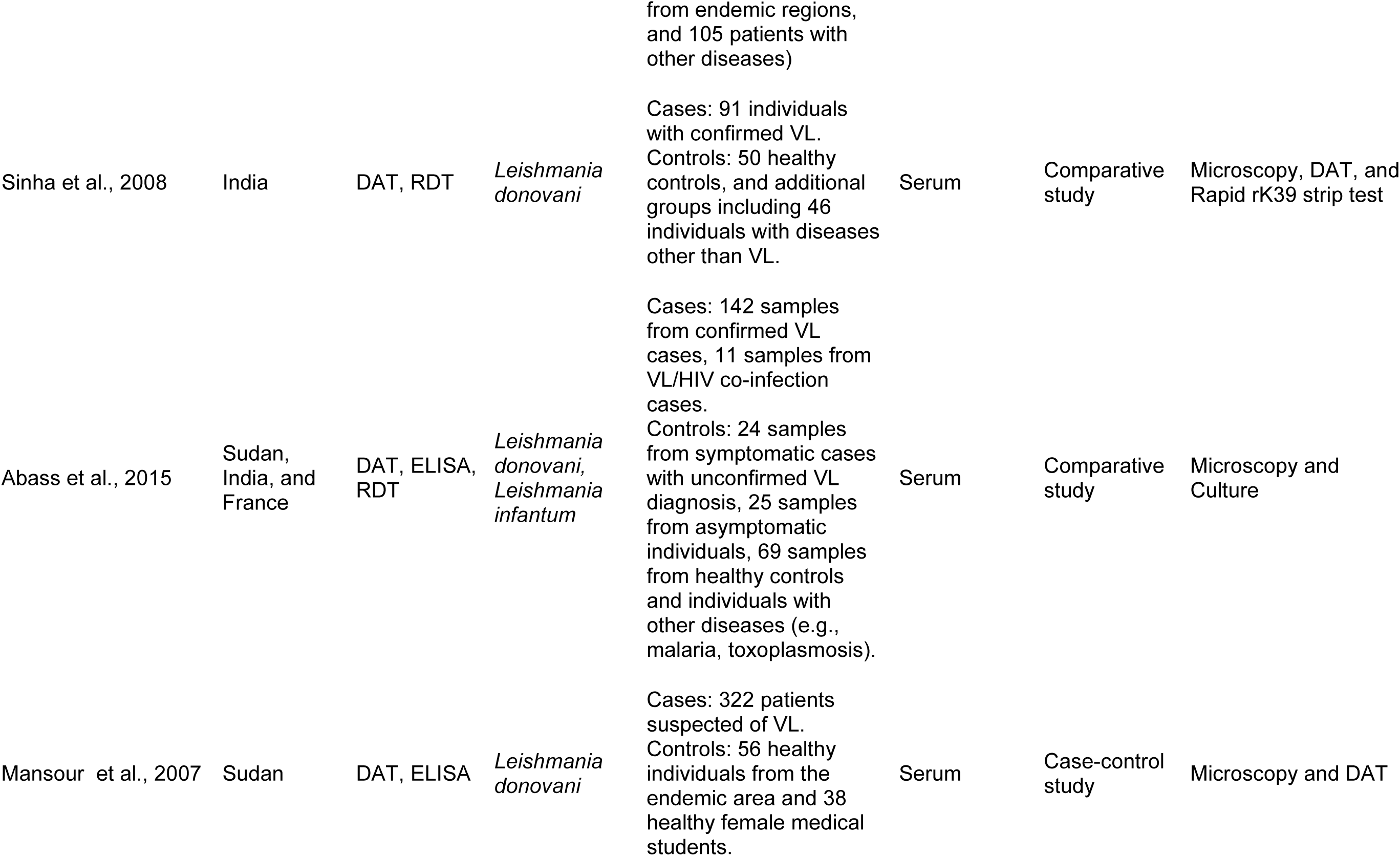
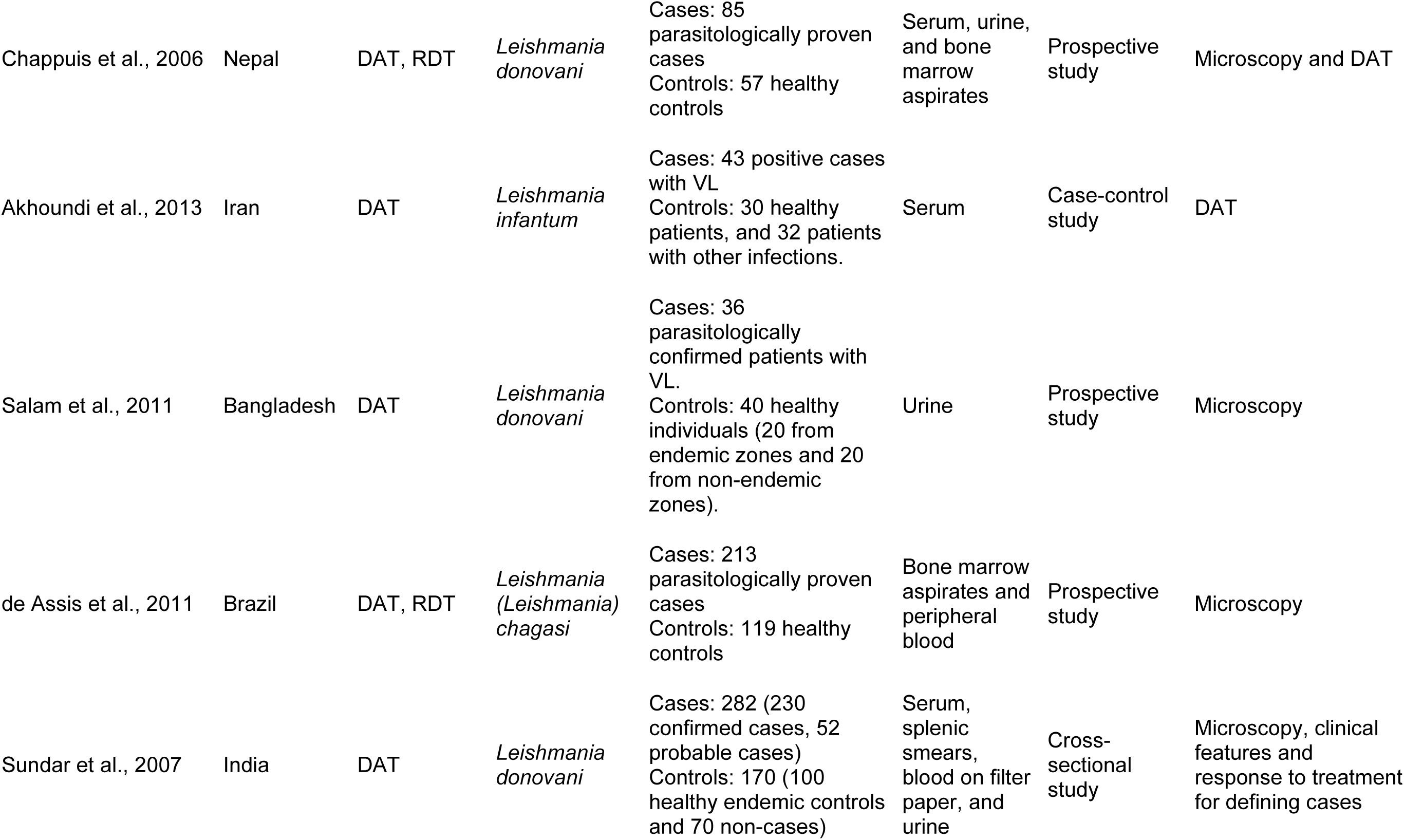
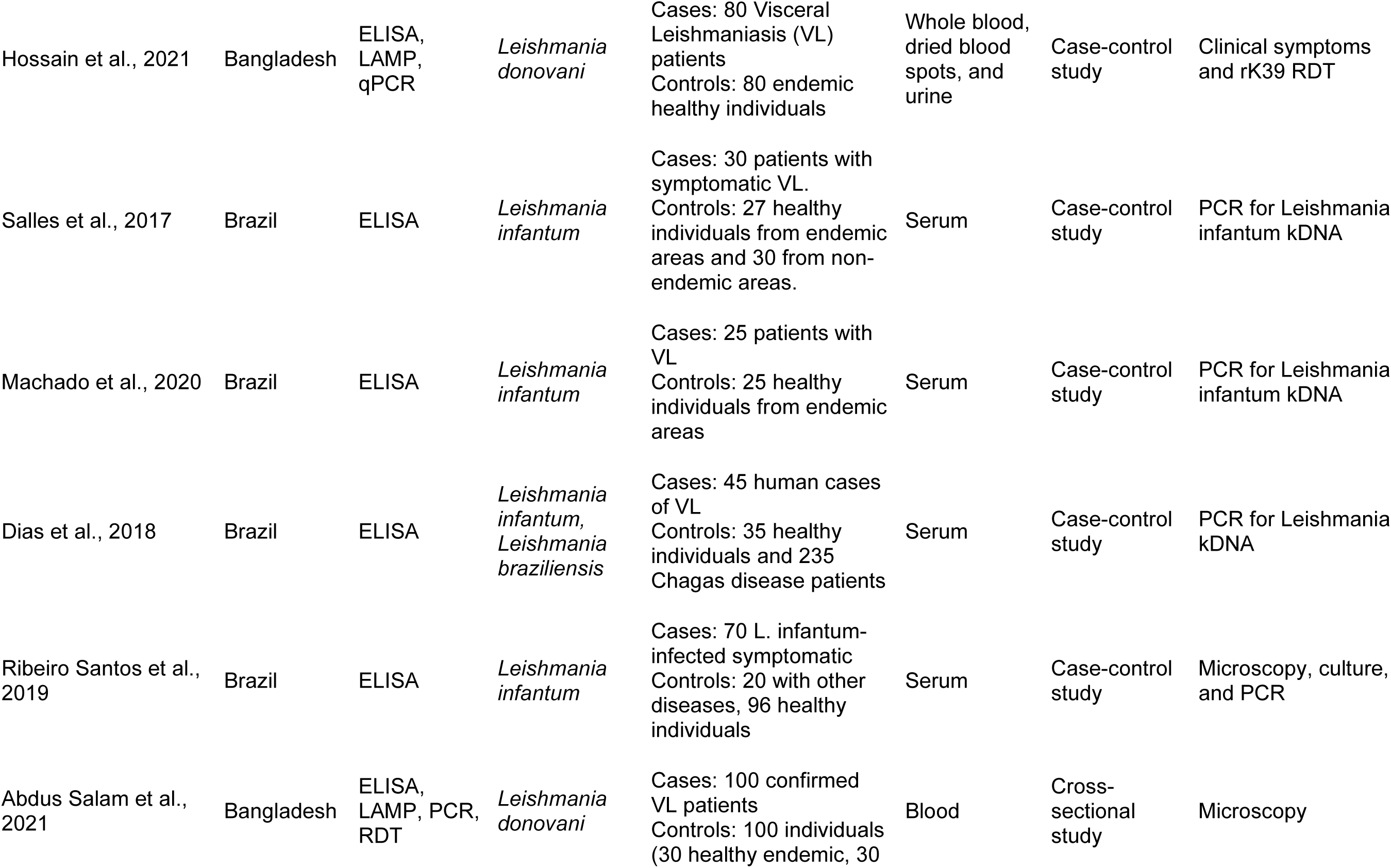
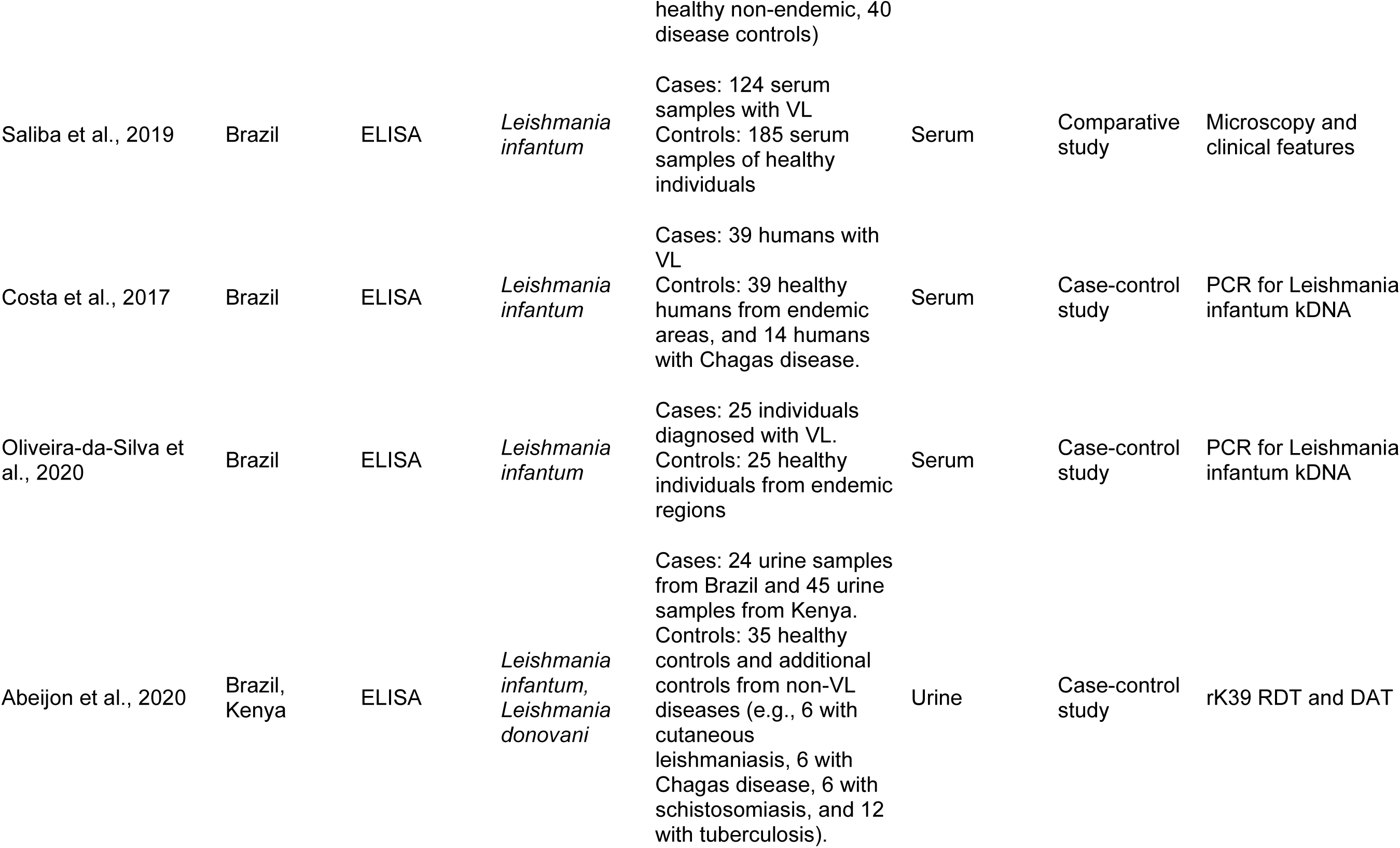
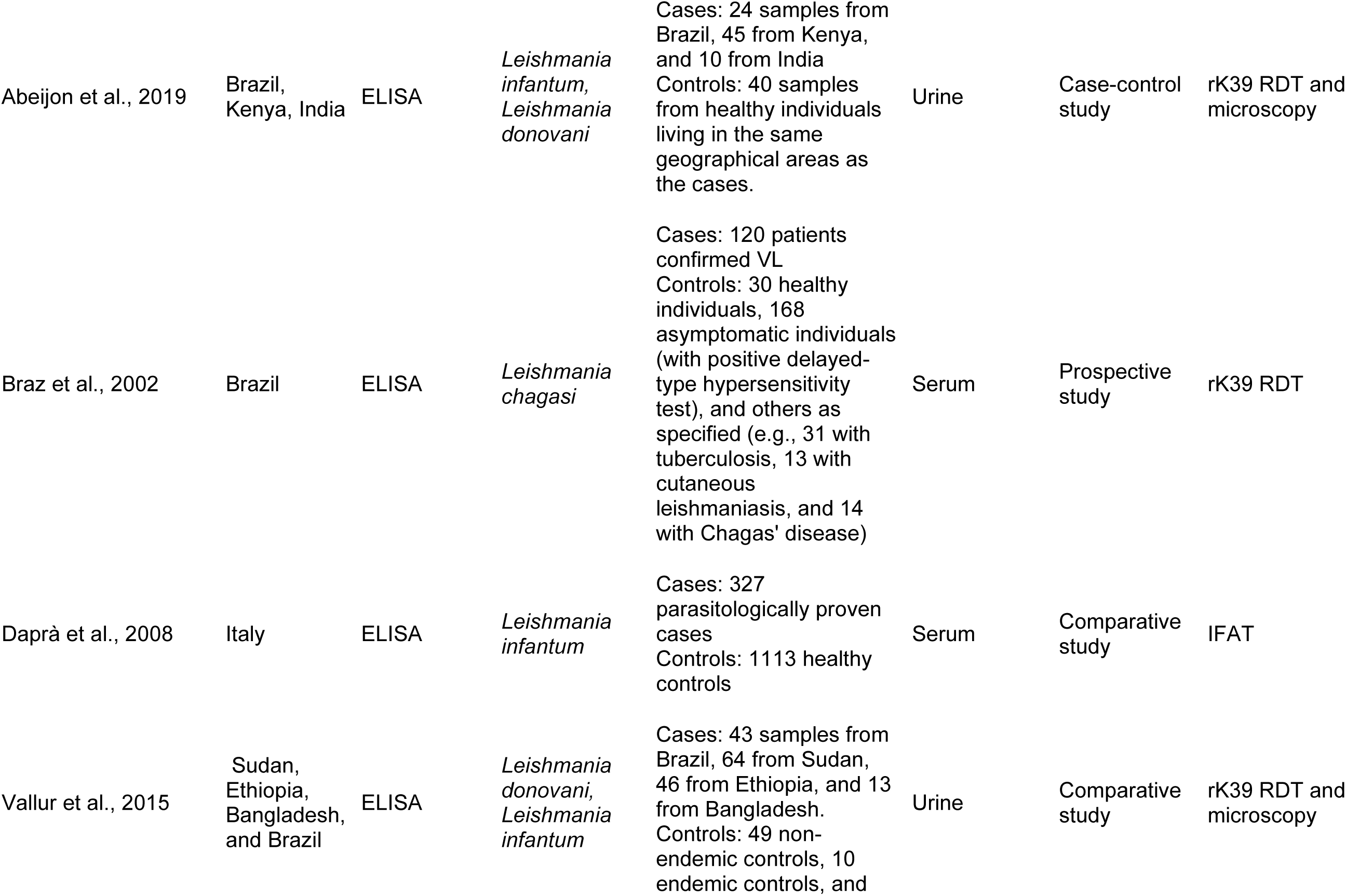
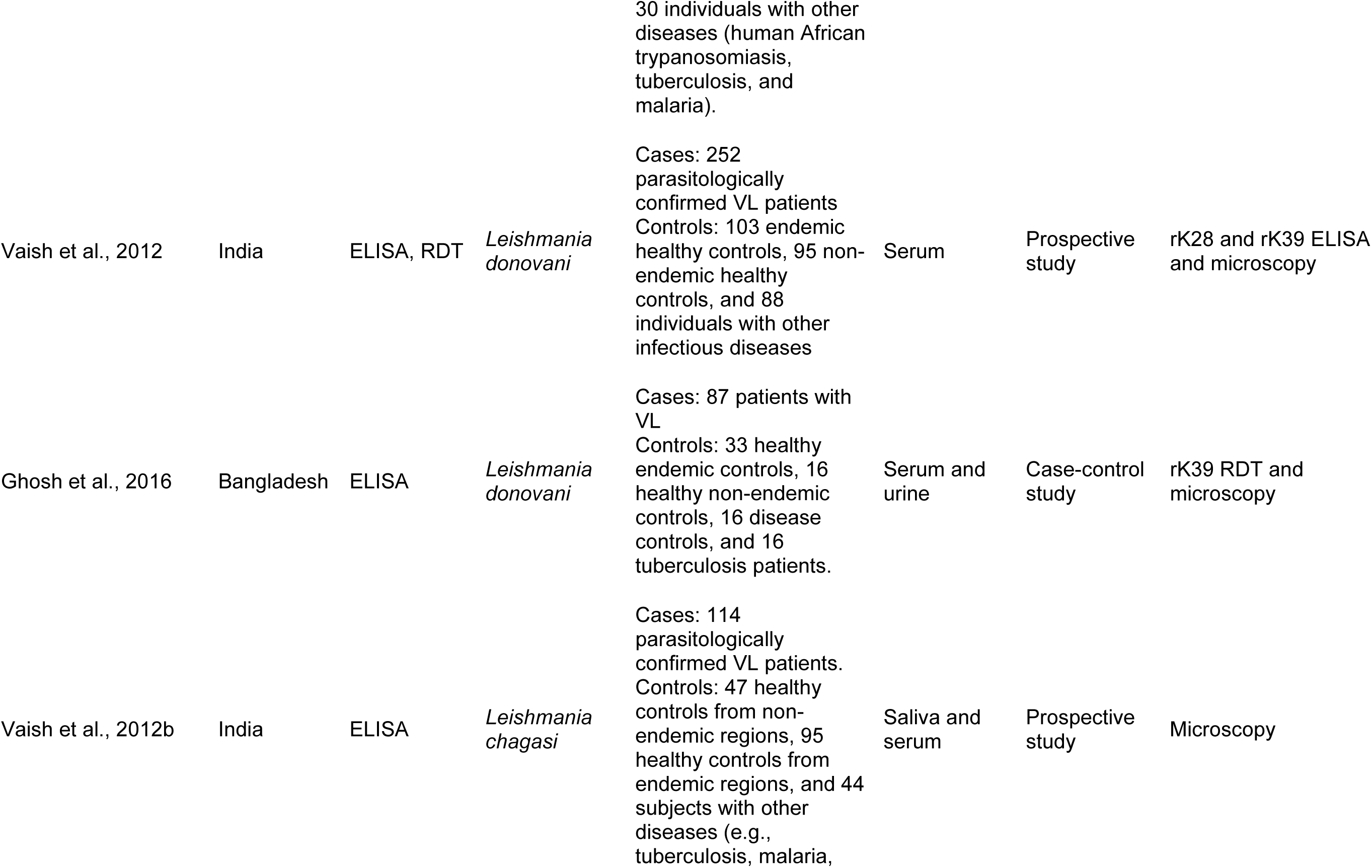
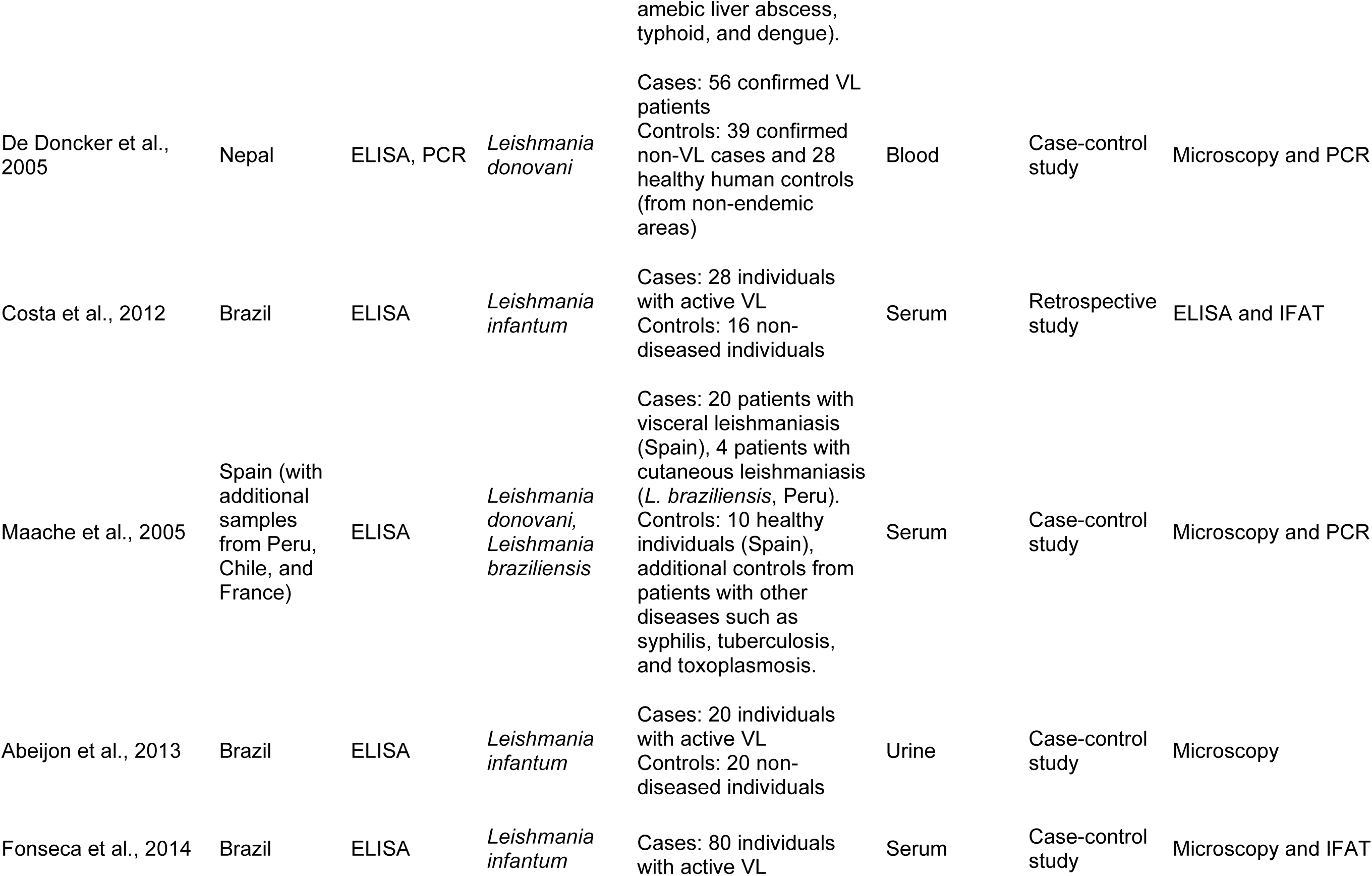
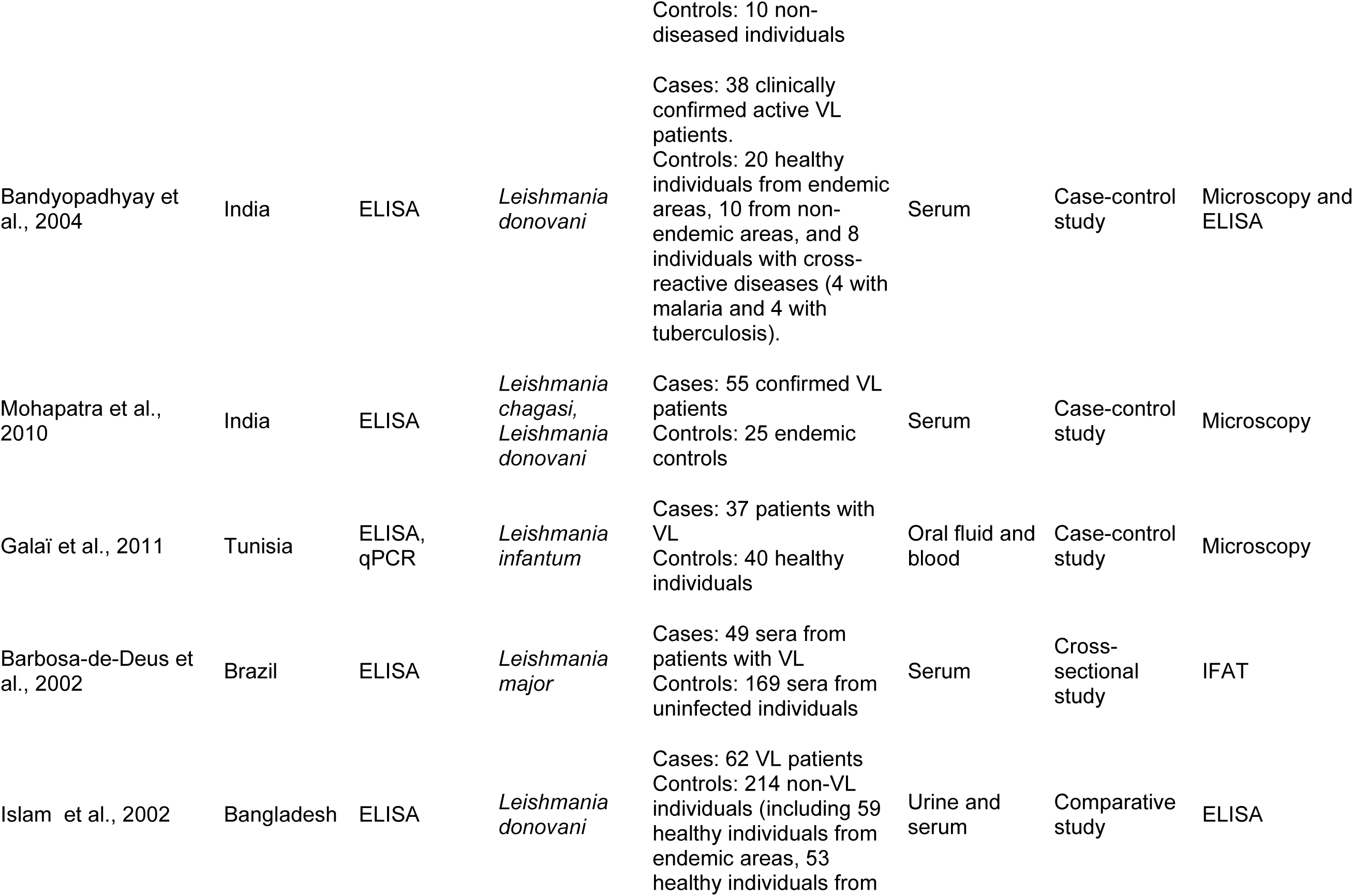
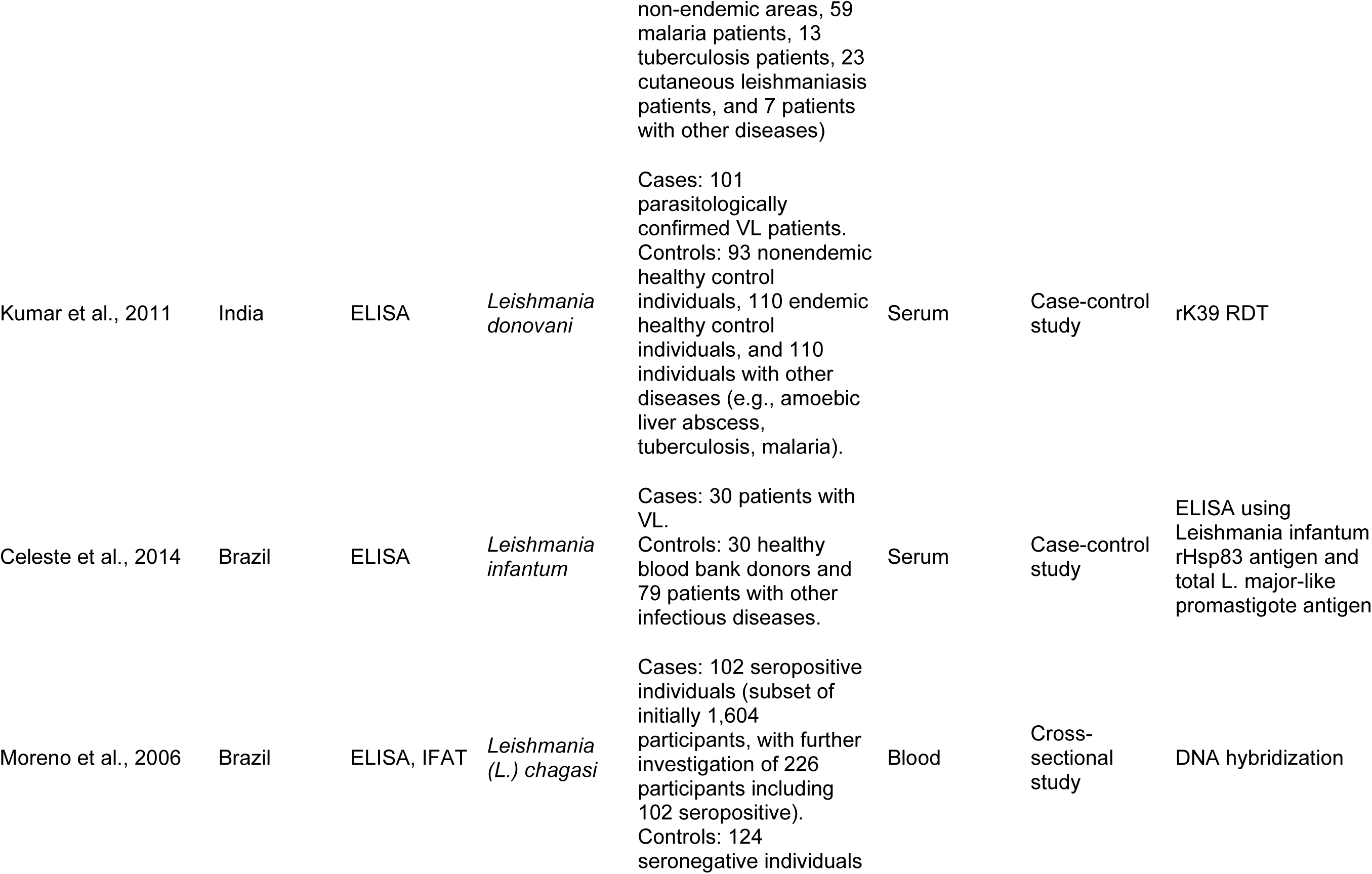
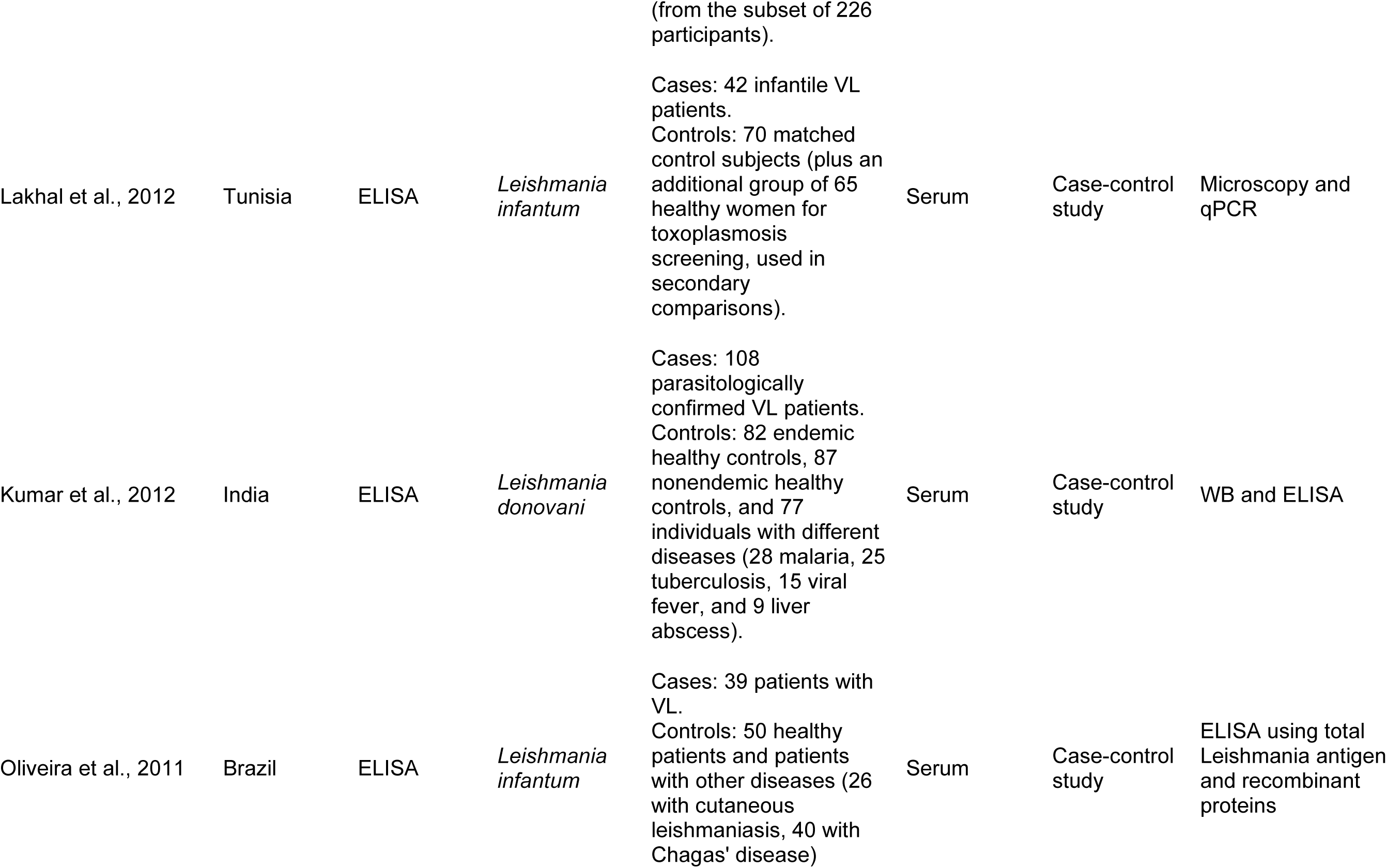
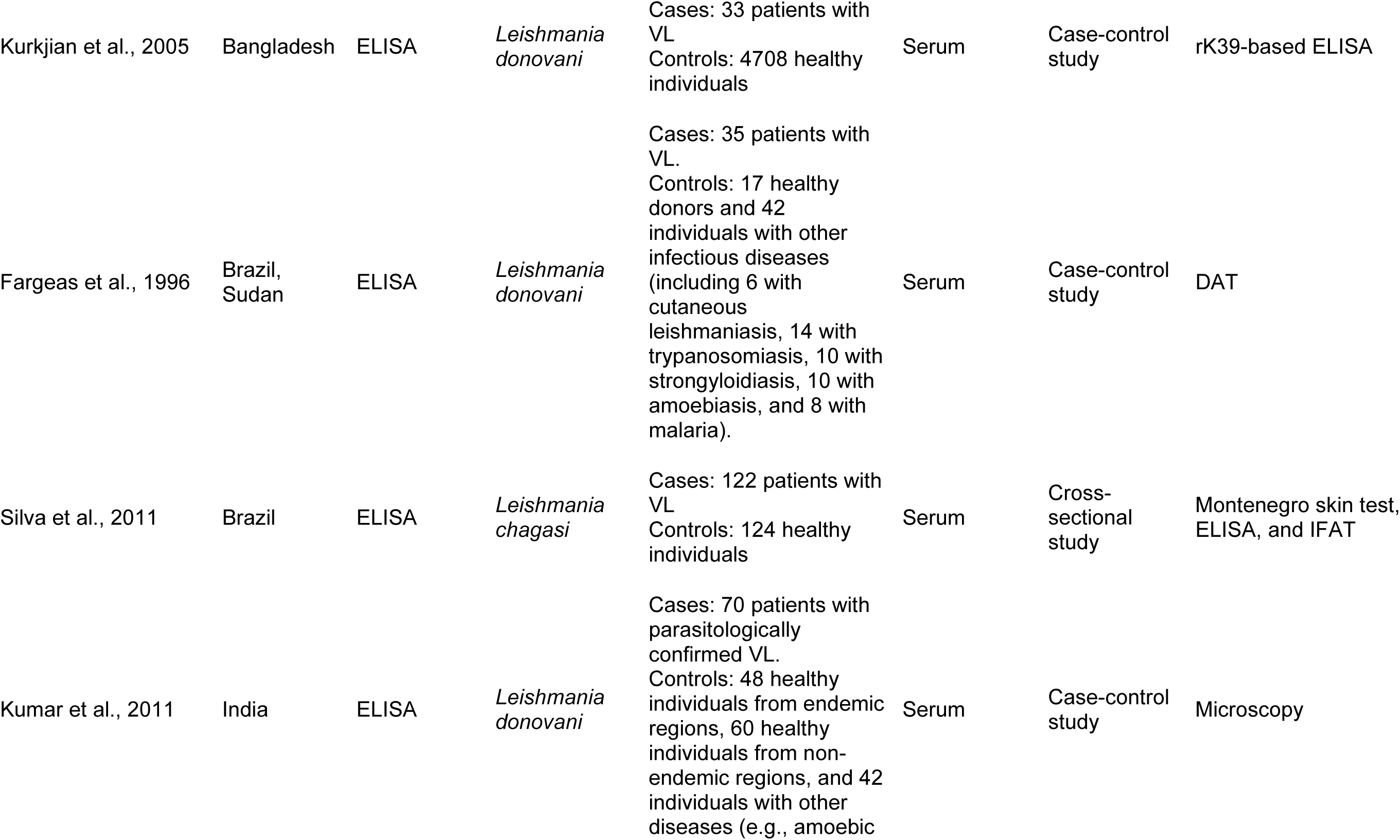
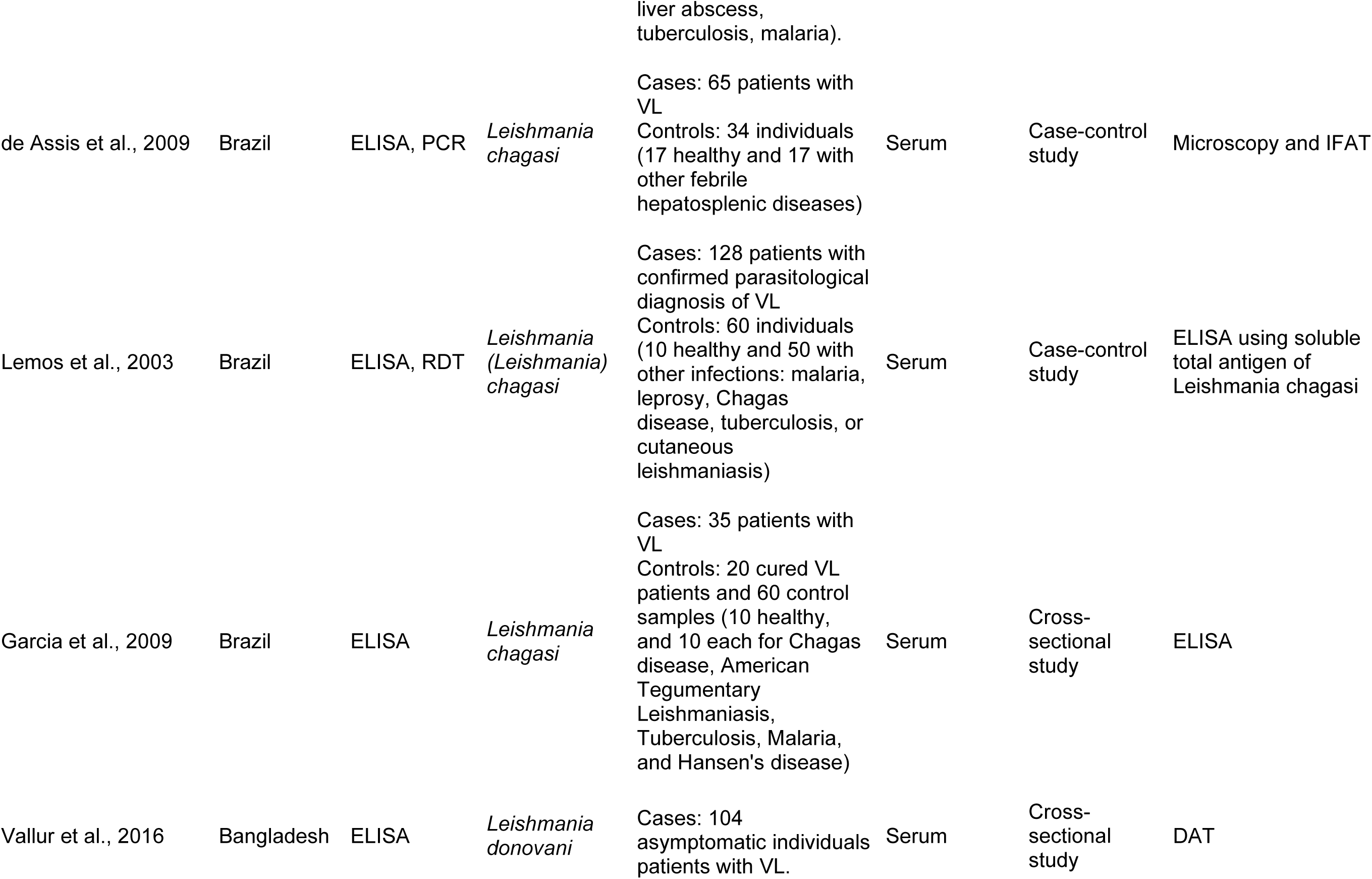
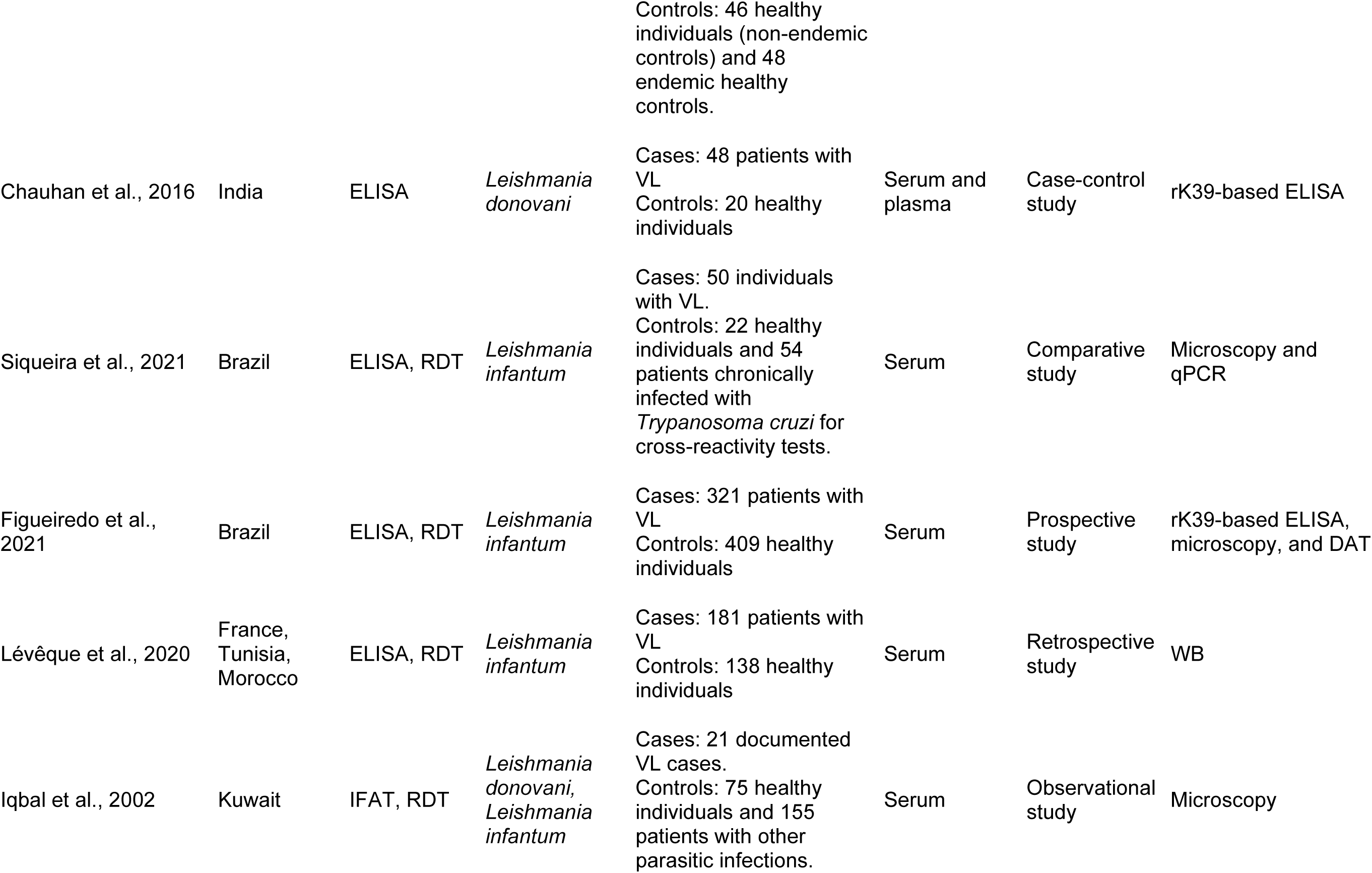
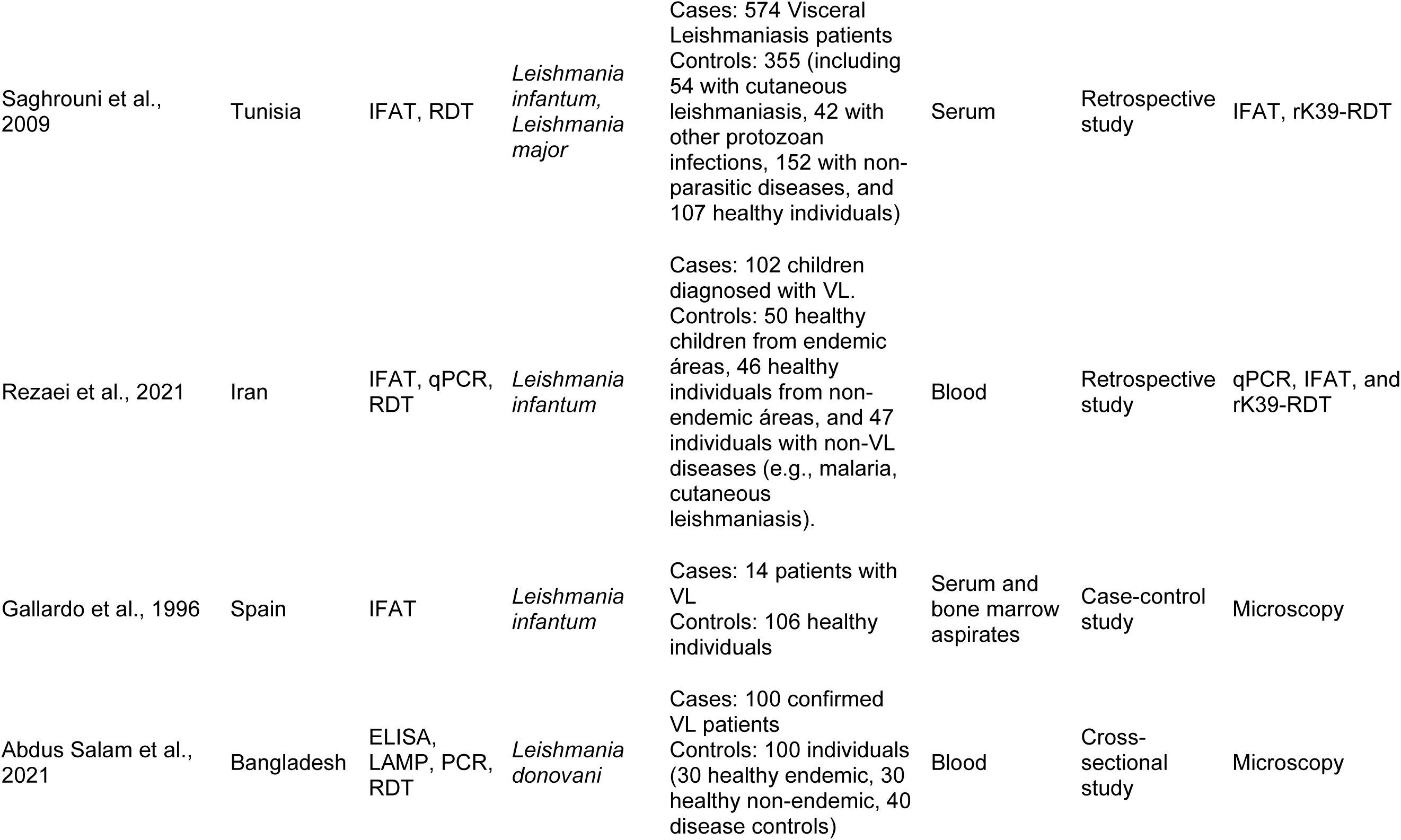
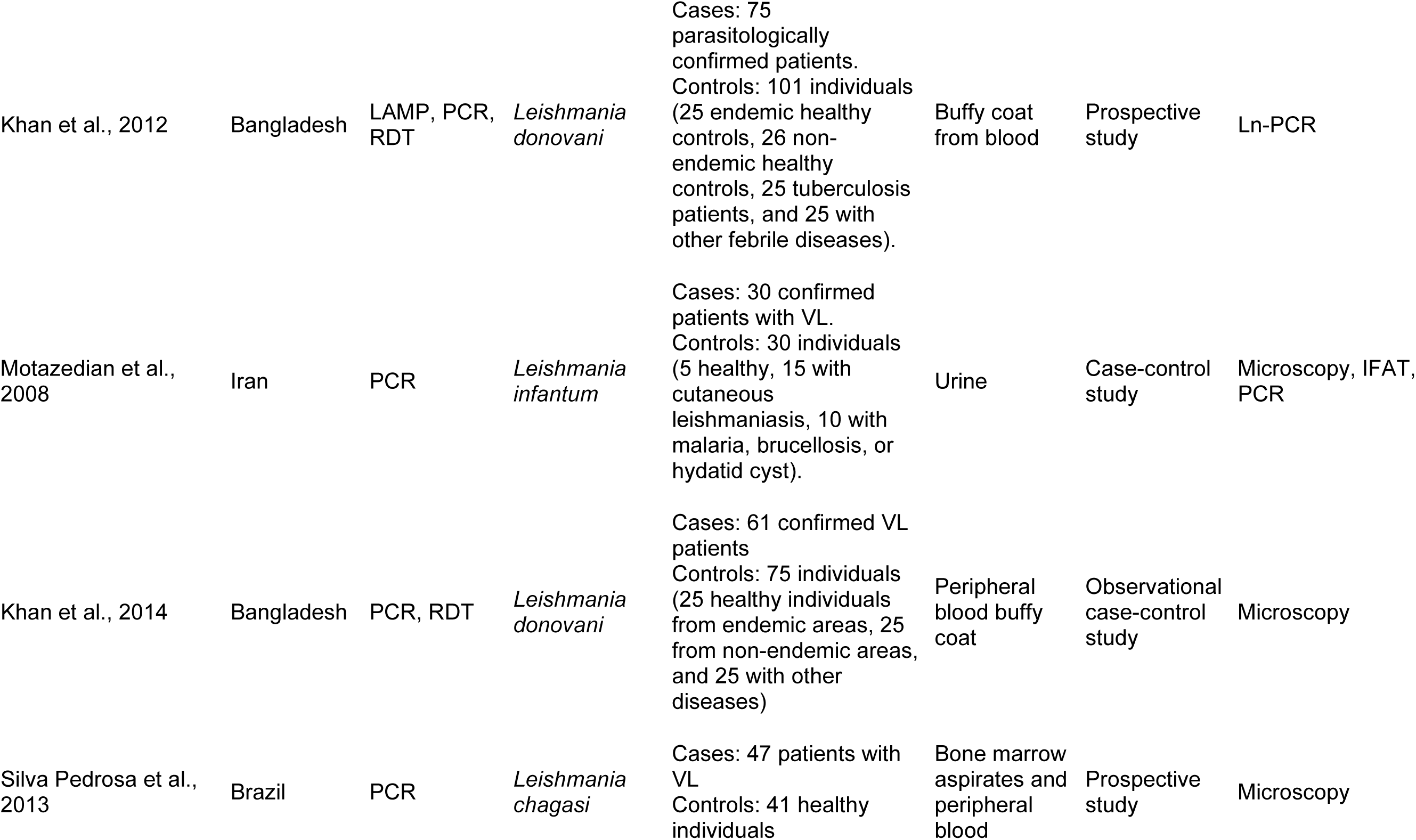
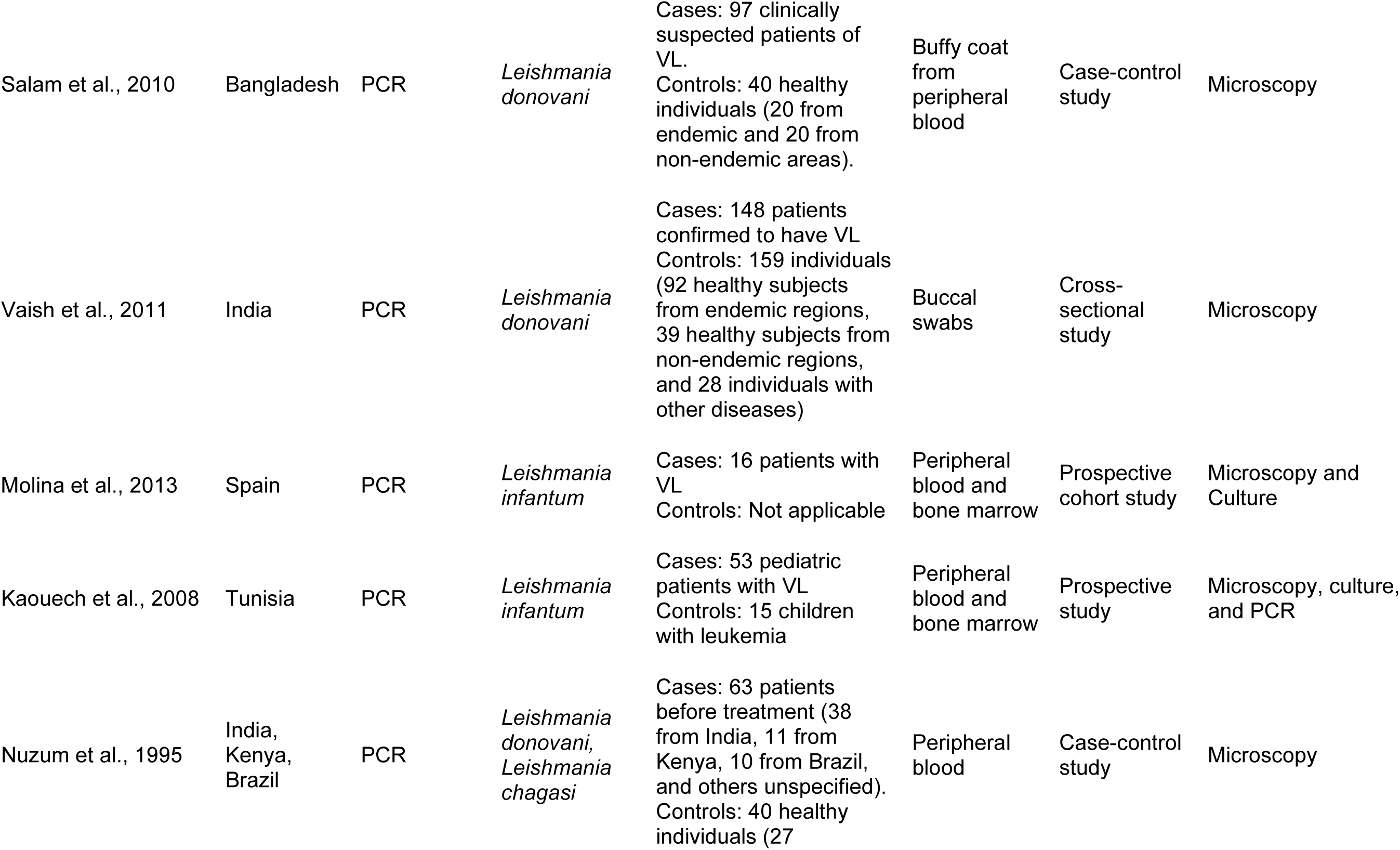
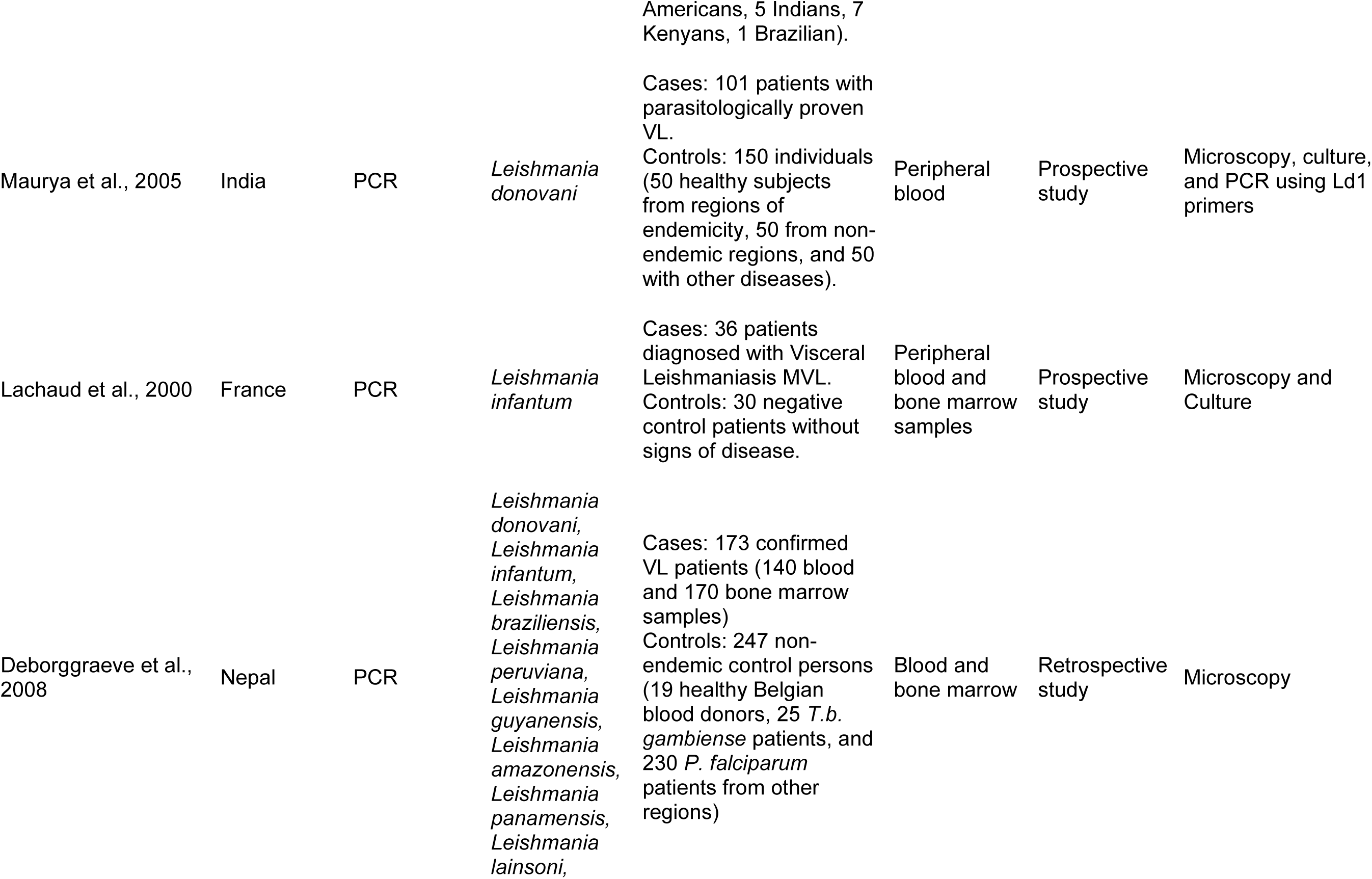
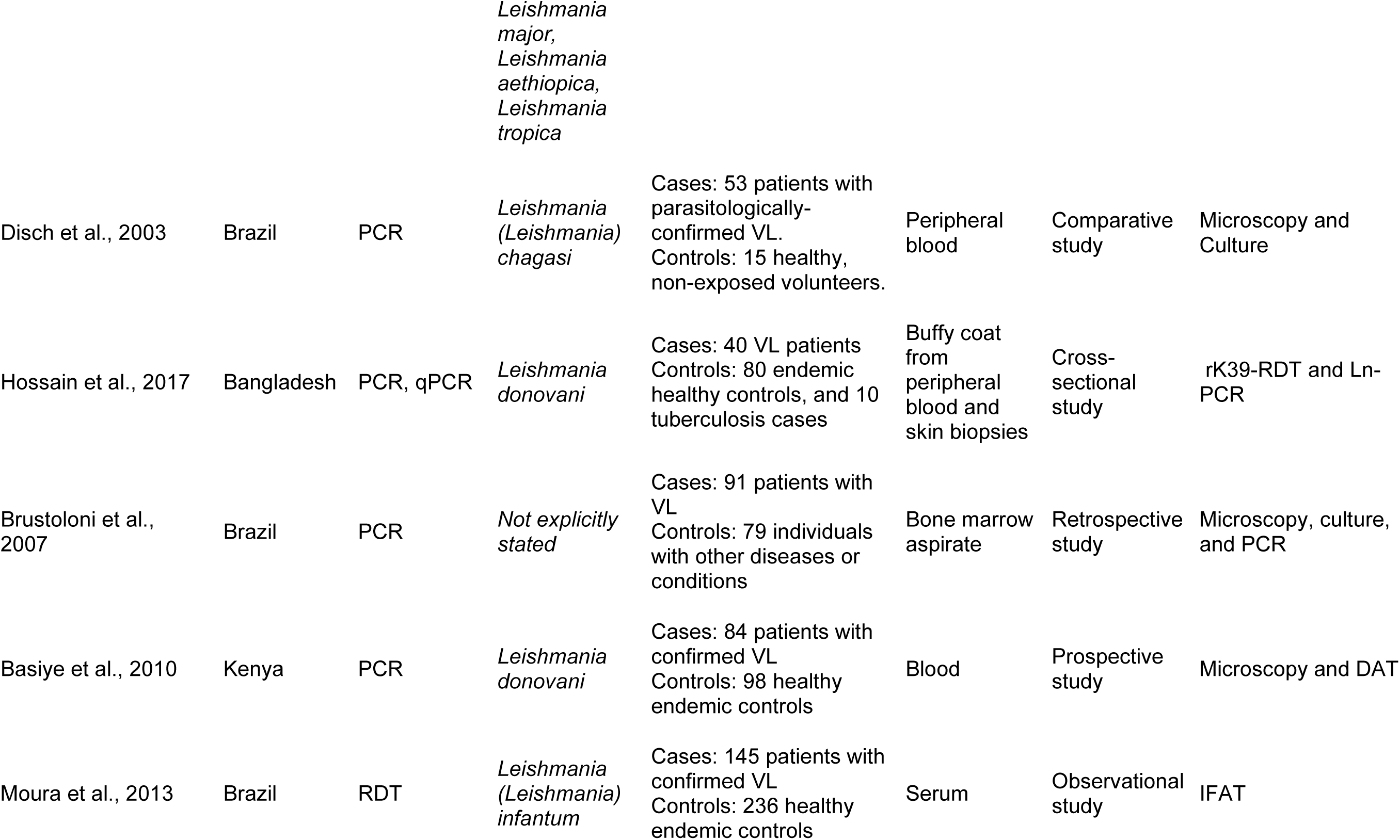
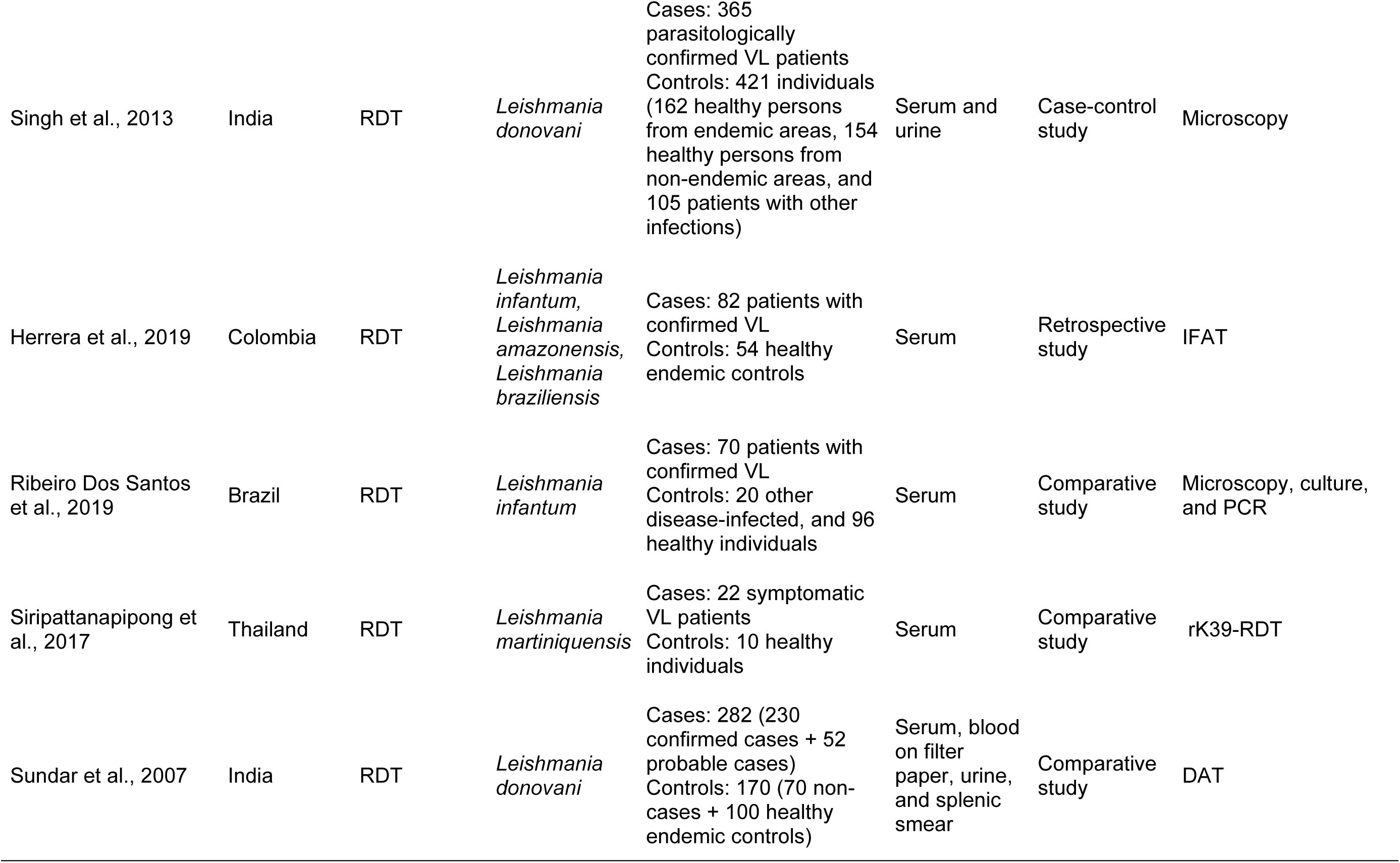
Main methodological aspects of studies on visceral leishmaniasis.

## Abbreviations

The following abbreviations are used in this review.

AUC: Area under the curve
AUC_FPR_: Area under the curve restricted to the false positive rates.
CI: Confidence interval
CL: Cutaneous leishmaniasis
DAT: Direct agglutination test
DNA: Deoxyribonucleic acid
DOR: Diagnostic likelihood ratio
ELISA: Enzyme-linked immunosorbent assay
FN: False negatives
FP: False positives
INPLASY: International Platform of Registered Systematic Review and Meta-analysis Protocols
IFAT: Indirect immunofluorescence assay
IHA: Indirect hemagglutination assay
ITS: Internal transcribed spacer
kDNA: Kinetoplast DNA
LAMP: Loop-mediated isothermal amplification
LST: Leishmanin skin test
LR−: Negative likelihood ratio
LR+: Positive likelihood ratio
MCL: Mucocutaneous leishmaniasis
MeSH: Medical subject headings
NCBI: National Center for Biotechnology Information
NLM: National Library of Medicine
PCR: Polymerase chain reaction
PRISMA: Preferred Reporting Items for Systematic Reviews and Meta-Analyses
qPCR: Real-time polymerase chain reaction
RDT: Rapid diagnostic test
ROC: Receiver operating characteristic
Se: Sensitivity
Sp: Specificity
sROC: Summary receiver operating characteristics
TN: True negatives
TP: True positives
VL: Visceral leishmaniasis
WB: Western blot
WHO: World Health Organization

